# Genetic and Etiological Insights from Automated Lumen Diameter Measurements in Carotid Ultrasounds of the UK Biobank

**DOI:** 10.1101/2025.01.07.25320106

**Authors:** Sofía Ortín Vela, Dennis Bontempi, Bianca Mazini, Leah Böttger, Olga Trofimova, Ian Quintas, David Presby, Ilaria Iuliani, Sven Bergmann

## Abstract

Carotid ultrasound is routinely used in clinical practice for non-invasive vascular anatomical and functional assessment. In particular, the carotid intima-media thickness (cIMT) is an important marker for quantifying atherosclerotic burden in the common carotid arteries (CCAs). Recent research suggests that several risk factors associated with higher cIMT, such as high blood pressure, can induce a compensatory increase in the carotid lumen diameter (cLD) of the CCAs. However, the genetic architecture of cLD and its association with other cardiovascular traits are still poorly understood. To address this gap, we trained a deep learning model to segment the carotid lumen from ultrasound images and developed an algorithm to measure the mean cLD. After quality control, we derived this trait for 18 808 genotyped individuals from the UK Biobank, comparing multiple measures of cLD from lateral and central views of the left and right carotid arteries. We then performed genome-wide association studies (GWAS) for cIMT, cLD and their ratio, revealing that cLD has an estimated heritability of 31%, substantially higher than that of cIMT (23%) and cIMT/cLD (14%). Finally, we conducted association and survival analyses and found that cLD was more strongly associated with several cardiovascular risk factors, such as hypertension and BMI, while cIMT/cLD performed the best in a prognostic setting after adjusting for demographics and common cardiovascular risk factors.

## Introduction

The right and left common carotid arteries (CCAs) supply the head, face, and neck with oxygenated blood (**Fig. 1a**). As changes in carotid morphology are closely linked to cerebrovascular risk, carotid ultrasound imaging is a standard tool for evaluating cardiovascular health, offering a non-invasive approach to characterise carotid anatomy and function ^1^. The main assessments in clinical settings are the measurement of carotid intima-media thickness (cIMT), i.e., the two innermost layers of the wall of an artery, widely recognised as a significant marker for atherosclerotic burden ^2–4^, and the detection of stenosis, where advanced plaque accumulation narrows the carotid lumen diameter (cLD), i.e., the inner width of the artery, and increases the risk of ischaemic stroke ^5^. However, lumen diameter does not necessarily decrease in the early stages of plaque formation: according to Glagov’s hypothesis, the CCAs can undergo outward remodelling in response to plaque buildup, effectively maintaining or even increasing cLD despite the presence of atherosclerosis. This serves as a compensatory mechanism to maintain blood flow despite structural changes and increased stiffness within the artery walls ^6,7^. More recent work suggests that similar enlargement of the lumen can also occur as part of a hemodynamic compensation response to increased systemic load, such as chronically elevated blood pressure, increased stroke volume, or obesity, even in the absence of significant plaque ^8,9^. Taken together, these mechanisms suggest that cLD may hold diagnostic value by reflecting both atherosclerotic and hemodynamic remodelling processes, capturing aspects of vascular adaptation that are not reflected by cIMT alone. However, its assessment has not been implemented as standard clinical practice, partly because its diagnostic utility has not been fully demonstrated, despite its known implication in compensatory vascular remodelling.

**Figure 1.**
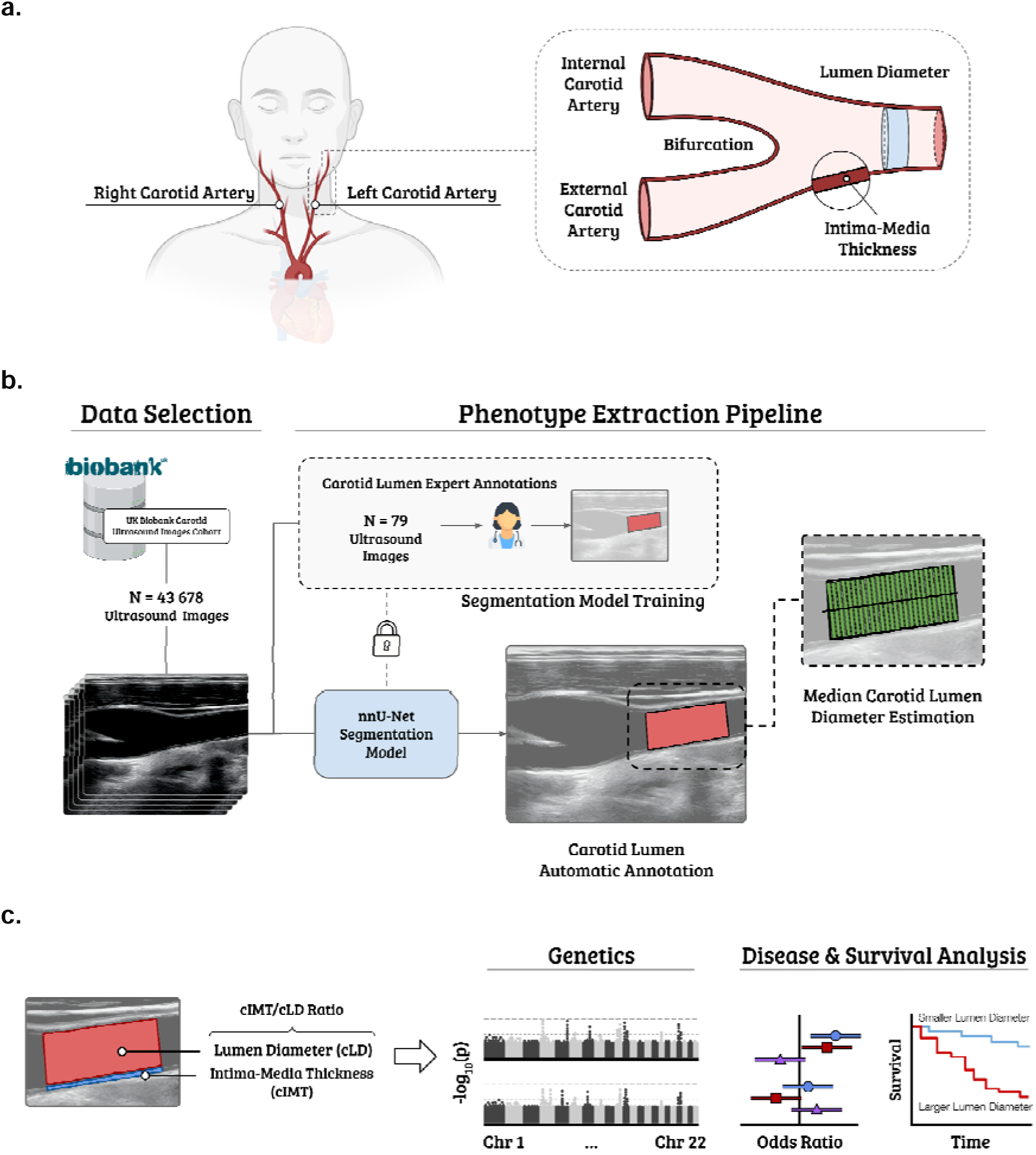
Study Overview. **a)** Anatomy of the carotid artery. The left and right carotid arteries bifurcate to the internal and external carotid arteries. The carotid intima-media thickness (cIMT) and the carotid lumen diameter (cLD) are measured before this bifurcation. **b)** We first trained a convolutional neural network (CNN) to segment the carotid from ultrasound still images automatically, using ground truth provided by a human annotator (N = 79 manual segmentations). We then used this CNN to process 43□678 ultrasound images of the left and right carotid, including central and lateral views, from the UK Biobank (UKB). Each segmentation was post-processed for quality control (QC) and then underwent classical image processing to estimate the cLD for each image by computing the median of multiple diameter measurements (green) perpendicular to the central axis of the segmentation mask. **c**) Finally, we studied the genetics of cLD, compared it to that of cIMT and their ratio, and evaluated the three phenotypes in terms of diagnostic and prognostic power.

A recent study suggested that increased cLD is associated with all cause mortality in the general population ^10^, and other studies ^11,12^ have explored the clinical significance of cLD for cardiovascular events. Two publications studied the genetic architecture of cLD: using genome-wide linkage analysis with data from 3 300 American Indian participants in the Strong Heart Family Study, Bella et al. ^13^ identified a locus influencing cLD on chromosome (Chr) 7 and suggested that cLD has a distinct genetic makeup from cIMT. Another study from Proust et al. ^14^ performed a GWAS involving a sample of 3 681 participants on the right carotid diameter, finding a significant association between cLD (and a trend for the external diameter) and the single-nucleotide polymorphism (SNP) rs2903692 mapping to the *CLEC16A* gene on 16p13. Despite these contributions, the genetic basis of cLD, its relationship with cIMT, and its potential diagnostic and prognostic utility remain insufficiently characterised, particularly in large population cohorts.

To address these gaps, we developed a fully automated analysis pipeline enabling high-throughput assessment of cLD. Specifically, we leveraged a deep convolutional neural network (CNN) trained on a dataset annotated by a clinical expert to segment a section of the carotid artery from ultrasound images and devised a method to measure the median cLD on such segmentations. Our tool allowed us to measure cLD phenotypes of 18 808 genotyped participants of the UK Biobank (UKB), establishing the largest dataset to date. This enabled us to investigate the correlations between cLD measures from the left and right carotid, as well as their lateral and central view. We then explored the genetic architecture of these measures through genome-wide association analyses (GWAS) and subsequent post-processing analysis, pointing to overall mean cLD as the most robust measure. Comparing the latter with mean cIMT revealed that these two ultrasound-derived phenotypes have a higher genetic than phenotypic correlation, yet several distinct genetic associations, supporting the notion of non-identical (genetic) mechanisms modulating these phenotypes. Building on the complementary nature of these two phenotypes, we propose the ratio cIMT/cLD as an additional, normalised phenotype to capture distinct aspects of the CCA’s characteristics that may not be fully represented by the two phenotypes individually. Furthermore, we investigated the associations of cIMT, cLD, and their ratio with diseases and associated risk factors, and evaluated their prognostic power across several endpoints, including cardiovascular events and all-cause mortality. From these analyses emerged that, while cIMT and cLD are strongly associated with risk factors and diseases, independent of demographics, their ratio performs the best in a prognostic setting after adjusting for demographics and common cardiovascular risk factors. Finally, by investigating the aforementioned carotid phenotypes in males and females separately, we observed that some of them showed significant diagnostic and prognostic value in one sex only, highlighting the importance of considering sex-specific effects when interpreting carotid phenotypes. In summary, this study combines deep-learning–derived measurements of cLD with established cIMT phenotypes to examine their genetic architecture, interrelationships, and clinical significance in a large population cohort. By introducing the cIMT/cLD ratio, we sought to capture complementary aspects of carotid structure that could enhance risk stratification beyond individual measures. Our results demonstrate that these carotid phenotypes capture distinct biological and clinical information, and that the cIMT/cLD ratio may improve prognostic value over cIMT when accounting for demographic and cardiovascular risk factors.

## Results

For our analysis we processed 21 838 left and 21 840 right carotid ultrasound DICOM image series, derived from 20 031 and 20 033 subjects of the UKB, respectively. Each series includes several still images captured from ultrasound movies during diastole. These images include views of the central and lateral regions of the right and left CCAs from which we extracted the *right lateral*, *right central*, *left lateral*, and *left central cLD*. From these four primary cLD measurements, we computed five additional derived cLD phenotypes, namely the *left*, *right*, *central*, *lateral*, and (overall) *mean cLD* by averaging across the relevant primary cLD measurements. Furthermore, we derived the *mean cIMT* and the *mean cIMT over mean cLD*. After quality control our study our sample size was N=18 808. An overview of our study design is presented in **Fig. 1b** and detailed in the Methods section.

### Correlations and Heritabilities of different Carotid Lumen Diameter measures

We first adjusted the nine cLD measures by regressing out the effects of common covariates (see Methods). We then computed pairwise phenotypic (Pearson) correlations between the corrected phenotypes (**Fig. 2a**, lower triangle). We observed high phenotypic correlations across the different cLD phenotypes (r ∈ [0.69, 0.98]). Central and lateral cLD phenotypes correlated more strongly with each other (r = 0.9) than the left and right cLDs (r = 0.73), **Suppl. Table 1a**).

**Figure 2.**
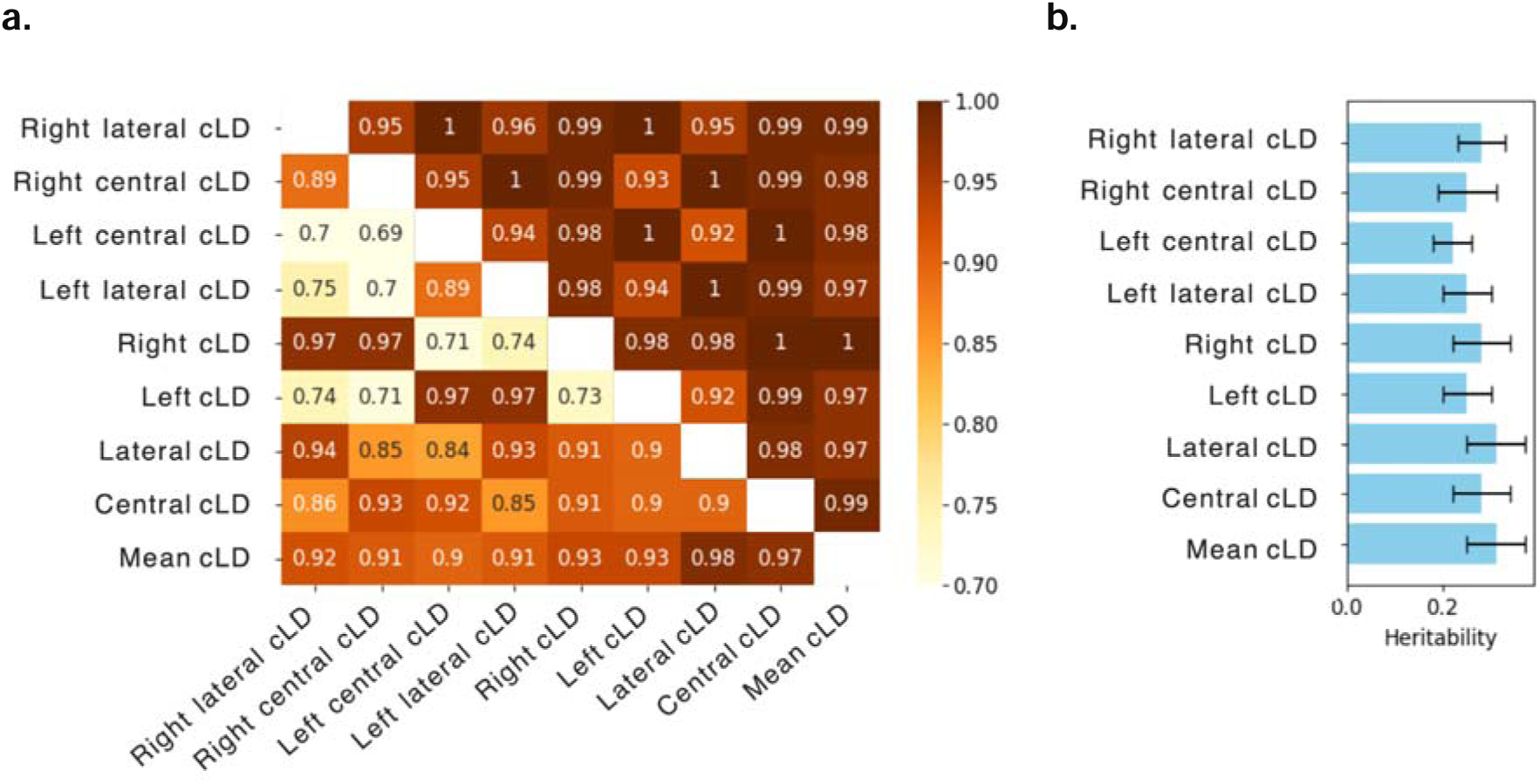
Correlation and heritability analysis. **a)** Phenotypic (lower-left triangle) and genetic (upper-right triangle) correlations of cLD phenotypes. Phenotypes were corrected for covariates before computing phenotypic correlation, while for the GWAS, phenotypes were rank-normalised and regressed on genotypes and covariates (see Methods). **b)** Heritability (h²) estimates and their standard error (error bars). The phenotype h² were estimated using LDSR. The sample sizes are as follows: Right lateral cLD (N=17 951), right central cLD (N=17 839), left central cLD (N=17 758), left lateral cLD (N=17 680), right cLD (N=18 634), left cLD (N=18 584), lateral cLD (N=18 686), central cLD (N=18 689), and mean cLD (N=18 808).

To investigate the genetic basis of cLD, we performed GWAS for each measure and analysed the resulting summary statistics using Linkage Disequilibrium Score Regression (LDSR) ^15,16^ to estimate cross-phenotype genetic correlations (**Fig. 2a**, upper triangle) and heritabilities (h²; **Fig. 2b**). Genetic correlations followed similar patterns to their phenotypic counterparts but were generally higher (r ∈ [0.92,1]) (**Suppl. Table 1a**). Manhattan plots summarising significant genetic loci are shown in **Suppl. Fig. 4**. Heritability estimates were similar for the nine cLD measures (**Fig. 2b**), with mean cLD and lateral cLD showing the highest values (h² = 0.31 ± 0.06). The smallest h² was observed for the left central cLD (h² = 0.22 ± 0.04) (**Suppl. Table 3**).

### Genes and Pathways influencing different Carotid Lumen Diameter measures

To identify genes associated with each phenotype, we used PascalX ^17,18^. The number of genes associated with cLD varied across phenotypes, ranging from 6 to 47 (**Fig. 3a**, diagonal and **Suppl. Data**). Notably, the left central cLD showed the fewest associated genes, consistent with its lowest h² estimate (22%, see Fig. 2b). Similarly, left lateral cLD and left cLD had fewer associated genes, compared to the other cLD measures, consistent with their smaller h² values. While lateral cLD and mean cLD obtained both the highest h² values, lateral cLD had slightly more associated genes. Typically the majority of the individually associated genes were shared between cLD phenotypes (off-diagonal of **Fig. 3**).

**Figure 3.**
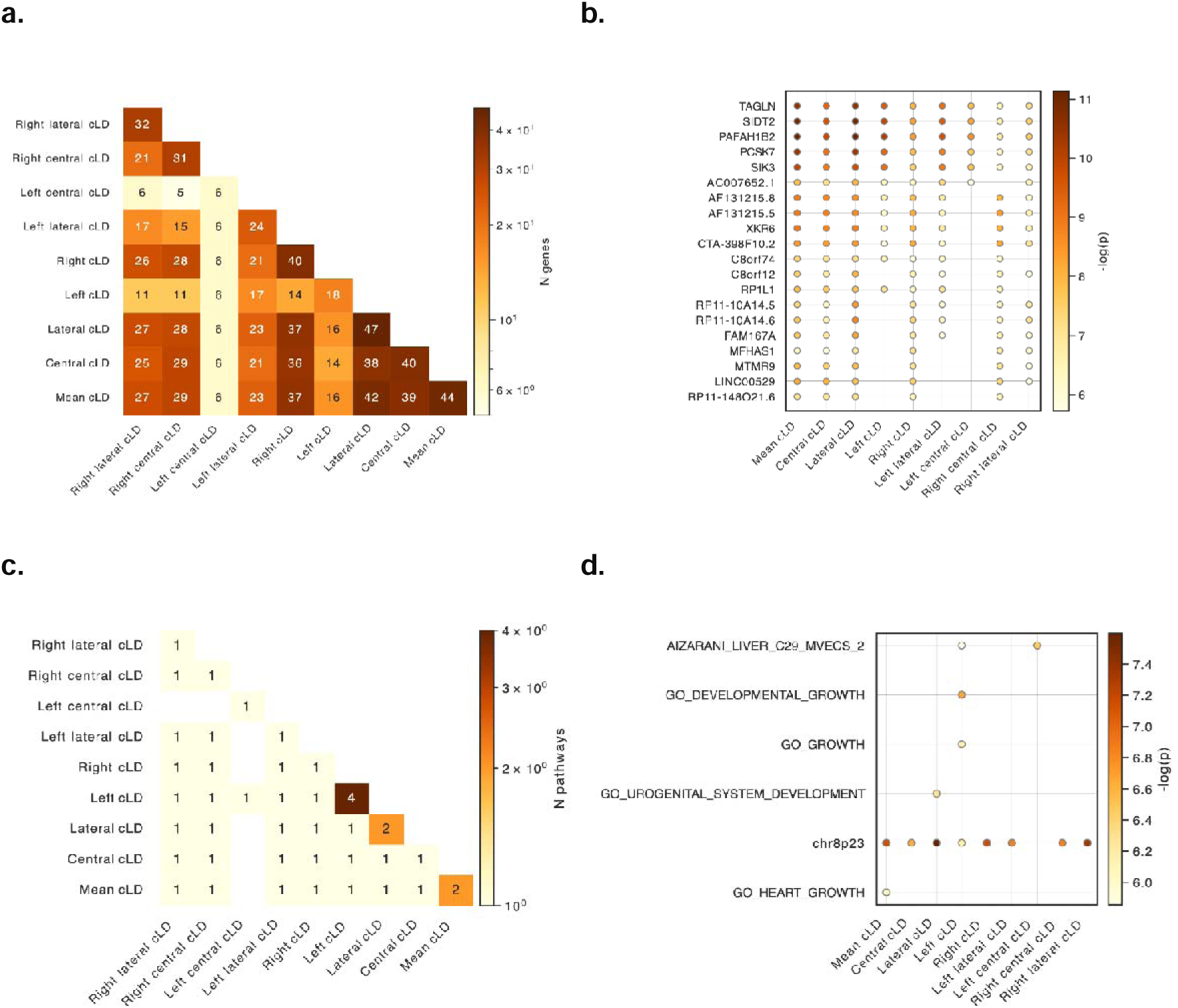
Gene and pathway analyses. **a)** Gene-scoring intersection showing the number of genes in common between cLD phenotypes. The diagonal shows the number of genes associated with each phenotype. **b)** Name of the most frequent genes across the cLD phenotypes. **c)** Pathway-scoring intersection showing gene sets in common between cLD phenotypes. The diagonal shows the number of pathways associated with each phenotype. **d)** Name of the most frequent gene sets across the cLD phenotypes.

Five genes were significantly associated with all cLD phenotypes: *TAGLN, SIDT2, PAFAH1B2, PCSK7*, and *SIK3* (**Fig. 3b**). Other genes, such as *C8orf12* and *RP11-10A14.5,* were associated with all the cLD phenotypes except the left cLD and left central cLD (see **Suppl. Data** for the complete lists of genes).

Using PascalX, we identified annotated gene sets (i.e., “pathways”) enriched with high-scoring genes (**Fig. 3c, d**). Three phenotypes were associated with more than one pathway: Left (4), lateral (2), and mean (2) cLD. Notably, the gene set *‘chr8p23’* was associated with all phenotypes except the left central cLD (see **Suppl. Data** for the complete list of pathways).

### Comparison of Carotid Lumen Diameter and Carotid Intima-Media Thickness

To compare cLD phenotypes with cIMT, we focused on mean cLD as the representative for all cLD phenotypes due to its comprehensive nature, being the average of all cLD measurements, and its high heritability. Additionally, we also considered the ratio of mean cIMT over mean cLD as a composite phenotype presenting a “normalised” cIMT.

The phenotypic correlation between mean cLD and mean cIMT was moderate (Pearson’s r = 0.37, see **Fig. 4a**). In contrast, the genetic correlation was substantially higher (0.58 ± 0.10). For the ratio cIMT/cLD, the phenotypic correlations were −0.23 with mean cLD and 0.81 with mean cIMT. The corresponding genetic correlations were −0.11 ± 0.11 and 0.74 ± 0.05, respectively (**Suppl Table 2a–b**).

**Figure 4.**
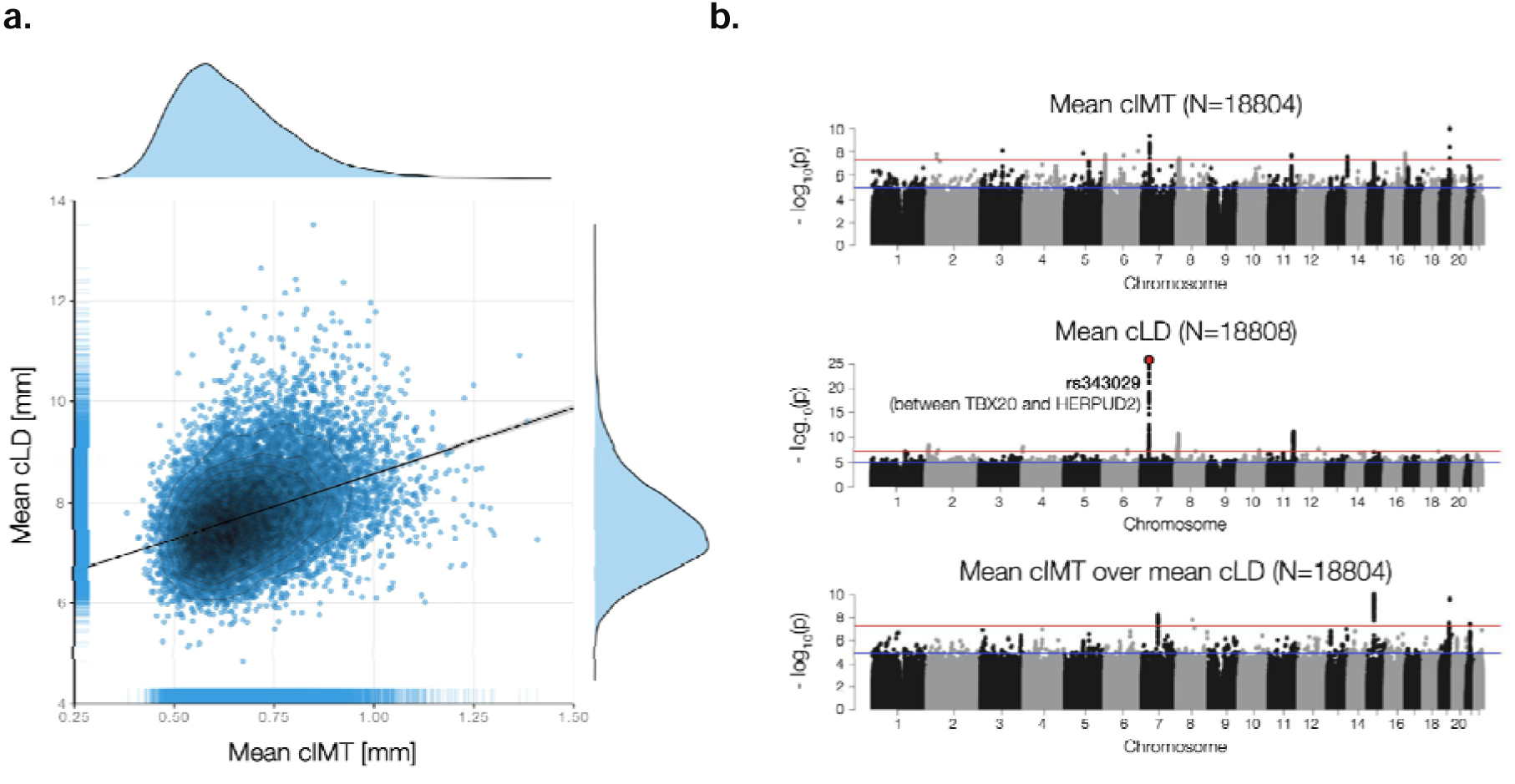
>Mean cLD, mean cIMT, and ratio comparison. **a)** Scatter-plot of *mean cIMT* against *mean cLD*. **b)** Manhattan plots of *mean cIMT*, *mean cLD*, and *mean cIMT over mean cIMT*. The sample sizes for each analysis are reported on top of each figure and are identical for the phenotypes shown in panel **a** and the GWAS summary statistics used for the genetic correlation analysis.

Manhattan plots revealed heterogeneity in genetic signals among the phenotypes (**Fig. 4b**). Despite identical sample sizes, mean cLD exhibited stronger genetic association signals than mean cIMT or cIMT/cLD. For mean cLD, the most significant SNPs were located at the start of Chr 7 (rs343029; p=2.73E-26), with additional associations on other chromosomes, including Chr 8 (rs7838131; p=3.20E-08, among others) and 11 (rs111677878; p=8.62E-12). Mean cIMT showed its strongest associations at the start of Chr 7 (rs342988; p=4.25E-10) and at the end of Chr 19 (rs1065853; p=8.99E-11), while cIMT/cLD had key signals on Chr 7 (rs7792074; p=5.21E-09, mid-region), 15 (rs625034; p=8.16E-11), and 19 (rs111688353; p=3.11E-08, rs1065853; p=2.10E-10), among others. For more details, see **Suppl. Table 2**.

Heritability estimates, using LDSR ^15^, showed mean cLD (h² = 0.31 ± 0.06) as the most heritable, followed by mean cIMT (0.23 ± 0.04) and cIMT/cLD (0.14 ± 0.03) (**Table 1a**; **Suppl. Table 3**). Mean cLD was also associated with the highest number of genes (44), compared to mean cIMT (14) and their ratio (2). 11 genes were significantly associated with both mean cLD and mean cIMT, including *XKR6*, *LINC00529*, *MTMR9*, *FAM167A*, *C8orf12*, *LINC00208*, *BLK*, and *MFHAS1* (**Suppl. Data**). The PascalX cross-GWAS coherence test ^18^ resulted in more coherent than anti-coherent signals between the three phenotypes (**Table 1b**; **Suppl. Data**). In particular, cLD and cIMT shared 47 coherent genes, which included the 11 genes from the intersection of their individual gene scores and 36 other genes, such as *RP1L1*, *GMDS*, *ERI1*, *SGK223*, *MSRA*, *SOX7*, *GATA4*, and *TMEM170A.* Notably, 43 of the 47 coherent genes are located on 8p23.1. Anti-coherent signals were *ELN* and *RP11-731K22.1.* The ratio between cIMT and cLD shared coherent signals with cIMT (*LINC00670*) and cLD (8 genes, including *LINC00670*, *TMEM170A*, and *CBFA2T3*). Anti-coherent signals included *PAFAH1B2* (with cIMT) and *ELN* (with cLD).

**Table 1.** Genetic *mean cLD*, *mean cIMT*, and *ratio* comparison. a) The heritability estimate (h²) was obtained using LDSR, and the number of genes and pathways was obtained using PascalX. **b)** Number of genes showing coherent (top right) or anti-coherent (bottom left) signals between pairs of phenotypes, obtained using PascalX cross-scoring.

| Phenotype | $h^2$ (std) | Number of genes | Number of pathways |
| --- | --- | --- | --- |
| Mean cIMT | 0.23 (0.04) | 14 | 1 |
| Mean cLD | 0.31 (0.06) | 44 | 2 |
| Mean cIMT over mean cLD | 0.14 (0.03) | 2 | 0 |

**Table 1 | Genetic mean cLD, mean cIMT, and ratio comparison.**
|  | Mean cIMT | Mean cLD | Mean cIMT over mean cLD |
| --- | --- | --- | --- |
| Mean cIMT |  | 47 | 1 |
| Mean cLD | 2 |  | 8 |
| Mean cIMT over mean cLD | 4 | 2 |  |

For cIMT, we identified a single gene set, *’chr8p23’*, while two gene sets were found for mean cLD, namely *’chr8p*23’ and *’GO HEART GROWTH*’ (**Table 1a**; **Suppl. Data**).

### Disease Association and Survival Analysis of Carotid Phenotypes

We investigated the associations of our three carotid phenotypes with diseases and associated risk factors by using logistic and linear regression models, respectively. We estimated the effects of cIMT, cLD, and their ratio separately, adjusting for sex, age, and standing height.

The three carotid measures were significantly associated with diastolic blood pressure (DBP) regardless of the subjects receiving medication for hypertension (**Fig. 5a**). While cIMT was positively correlated with DBP for un-medicated subjects, we observed an inverse correlation for medicated subjects. All three carotid phenotypes were also associated with pack-years of smoking, arterial stiffness, and resting heart rate (RHR) (p<0.05 after Benjamini-Hochberg correction), with the latter exhibiting an inverse association with cIMT and the ratio. While the cIMT/cLD ratio was not associated with BMI and systolic blood pressure (SBP) in subjects medicated for hypertension, both cIMT and cLD were positively associated. Except for arterial stiffness and pack-years of smoking, standardised effect sizes were higher, more than doubling in some cases, for cLD compared to the other two phenotypes (standardised β for DBP, un-medicated subjects: 0.1605, 95%CI 0.1384–0.1827 for cLD, p<0.0001; 0.04202, 95%CI 0.02156–0.06249 for cIMT, p<0.0001; standardised β for BMI: 0.2339, 95%CI 0.2181–0.2496 for cLD, p<0.0001; 0.1080, 95%CI 0.09279–0.1233 for cIMT, p<0.0001; see **Suppl. Table 11a**). Sex-stratified analyses showed no pronounced sex-specific effect for any of the phenotype-risk factor pairs besides cIMT and SBP in un-medicated subjects, and cIMT/cLD ratio and BMI (see **Suppl. Fig. 6a, Suppl. Fig. 7**, and **Suppl. Table 12a**).

**Figure 5.**
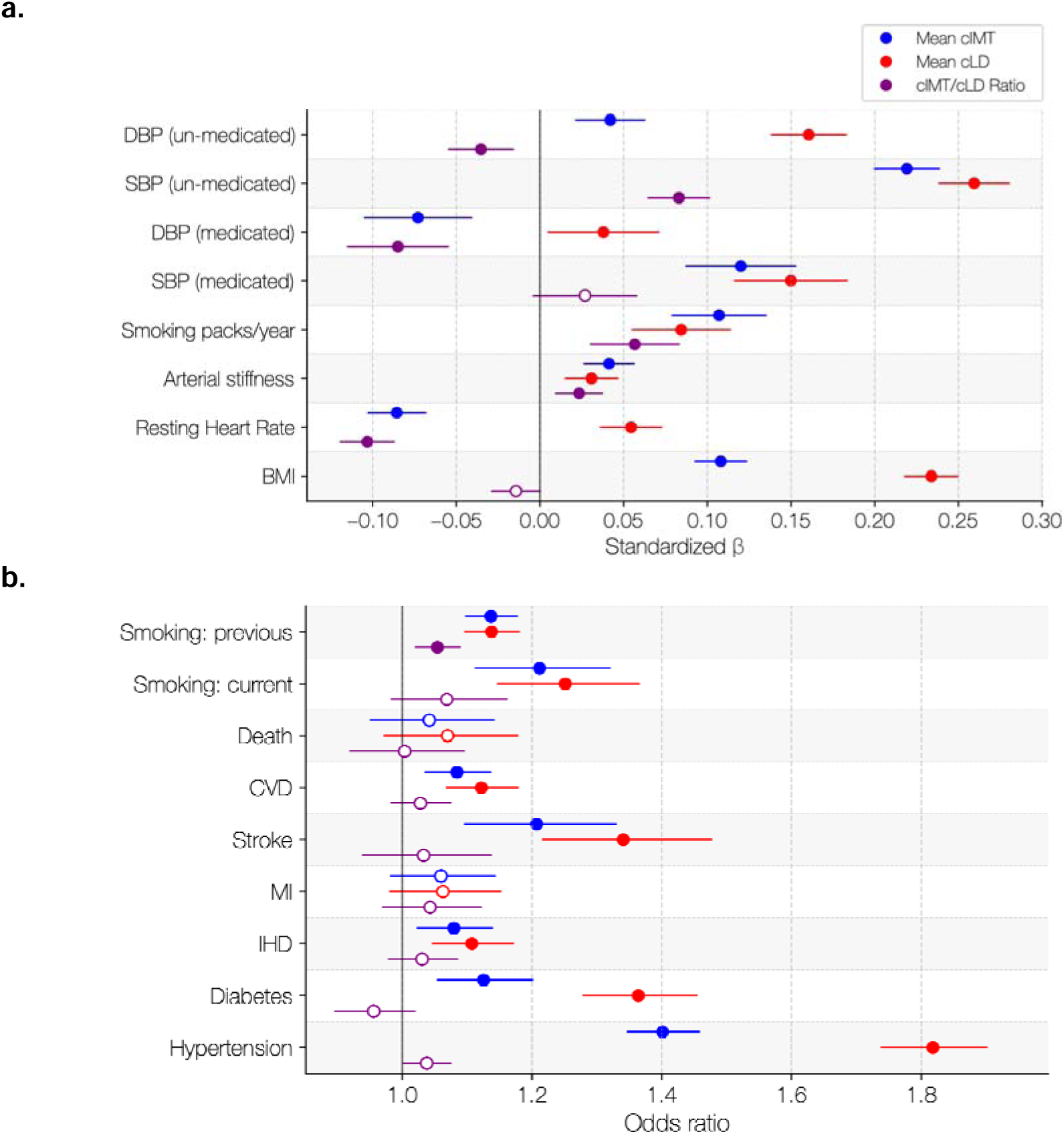
Clinical associations of mean cIMT, mean cLD, and their ratio. **a)** Standardised regression coefficients from models assessing the independent associations of each phenotype with continuous risk factors (adjusted for age, sex, and height; N=18 767). **b)** Odds ratios from analogous models evaluating associations with major categorical risk factors and prevalent disease outcomes (adjusted for age, sex, and height; N=18 767). Abbreviations: SBP/DBP: systolic/diastolic blood pressure; BMI: body mass index; CVD: cardiovascular diseases; MI: myocardial infarction; IHD: ischaemic heart disease.

In our analysis of associations with major categorical risk factors and prevalent disease outcomes, the only significant association shared by all three carotid phenotypes was with previous smoking. cLD and cIMT showed significant associations with current smoking, prevalent cardiovascular and stroke events, prevalent ischaemic heart disease, diabetes, and hypertension. Notably, we found consistently higher odds ratios (OR) for cLD. In particular, for diabetes and hypertension, ORs doubled for cLD compared to cIMT (OR for diabetes: 1.364, 95%CI 1.279–1.454 for cLD, p<0.0001; 1.125, 95%CI 1.055–1.200 for cIMT, p<0.001; OR for hypertension: 1.818, 95%CI 1.739–1.901 for cLD, p<0.0001; 1.401, 95%CI 1.347–1.458 for cIMT, p<0.0001; see **Fig. 5b** and **Suppl. Table 11b**). In the sex-stratified analyses, the association of current smoking and stroke with the carotid phenotypes was much stronger in males. For most of the other binary outcomes, the associations were stronger in females, particularly for cLD, despite the reduced prevalence of cardiovascular events (see **Suppl. Fig. 6b**, **Suppl. Fig. 7, Suppl. Table 7b**, and **Suppl. Table 12b**).

To assess the prognostic power of cIMT, cLD, and their ratio, we examined their capacity to predict subsequent cardiovascular events and all-cause mortality using Cox proportional hazard (PH) models. We fitted a series of models to evaluate how each phenotype predicted different clinical endpoints, while adjusting for basic covariates and common cardiovascular risk factors.

We first examined the prognostic ability of cIMT, cLD, and their ratio using Kaplan-Meier survival analyses. For cardiovascular events, we observed clear separation across different groups, with cIMT providing the strongest stratification, cLD performing similarly, and their ratio showing the weakest separation (left panel of **Fig. 6a**). Overall, higher values were consistently associated with increased event rates (log-rank p<0.0001 for all three phenotypes). The same pattern emerged for stroke events (log-rank p<0.001 for cIMT/cLD ratio, p<0.0001 for the others; right panel of **Fig. 6a**) and all-cause mortality, where participants in higher strata showed poorer survival compared with those in lower strata (log-rank p<0.0001 for all three phenotypes; **Suppl. Fig. 9a**). To quantify these associations further, we fitted Cox PH models using individuals with values below the median, for each phenotype respectively, as reference. For cardiovascular events, after adjusting for demographics (age and sex), participants with values above the median for cIMT, cLD, or their ratio showed an increased risk (HR up to 1.274, 95%CI 1.101–1.475 for cIMT, p<0.01; left panel of **Fig. 6b**). After further adjusting for cardiovascular risk factors (BMI, systolic blood pressure, smoking status, diabetes, blood pressure medications, and previous cardiovascular events), the significant association remained only for the cIMT/cLD ratio (HR=1.188, 95%CI 1.018–1.388, p<0.05; left panel of **Fig. 6c**), suggesting it to be prognostic, independent of some of the most common cardiovascular risk factors. We observed similar results for stroke events (HR up to 1.783, 95%CI 1.258–2.527 for cIMT, p<0.01 after adjustment for demographics; HR=1.527, 95%CI 1.018–1.388 for the cIMT/cLD ratio, p<0.05 after full adjustment; right panels of **Fig. 6b–c**). Associations with all-cause mortality were not significant after adjustment for demographics and further adjustment for common cardiovascular risk factors (**Suppl. Fig. 8b–c**). Using carotid phenotypes as continuous predictors, all the phenotypes were significantly associated with cardiovascular events after adjusting for demographics (left panel of **Suppl. Fig. 9a**), but only cIMT and the cIMT/cLD ratio remained significant after further adjustment for cardiovascular risk factors (right panel of **Suppl. Fig. 9a**). For stroke events, cIMT and cLD were significantly associated with this outcome after adjusting for demographics and demographic plus cardiovascular risk factors (**Suppl. Fig. 9b**). Similarly to the group analysis, none of the markers were associated with all-cause mortality after adjusting for demographics, and for demographics plus common cardiovascular risk factors (**Suppl. Fig. 9c**). Finally, we ran the same analysis stratifying the cohort by sex. Kaplan-Meier curves revealed a clear separation (log-rank from p<0.01 to p<0.0001) between risk groups for both sexes and all the carotid phenotypes for cardiovascular events (**Suppl. Fig. 10a**). Although cLD in females and all the phenotypes in males were significantly associated with this endpoint when adjusting for age (**Suppl. Fig. 10b**), the statistical significance of this association was lost after adjusting for cardiovascular risk factors (**Suppl. Fig. 10c**). For stroke events, all three carotid phenotypes showed good stratification power (log-rank from p<0.01 to p<0.0001; **Suppl. Fig. 11a**) with the exception of the cIMT/cLD ratio in females. Unlike for cardiovascular events, in males, cIMT and cIMT/cLD ratio remained significantly associated with stroke after adjusting for age and cardiovascular risk factors, with the HRs for males trending substantially higher than the ones for females (**Suppl. Fig. 11b–c**), consistent with the sex-stratified disease association analysis. With all-cause mortality as the endpoint, all the phenotypes stratified the subjects into different risk groups (log-rank from p<0.01 to p<0.0001; **Suppl. Fig. 12a**), but only cLD in females was significantly associated with the endpoint after adjusting for age and all the other cardiovascular risk factors (**Suppl. Fig. 12b–c**).

**Figure 6.**
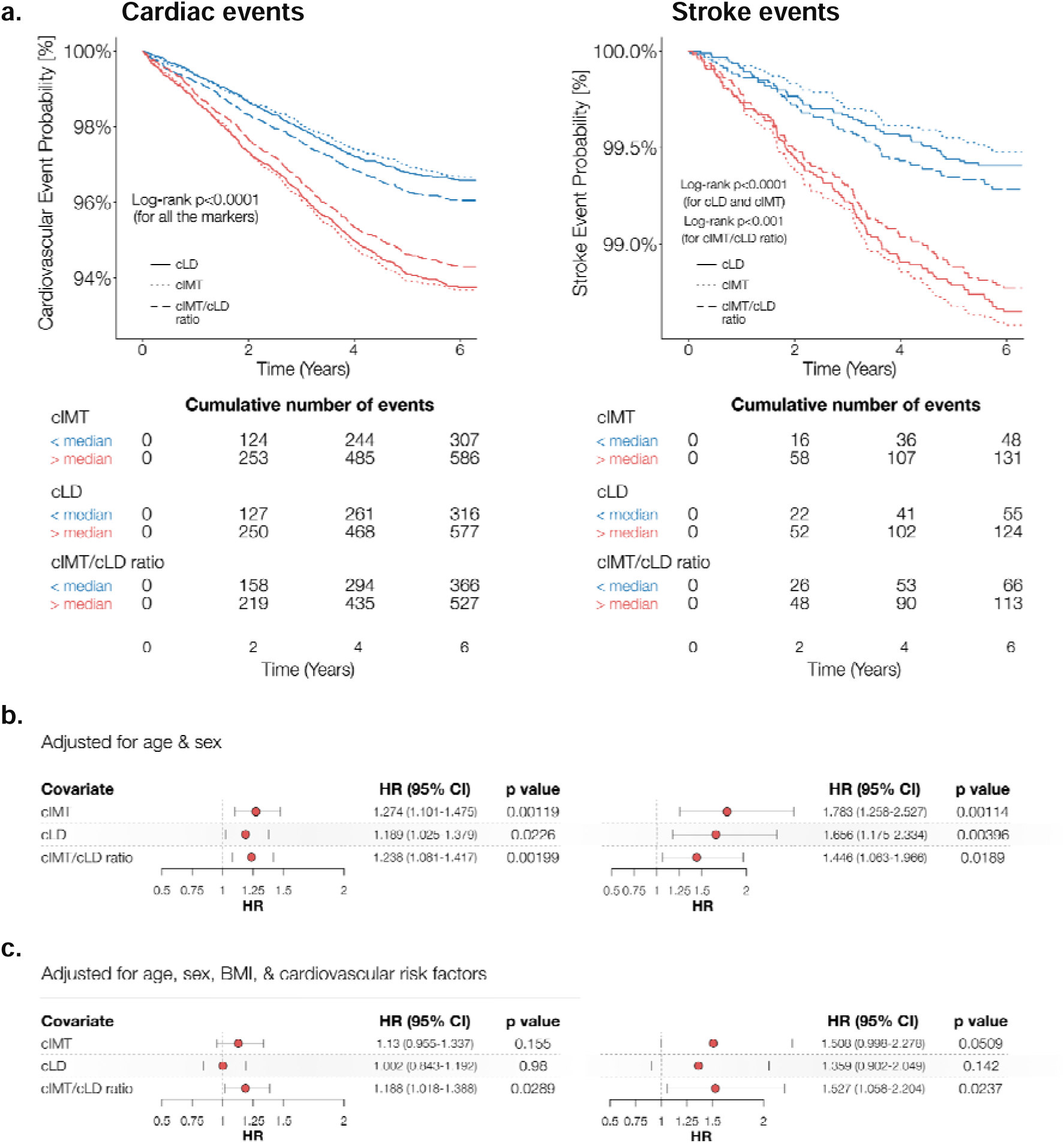
Prognostic performance of mean cIMT, mean cLD, and their ratio. **a)** Kaplan-Meier survival analysis of the three different phenotypes for cardiac (left) and stroke (right) events (N=18 767). The two groups were obtained by splitting the data on the median of the respective phenotype. **b)** Forest plots for the three different phenotypes for cardiac and stroke events. Here, the hazard ratios (HR) are calculated relative to individuals with phenotype values below the median. The HRs and the p-values were obtained from three independent Cox models (one per phenotype) adjusted for age and sex (N=18 767; total number of cardiac events: 893; total number of stroke events: 179). In **c)**, we additionally adjusted for BMI, systolic blood pressure, smoking status, diabetes, blood pressure medications, and previous cardiovascular events (N=15 011 after excluding non-complete observations; total number of cardiac events: 685; total number of stroke events: 129).

## Discussion

This study aimed to measure the carotid lumen diameter (cLD) in a large population with the following three goals: (i) elucidate the relationships between the left and right cLD and assess the potential impact of ultrasound orientation; (ii) compare cLD to the well-established carotid intima-media thickness (cIMT) phenotype and with their ratio (cIMT/cLD); and (iii) unravel their common and distinct genetic architectures and disease associations. By employing a convolutional neural network to segment carotid ultrasound images and incorporating image processing techniques, we automated the extraction of the cLD phenotypes, overcoming limitations in prior studies such as small sample sizes and labour-intensive methodologies. The resulting dataset for 18 808 genotyped participants of the UK Biobank (UKB), the largest of its kind, enabled a robust investigation of the genetic and phenotypic relationships of the cLD and cIMT/cLD, revealing that both provide complementary information to cIMT, with distinct genetic architectures and phenotypic patterns. Furthermore, we evaluated the clinical relevance of cLD, cIMT, and their ratio by examining their associations with prevalent disease and key cardiovascular risk factors, as well as their ability to predict future events. All three phenotypes showed significant associations with major cardiometabolic conditions independent of demographic influences. When adjusted for major cardiovascular risk factors, the cIMT/cLD ratio demonstrated a better prognostic performance across multiple endpoints, including incident cardiovascular events and stroke. Taken together, these results show that carotid structural phenotypes encode complementary information about vascular state and disease risk, supporting their utility as markers of vascular health. While cIMT and cLD independently associate with prevalent disease and risk factors, their ratio may offer additional prognostic insight by better reflecting the status of cardiovascular remodelling. Overall, these observations support the value of incorporating simple carotid imaging metrics into cardiovascular risk assessment frameworks to refine early detection of vascular dysfunction.

Studying the different cLD phenotypes we derived from ultrasound images, we found that, while the left and right cLD were highly correlated (<75%), this correlation was still substantially below that between the lateral and central views of the same carotid (89%), indicating a sizable left-right variability. We also noticed that the left cLD had a substantially lower heritability (22%) and fewer associated genes (6) than the right cLD phenotype (28% and 40, respectively), suggesting that it may undergo a stronger modulation by environmental effects. This left-right asymmetry may be due to the different anatomical origins of CCAs, with the left one typically arising directly from the aortic arch and the right one usually originating from the brachiocephalic trunk. Due to this different origin, the left common carotid artery is often longer than the right, which may contribute to differences in blood flow dynamics between the two sides ^19^ and could play a role in the formation of carotid atherosclerosis ^20^, with a higher prevalence in the left carotid ^4,21^.

Our GWAS significantly advances our understanding of cLD genetics, which, to the best of our knowledge, has only been explored in two previous studies with substantially smaller sample sizes. The linkage study by Bella et al. ^13^ found a significant SNP influencing cLD on Chr 7 at 120 centimorgans (cM), as well as a suggestive linkage on Chr 12 at 153 cM and 9 at 154 cM. Notably, while we found significant signals for rs343029 and rs11108966 on Chr 7 and 12, respectively, as plausible candidates for the respective linkage regions, we did not observe any significant associations on Chr 9. The GWAS by Proust et al. ^14^ found a marginally significant association (p=4E-7) for the right internal carotid diameter (rs2903692, Chr 16) mapping to the gene *CLEC16A*, which we did not replicate for our right or mean cLD (possibly due to the different population, namely American Indian participants in the *Strong Heart Family Study*, they studied).

Here, with a significantly larger sample size, we uncovered several novel loci for cLD, among which our strongest association was rs343029 (Chr 7). This SNP is close to the long non-coding RNA *AC007652.1* (i.e., ENSG00000235464) located between the protein-coding genes *TBX20,* associated with heart-related diseases ^22–26^, and *HERPUD2*, both of which have been linked to electrocardiogram signals ^27^. These coding genes were not detected using PascalX, because they are outside of its default 50kb window around rs343029. Nevertheless, a regulatory effect by *AC007652.1* seems plausible. Furthermore, this SNP has previously been associated with cIMT ^28^. Other new loci uncovered are rs7838131 (Chr 8), which has been associated with body mass index ^29^, systolic blood pressure (SBP) ^29^, hypertension ^29^, as well as coronary artery disease ^30^, and rs111677878 (Chr 11).

In contrast to cLD, the genetic architecture of cIMT has been extensively studied with high-powered GWAS ^28,31,32^. Consistent with prior cIMT research ^28,33^, SNPs rs342988 (Chr 7) and rs1065853 (Chr 19) were also found to be significant in our analysis. These SNPs have further been associated with lipoprotein levels ^34,35^ and myocardial infarction ^36^, emphasising their importance for cardiovascular disease risk. Additionally, rs974819 (Chr 11), which was significantly associated with mean cIMT in this study, has previously been linked to coronary heart disease and was found to exhibit a sex-dependent effect ^37^. To our knowledge, however, several loci identified in this study have not been reported previously in relation to cIMT (including rs374418010, rs146453480, rs559857649, see Supplementary Table 2). Importantly, one of the lead SNPs for cIMT (rs342988) on Chr 7 is in strong linkage disequilibrium with the nearby lead SNP for cLD (rs343029) with R^2^ = 0.77 (see Suppl. Section Genetic Variants) ^38^.

Overall, these results underline the distinct yet interconnected roles of the cLD and cIMT in vascular health, characterised by a relatively high phenotypic correlation (37%) and an even higher genetic correlation (58%). Comparative analyses of our genetic association signals suggest substantial (11 of the 14 genes associated with cIMT were also associated with cLD) but not complete overlap in genetic influences (33 genes were unique to cLD and 3 to cIMT). Furthermore, our study was the first to explore the genetic basis of cIMT/cLD, motivated by the potential biological significance of this ratio in reflecting the relationship between arterial wall thickening and cLD, which may inform about mechanisms such as arterial distensibility and remodelling. SNP rs1065853 (Chr 19) was the only one significantly associated with cIMT/cLD and mean cIMT, while all other significant hits were distinct. Among those were rs625034 (Chr 15), which has previously been associated with thoracic aortic aneurysm ^39^. These findings suggest that the cIMT/cLD captures aspects of vascular health that are not fully represented by either the cLD or cIMT alone. In particular, the lower heritability and weaker genetic signals of the ratio cIMT/cLD, in comparison to the two original phenotypes, may reflect a larger contribution of environmental factors to its variability.

Moreover, our gene-wise analysis highlighted several genes associated with cLD, such as *PCSK7*, associated with blood lipids ^40^, *SIK3*, associated with high-density lipoprotein ^41^, *TAGLN*, associated with triglycerides ^42^, and *APOA1,* associated with blood pressure (BP) ^43^ and lipoproteins ^44^, as well as genes linked to cardiovascular development such as *GATA4* ^45^. Furthermore, genes in the *‘chr8p23’* gene set, implicated in embryonic development ^46–48^, metabolism ^49–51^, and inflammation ^52–54^, shared coherent signals with both cLD and cIMT. These associations emphasise the metabolic and inflammatory pathways underlying vascular health. Interestingly, *ELN*, a gene involved in tissue elasticity and associated with heart diseases ^55–57^, exhibited an anti-coherent association between cLD and cIMT. Some of these genes are also involved in vascular biology, including extracellular-matrix remodelling (e.g., *TAGLN*) ^58^ and regulation of smooth-muscle tone (e.g., *SIK3*) ^59^. This aligns with studies showing that reduced elastin levels result in a narrowed arterial lumen and increased arterial stiffness in both humans and mice ^55,56^.

Our disease association analyses revealed that both cIMT and cLD were strongly associated with established cardiovascular risk factors and prevalent diseases (including BP, arterial stiffness, resting heart rate, pack-years of smoking, and BMI), confirming their relevance as markers of vascular health ^2,3,10^. These associations remained significant after adjusting for age, sex, and height, indicating that these carotid phenotypes capture aspects of vascular function beyond demographic- or size-related effects. The consistently stronger associations we observed for cLD in the disease association analysis suggest that cLD might be more sensitive to systemic stress and be a better biomarker reflecting structural adaptation to risk factors such as chronically elevated BP metabolic load ^8^. In contrast, the unexpected inverse relationship between cIMT and resting heart rate, and cIMT and DBP in medicated participants, may reflect the effects of pharmacological BP control or compensatory arterial remodelling, highlighting distinct physiological roles for these measures, and/or the presence of unmeasured residual confounding factors. In the analysis of binary disease outcomes, both cIMT and cLD were directly associated with prevalent cardiovascular and cerebrovascular disease, hypertension, and diabetes, in line with their known involvement in cardiometabolic risk ^3^. Interestingly, effect sizes and odds ratios were substantially higher for cLD than for cIMT. This could partially be explained by the Glagov phenomenon, where pronounced lumen dilation occurs in individuals with advanced vascular remodelling ^6,7^, i.e., arteries expand to maintain flow as a compensatory response to the wall thickening and stiffening that accompany vascular disease progression. Furthermore, it has been reported in the literature that BP-lowering medications can alter the relationship between the cIMT and cLD increase, with treated hypertensive individuals showing a slower increase in cIMT, but not cLD, than untreated patients (after adjusting for risk factors) ^60^. In general, the shared associations of cIMT and cLD with major categorical risk factors and prevalent disease outcomes reinforce the view that carotid geometry reflects the cumulative impact of systemic risk exposure.

In the survival analysis, higher values of both cIMT and cLD predicted an increased risk of incident cardiovascular events and poorer survival, confirming their prognostic relevance, consistent with previous literature ^10,12,61^. Nevertheless, after full adjustment for major cardiovascular risk factors, only the cIMT/cLD ratio remained significantly associated with cardiovascular outcomes, suggesting that the proportional relationship between arterial wall thickness and lumen diameter retained residual prognostic information not captured by cIMT or cLD alone. Although limited by a smaller number of incident events compared with prevalent ones, this finding indicates that this composite measure could represent an integrated marker of arterial remodelling that remains informative even after controlling for traditional risk factors. Notably, the associations for cIMT and cLD in our group analysis (**Fig. 6** and **Suppl. Fig. 8**) were weaker than those observed in previous literature ^10,12^. This discrepancy can be explained, at least partially, by differences in population characteristics: compared with prior cohorts, our sample from the UK Biobank is younger (54.9 ± 8.9 for the UKB, see **Suppl. Table 9**; 67.4 ± 9.1 for Sedaghat et al.), exhibits a lower burden of cardiometabolic comorbidity (Sedaghat et al. pooled cohort has a substantially higher portion of subjects with diabetes, previous cardiovascular disease, BP lowering medications, and is partially composed by patients diagnosed with chronic kidney disease or on hemodialysis for at least 3 months), and contains substantially fewer events, which may expand confidence intervals and attenuate effect estimates. Furthermore, the reduced cardiovascular risk in our younger and relatively healthier sample may reduce the signal available for cIMT- or cLD- based group stratification, relative to older or higher-risk populations. Additional methodological differences may also contribute to the discrepancy in results, including the set of covariates available for adjustment: in our dataset, blood lipid measurements were not collected at the time of carotid imaging (UKB instance 2), while previous studies adjusted for such risk factors. Finally, the variability in ultrasound acquisition and cLD segmentation protocols, such as the exact location and angle of the common carotid artery imaged and segmented, and the effective segment length used to estimate cLD, may further explain discrepancies in risk estimates across studies. In continuous Cox analyses, the results were more consistent with prior expectations: both cIMT and the cIMT/cLD ratio were significantly associated with incident cardiovascular disease, whereas cIMT and cLD were associated with stroke (**Suppl. Fig. 9a–b**). In sex-stratified Cox models, cIMT remained significantly associated with incident stroke for males, who typically exhibit larger cLD and a higher burden of adverse hemodynamic load, after full adjustment (**Suppl. Fig. 11c**).

Despite its promise, the study has practical limitations that warrant consideration. First, the sizable variability in ultrasound acquisition, such as differences in imaging angle and location along the CCA, might affect the automated cLD segmentation pipeline and thus introduce measurement noise. Second, while the cLD and cIMT were evaluated in a large, population-based cohort, the UKB is not fully representative of the general population, which may limit the generalisability of these findings. Additionally, the prognostic utility of the cLD needs further validation in diverse demographic and clinical populations to establish its role in routine cardiovascular screening. It should also be noted that other research suggests the effect of hypertensive treatments on vascular remodelling can vary substantially based on the class of medications ^62^, and our study did not differentiate between different classes of BP-lowering medication. Such distinctions are not reliably possible in the UKB, where medication information is self-reported, recorded in free-text fields, and often includes brand names, partial names, or non-specific descriptions, which limits the accuracy and completeness of mapping to pharmacological subclasses. Furthermore, we did not adjust for other treatments which have been shown to alter cIMT progression, such as lipid-lowering medications ^63^. Regarding survival analysis, as we mentioned previously, our cohort lacked blood lipid measurements at the imaging visit, which limited covariate adjustment in the Cox models. Moreover, the relatively small number of events for some endpoints may have widened confidence intervals. Taken together, these could act as confounding factors in our disease association and survival analysis, and explain the magnitude and strength of some associations. Finally, the results in our sex-stratified analyses should be interpreted with caution. Although adjustment for age and height reduced some of the expected morphological differences between males and females, the two groups nevertheless differed substantially in their underlying cardiovascular risk profiles, medication use, and, critically, in the number of incident events (see **Suppl. Table 10b**). These imbalances reduce statistical power unevenly across strata and weaken the reliability of estimates, particularly in females, where event counts were lower. As a result, apparent sex-specific differences may partly reflect differential risk-factor distributions and limited power rather than true biological modification of associations. Furthermore, residual confounding by sex-dependent lifestyle or metabolic factors cannot be excluded. Finally, our results have not yet been replicated in an independent cohort, which will be essential to confirm the robustness and reproducibility of the observed associations.

In conclusion, this study explored the genetic architecture of the cLD and showed that it is distinct from the more commonly assessed phenotype of cIMT. By automating the extraction of the cLD from carotid ultrasound images and investigating its genetic determinants, we have established a foundation for incorporating the cLD as a routine measure in cardiovascular screenings. Our findings underline the importance of both cIMT and the cLD as complementary markers, and posit that combining the information on arterial wall thickening and lumen expansion may capture additional information more than either dimension alone. Future studies should carefully assess whether observed associations are genuinely population-wide or predominantly driven by one sex, as differences in baseline risk-factor distributions and vascular behaviour may limit generalisability.

## Methods

### UK Biobank and Carotid Ultrasound Images

The UK Biobank (UKB) is a large-scale biomedical database and research resource containing anonymised genetic, lifestyle, and health information from half a million UK participants. The UKB’s database, which includes blood samples, heart and brain scans, and genetic data of the volunteer participants, is globally accessible to approved researchers who are undertaking health-related research that is in the public interest. UKB recruited 500k people between the ages of 40 and 69 years in 2006-2010 from across the UK. With their consent, they provided detailed information about their lifestyle and physical measures, and had blood, urine, and saliva samples collected and stored for future analysis. It includes multi-organ imaging for many participants, such as magnetic resonance image scans of the brain, heart, and liver, carotid ultrasounds, and retinal colour fundus images ^64^.

We processed 21 838 left and 21 840 right carotid ultrasound DICOM image series, derived from 20 031 and 20 033 subjects of the UKB, respectively, that had been collected to measure cIMT, a marker for subclinical atherosclerosis and cardiovascular disease risk. Images were acquired from both left and right carotid arteries using standardised protocols across all assessment centres. For each artery, images were taken below and near the carotid bifurcation with both a central and lateral viewing orientation (see below for details). The angle of acquisition for each still image did not always align exactly with the target reference angles, resulting in multiple images attempting to capture the same reference angle. For such cases, in our study, we averaged all cLD measures from the same subject and medical visit that passed quality control (QC) to yield a single value per reference angle. We note that only a subset of images captured for each subject showing the CCA are correctly locked on the diastole and feature a small bounding box marking the cIMT. For these images, maximum, mean, and minimum cIMT values are available as part of the UKB. To ensure uniformity in the measurements, we only used these images and corresponding cIMT values for our analyses, even though cIMT values were available for more UKB subjects than those for which we had access to the carotid ultrasound images.

### Lumen Diameter Segmentation

To automate the carotid artery segmentation from ultrasound images, we trained a Deep Learning CNN using N = 79 randomly sampled images labelled by an expert radiologist with more than six years of experience using ITK-SNAP ^65^. During the manual segmentation process, we only selected images showing clear interfaces for blood/intima and media/adventitia. We carefully segmented the anechoic portion of the CCAs, avoiding the inclusion of the intima, to ensure accurate delineation of the lumen. Additionally, to prevent overestimation of cLD, we systematically excluded the carotid bulb. Then, we cropped the labelled ultrasound stills to the image area and converted them to grayscale. For the training, we used the PyTorch-based nnU-Net deep framework ^66^, which implements a heuristic that enables data-driven hyperparameter search. The nnU-Net framework has been shown in numerous studies to be very data-efficient as it only needs small training sample sizes to show substantial generalisability. Given the nature of our images, we limited the training to a 2D patch-based model only. We first trained five different 2D models using nnU-Net’s default 5-fold cross-validation to assess the models’ performance on previously unseen data. After observing a high enough Dice Coefficient on all the folds (Dice Coefficient > 0.95), we trained a model using all the human-labelled data. All the training runs minimised a composite loss of Dice Coefficient and Cross-Entropy and consisted of up to 1000 epochs. Only the best model checkpoint was saved.

Using this model, we segmented all the carotid images in the UKB dataset with an associated cIMT measurement (i.e., correctly locked on diastole, as explained in the previous section). Since such information was annotated in the corner of each image, we used an out-of-the-box optical character recognition model (PyTesseract).

### Lumen Median Diameter Measurement Per Image

The QC began with evaluating the number of segmented objects in each image. If more than one object was identified, we compared their areas and excluded objects with an area smaller than half the maximum object area. Furthermore, we discarded images if more than one object remained after this step (**Suppl. Fig. 1a**). Next, we assessed the shape and regularity of the remaining segmented object. Segmented objects were required to resemble rectangles or squares and to have fewer than a defined number of contour points. We set this threshold to 215 based on an initial analysis of a subset of our dataset. While a perfect rectangle or square would have four points, minor pixel-level irregularities justified a higher threshold (**Suppl. Fig. 1b**).

After QC, we measured the median cLD for each image in pixels using the segmented cLD (described in the previous section). For that purpose, we computed the centre of gravity of the segmentation using the central moments (**Equation 1**) along with the major axis angle (Θ) (**Equation 2**). To ensure consistent alignment of the major axis with the cLD, the angle was adjusted when |Θ| exceeded π/4 by applying the transformation Θ = |Θ| - π/2.

We then defined transverse lines across the length of the major axis at intervals of two pixels. For each line, we measured the distance between the upper and lower segmented boundaries (**Suppl. Fig. 2**; green lines in **Fig. 1b**). To mitigate irregularities, we excluded lines with distances less than half the median line length. The median of the remaining line distances was calculated to represent the median cLD for the image.

This approach assumes that defining the cLD linearly is a reasonable approximation for this dataset. If this assumption does not hold for other datasets, alternative methods incorporating higher-order approximations may be required.

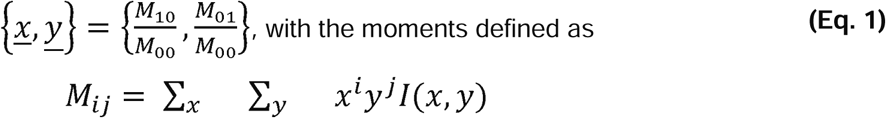

and *l*(*x,y*) being the image pixel intensities.

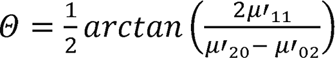, with the central moments defined as

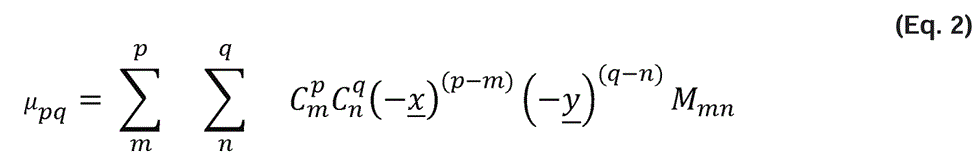

### Lumen Diameter Phenotypes Measurement Per Subject

After obtaining the median cLD for each image, we calculated cLD phenotypes for each participant. At least four carotid images were measured per participant during the same session, corresponding to four reference angles ( =120°, 150°, 210°, and 240°), corresponding to right lateral cLD, right central cLD, left central cLD, and left lateral cLD, respectively. (see **Fig. 1a**).

In some cases, multiple images were taken for the same reference angle for a single participant. Our analysis revealed that measurements for images targeting the same angle were nearly identical, with only minimal variations. Therefore, for participants with multiple images per angle, we averaged the median cLD values to generate a single value per angle. Sometimes there were multiple instances per subject; however, we only included the first instance, since not many subjects would have been added by using different instances (**Suppl. Table 6**; **Suppl. Fig. 5**).

### Derived Lumen Diameter Phenotypes Definition

We derived five additional cLD phenotypes:

- Right cLD = (right central cLD + right lateral cLD) / 2
- Left cLD = (left central cLD + left lateral cLD) / 2
- Central cLD = (right central cLD + left central cLD) / 2
- Lateral cLD = (right lateral cLD + left lateral cLD) / 2
- Mean cLD = (right central cLD + right lateral cLD + left central cLD + left lateral cLD) / 4

In case some of the angle-specific primary cLD measurements were missing, we computed these derived phenotypes using the available data in order to maximise the number of subjects with these cLD phenotypes.

### Carotid Intima Media Thickness

The carotid ultrasound images displayed data on the mean, minimum, and maximum cIMT. Using an optical character recognition model, we extracted this information from each image. We then applied the same preprocessing pipeline as we had used previously for the cLD measurements to calculate the mean cIMT across the four reference angles. This resulted in a single measure of mean cIMT for each participant.

To examine the relationship between cIMT and cLD, we derived a normalised phenotype as the ratio of mean cIMT over mean cLD. The distributions of all phenotypes can be found in the **Suppl. Fig. 3**, and cohort characteristics can be found in **Suppl. Table 4**.

The resulting dataset comprised nine cLD phenotypes per participant, along with the mean cIMT and the cIMT/cLD ratio.

### Genome-wide association analysis (GWAS)

We performed the GWAS for all phenotypes using regenie ^67^. Prior to the analysis, the genotype data underwent QC as recommended for UKB genotype data ^68^ using PLINK2 ^69^ (MAF = 0.01, MAC = 100, SNP genotype missingness = 0.1, individual genotype missingness = 0.1, HWE = 1E-15). We rank-inverse normal transformed all the phenotypes prior to analysis, and included in the GWAS sex, age, age-squared, assessment centre, standing height, and the first 20 genetic principal components (PCs) as covariates.

### Genetic Correlations and SNP-Heritabilities

We used summary statistics as input to compute the genetic correlations and h², which were computed using LDSR ^15,16^.

### Genes and Pathways

We computed gene and pathway scores using PascalX ^17,18^. We scored both protein-coding genes and lincRNAs using the approximate “saddle” method, taking into account all SNPs with a minor allele frequency > 0.05 within a 50 kb window around each gene. We scored all pathways available in MSigDB v7.2 using PascalX’s ranking mode, fusing and rescoring any co-occurring genes less than 100kb apart. PascalX requires linkage disequilibrium structure to accurately compute gene scores, which in our analyses was provided with the UK10K (hg19) reference panel. We corrected for bias due to sample overlap using the intercept from pairwise LDSR genetic correlation. We set the significance threshold at 5.7 for genes and 5.8 pathways, i.e., 0.05 divided by the number of tested genes/pathways.

### Disease Association and Survival Analysis

In our association analyses, we included major continuous systemic risk factors, known to influence disease susceptibility, acquired at the time of the first carotid imaging (UKB’s instance two). Furthermore, we incorporated additional disease and risk factor information with binary (or categorical) outcomes, favouring objectively measured sources, including health-related records (Category 100091) and physical measurements (Category 100006), over self-reported information collected through the assessment centre Touchscreen questionnaire (Category 100025) or verbal interviews (Category 100071). We prioritised these sources to minimise the impact of reporting errors and inconsistencies commonly found in self-reported data ^70,71^, with the only exception being smoking status (i.e., never, previous, current). Given the high prevalence of undiagnosed diabetes, prediabetes, and related complications reported in previous studies ^72,73^, we determined diabetes status using a combination of clinical and biochemical criteria. We classified participants as diabetic if they had an official diagnosis recorded as ICD code E10 (insulin-dependent diabetes mellitus), glycated haemoglobin (HbA1c) levels ≥48 mmol/mol, or glucose levels ≥11.1 mmol/L (using blood biochemistry data from previous visits) ^74,75^. Instead of adjusting the automated blood pressure readings taken at the assessment centre visits to account for the effect of blood pressure-lowering medications, we decided to split the cohort into un-medicated and medicated (for hypertension) subjects. In our analysis of binary disease outcomes, we defined hypertension as having SBP > 140 or DBP > 90 or the use of blood pressure-lowering medication. We used multiple linear regression for continuous risk factors and multiple logistic regression for binary ones. All models were corrected for sex, age, and standing height. We standardised all the independent variables used for the linear regressions and binarised all diseases collected as “date first diagnosed” for the logistic regressions. To determine the significance of regression coefficients, we computed p-values and corrected for multiple hypothesis testing using the Benjamini-Hochberg method ^76^, using a false discovery rate of 0.05. All the fields used for the analysis, along with their descriptions, can be found in **Suppl. Tables 7–8**. Moreover, we reported the number of prevalent events in the disease analysis cohort **Suppl. Table 10a**. Finally, prompted by recent research highlighting sex-specific differences in vascular phenotypes ^77^, we conducted a sex-stratified analysis to investigate whether effect sizes differed in males and females. We documented the number of prevalent events, stratified by sex, in **Suppl. Table 10b**.

For survival analysis, we defined two distinct clinical endpoints: (1) cardiovascular events, derived by aggregating ischaemic heart disease (IHD), myocardial infarction (MI), stroke, non-ischaemic cardiomyopathies (NIC), and thrombotic events, and (2) all-cause mortality. Time to event (TTE) was defined as the number of days between the date of carotid ultrasound imaging (instance two) and the date of the recorded event. For participants who experienced multiple events, we used the first event occurring after the imaging visit. We fitted Cox proportional hazards models with two levels of covariate adjustment to assess the sensitivity of the different carotid phenotypes to demographics and major cardiovascular risk factors. First, we adjusted for age, sex, and height, as these are key determinants of cLD and drive cIMT as well. Second, we additionally adjusted for cardiovascular risk factors consistent with those used in the Framingham risk score [62], including age, sex, systolic blood pressure (SBP), BMI, smoking status, diabetes diagnosis, and hypertension treatment, as well as whether the participant had a previous cardiovascular event. We defined hypertension treatment by aggregating male- and female-specific medication data and filtering for “blood pressure medication.” We included prior cardiovascular events as a covariate to account for the increased risk of recurrence [63]. We were unable to adjust for HDL and total cholesterol since these measurements were not taken for subjects imaged at instance 2. For a detailed breakdown of the fields used to define risk factors, events, and endpoints, see **Suppl. Tables 7–8**. Furthermore, we reported the baseline characteristics for the survival analysis cohort in **Suppl. Table 9**, and the number of incident events (for the different levels of adjustment described above) in **Suppl. Table 10a**. Similarly to our association analysis, we conducted a sex-stratified analysis to investigate whether HR differed in males and females. We noted the number of incident events, stratified by sex, before and after excluding incomplete observations, in **Suppl. Table 10b**.

## Data and Code Availability

GWAS summary statistics will be available on Zenodo after the peer-reviewed publication. Image-derived phenotypic data are under restricted access and will only be available through the UKB cohort platform (https://www.ukbiobank.ac.uk/) after peer-reviewed publication. The raw UKB data are protected and not open access; however, they can be obtained upon project creation and acceptance. The code will be available on GitHub after the peer-reviewed publication of the manuscript.

## Data Availability

GWAS summary statistics will be available on Zenodo after the peer-reviewed publication.
Image-derived phenotypic data is under restricted access and will only be available through
the UKB cohort platform (https://www.ukbiobank.ac.uk/) after peer-reviewed publication. The
raw UKB data are protected and not open access; however, they can be obtained upon
project creation and acceptance. The code will be available on GitHub after the
peer-reviewed publication of the manuscript.

https://docs.google.com/spreadsheets/d/1aRe5bXPm591-0RDubQ2HCm00jrY39a4l5nxCL_lXmF8/edit?gid=539079828#gid=539079828

## Acknowledgements

The authors thank Dr. Tamar Itach for her feedback on the manuscript and are grateful to the study participants and the staff from the UKB.

## Author contributions

SB and SOV conceptualised and designed the study. BM and SOV worked on segmenting the ground truth images. SOV performed phenotype extraction and quality control. SOV and LB performed GWAS analyses collaboratively. SOV estimated heritability, cross-phenotype correlations, and conducted gene and pathway analyses. OT analysed cross-phenotype gene associations using PascalX’s coherence scores. DB performed image preprocessing, trained the image segmentation pipeline, and conducted the survival analyses. IQ, DB and DP conducted the disease association analysis. The manuscript was written by DB, SOV, and SB, with contributions from all other authors.

## References

1. Pignoli, P., Tremoli, E., Poli, A., Oreste, P. & Paoletti, R. Intimal plus medial thickness of the arterial wall: a direct measurement with ultrasound imaging. Circulation 74, 1399– 1406 (1986).

2. Baldassarre, D., Amato, M., Bondioli, A., Sirtori, C. R. & Tremoli, E. Carotid artery intima-media thickness measured by ultrasonography in normal clinical practice correlates well with atherosclerosis risk factors. Stroke 31, 2426–2430 (2000).

3. van den Oord SC, Sijbrands EJ, ten Kate GL, van Klaveren D, van Domburg RT, van der Steen AF, Schinkel AF. Carotid intima-media thickness for cardiovascular risk assessment: systematic review and meta-analysis. Atherosclerosis 228, 1–11 (2013).

4. Omarov, M. et al. Automated Deep Learning-Based Detection of Early Atherosclerotic Plaques in Carotid Ultrasound Imaging. medRxiv 2024.10.17.24315675 (2025) doi:10.1101/2024.10.17.24315675.

5. Howard, D. P. J., Gaziano, L., Rothwell, P. M. & Oxford Vascular Study. Risk of stroke in relation to degree of asymptomatic carotid stenosis: a population-based cohort study, systematic review, and meta-analysis. Lancet Neurol 20, 193–202 (2021).

6. Korshunov, V. A., Schwartz, S. M. & Berk, B. C. Vascular Remodeling. Arteriosclerosis, Thrombosis, and Vascular Biology (2007) doi:10.1161/ATVBAHA.106.129254.

7. Watase, H. et al. Carotid artery remodeling is segment specific: An in vivo study by vessel wall magnetic resonance imaging: An in vivo study by vessel wall magnetic resonance imaging. Arterioscler. Thromb. Vasc. Biol. 38, 927–934 (2018).

8. Kozakova, M. et al. Obesity and carotid artery remodeling. Nutrition & Diabetes 5, e177–e177 (2015).

9. Morbid obesity is associated with hypertrophic outward remodeling and increased stiffness of small conduit arteries: An ultra-high frequency ultrasound study. Nutrition, Metabolism and Cardiovascular Diseases 33, 408–415 (2023).

10. Fritze, F. et al. Carotid Lumen Diameter Is Associated With All-Cause Mortality in the General Population. J Am Heart Assoc 9, e015630 (2020).

11. Eigenbrodt, M. L. et al. Common carotid artery wall thickness and external diameter as predictors of prevalent and incident cardiac events in a large population study. Cardiovasc. Ultrasound 5, 11 (2007).

12. Sedaghat, S. et al. Common carotid artery diameter and risk of cardiovascular events and mortality: Pooled analyses of four cohort studies: Pooled analyses of four cohort studies. Hypertension 72, 85–92 (2018).

13. Bella, J. N. et al. Genome-wide linkage analysis of carotid artery lumen diameter: the strong heart family study. Int. J. Cardiol. 168, 3902–3908 (2013).

14. Proust, C. et al. Contribution of Rare and Common Genetic Variants to Plasma Lipid Levels and Carotid Stiffness and Geometry: A Substudy of the Paris Prospective Study 3. Circ Cardiovasc Genet 8, 628–636 (2015).

15. Bulik-Sullivan, B. K. et al. LD Score regression distinguishes confounding from polygenicity in genome-wide association studies. Nat. Genet. 47, 291–295 (2015).

16. Bulik-Sullivan, B. et al. An atlas of genetic correlations across human diseases and traits. Nat. Genet. 47, 1236–1241 (2015).

17. Lamparter, D., Marbach, D., Rueedi, R., Kutalik, Z. & Bergmann, S. Fast and rigorous computation of gene and pathway scores from SNP-based summary statistics. PLoS Comput. Biol. 12, e1004714 (2016).

18. Krefl, D. & Bergmann, S. Cross-GWAS coherence test at the gene and pathway level. PLoS Comput. Biol. 18, e1010517 (2022).

19. Manbachi, A., Hoi, Y., Wasserman, B. A., Lakatta, E. G. & Steinman, D. A. On the shape of the common carotid artery with implications for blood velocity profiles. Physiol Meas 32, 1885–1897 (2011).

20. Han, N. et al. The hemodynamic and geometric characteristics of carotid artery atherosclerotic plaque formation. Quant Imaging Med Surg 14, 4348–4361 (2024).

21. Selwaness, M. et al. Atherosclerotic Plaque in the Left Carotid Artery Is More Vulnerable Than in the Right. Stroke (2014) doi:10.1161/STROKEAHA.114.005202.

22. Luyckx, I. et al. Copy number variation analysis in bicuspid aortic valve-related aortopathy identifies TBX20 as a contributing gene. Eur J Hum Genet 27, 1033–1043 (2019).

23. Yang, X. F. et al. Relationship between TBX20 gene polymorphism and congenital heart disease. Genet Mol Res 15, (2016).

24. Zhou, Y.-M. et al. A novel TBX20 loss of function mutation contributes to adult onset dilated cardiomyopathy or congenital atrial septal defect. Mol Med Rep 14, 3307–3314 (2016).

25. Amor-Salamanca, A., et al. Role of Truncating Variants in Dilated Cardiomyopathy and Left Ventricular Noncompaction. Circ Genom Precis Med 17, e004404 (2024).

26. Zhao, C.-M. et al. TBX20 loss-of-function mutation associated with familial dilated cardiomyopathy. Clin Chem Lab Med 54, 325–332 (2016).

27. Scholman, K. T., Meijborg, V. M. F., Gálvez-Montón, C., Lodder, E. M. & Boukens, B. J. From Genome-Wide Association Studies to Cardiac Electrophysiology: Through the Maze of Biological Complexity. Front Physiol 11, 557 (2020).

28. Yeung, M. W. et al. Twenty-five novel loci for carotid intima-media thickness: A genome-wide association study in >45 000 individuals and meta-analysis of >100 000 individuals. Arterioscler. Thromb. Vasc. Biol. 42, 484–501 (2022).

29. Christakoudi, S., Evangelou, E., Riboli, E. & Tsilidis, K. K. GWAS of allometric body-shape indices in UK Biobank identifies loci suggesting associations with morphogenesis, organogenesis, adrenal cell renewal and cancer. Sci. Rep. 11, 10688 (2021).

30. Gill, D. et al. Genetically predicted midlife blood pressure and coronary artery disease risk: Mendelian randomization analysis. J. Am. Heart Assoc. 9, e016773 (2020).

31. Meena, D. et al. Genome-wide association study and multi-ancestry meta-analysis identify common variants associated with carotid artery intima-media thickness. medRxiv 2025.04.11.25325582 (2025) doi:10.1101/2025.04.11.25325582.

32. Starčević, J. N. & Petrovič, D. Carotid intima media-thickness and genes involved in lipid metabolism in diabetic patients using statins--a pathway toward personalized medicine. Cardiovasc Hematol Agents Med Chem 11, 3–8 (2013).

33. Strawbridge, R. J. et al. Carotid intima-media thickness: Novel loci, sex-specific effects, and genetic correlations with obesity and glucometabolic traits in UK Biobank. Arterioscler. Thromb. Vasc. Biol. 40, 446–461 (2020).

34. Surakka, I. et al. The impact of low-frequency and rare variants on lipid levels. Nat. Genet. 47, 589–597 (2015).

35. Richardson, T. G. et al. Characterising metabolomic signatures of lipid-modifying therapies through drug target mendelian randomisation. PLoS Biol. 20, e3001547 (2022).

36. Hartiala, J. A. et al. Genome-wide analysis identifies novel susceptibility loci for myocardial infarction. Eur. Heart J. 42, 919–933 (2021).

37. Zhou, J. et al. A case-control study provides evidence of association for a common SNP rs974819 in PDGFD to coronary heart disease and suggests a sex-dependent effect. Thromb. Res. 130, 602–606 (2012).

38. NCI, CBIIT, DCEG & Machiela. LDlink. https://ldlink.nih.gov/?var1=rs343029&var2=rs7792074&pop=GBR&genome_build=grch37&tab=ldpair.

39. Ashvetiya, T. et al. Analysis of UK Biobank Cohort Reveals Novel Insights for Thoracic and Abdominal Aortic Aneurysms. Genomics (2021).

40. GeneCards Human Gene Database. PCSK7 Gene - GeneCards. https://www.genecards.org/cgi-bin/carddisp.pl?gene=PCSK7&keywords=PCSK7.

41. GeneCards Human Gene Database. SIK3 Gene - GeneCards. https://www.genecards.org/cgi-bin/carddisp.pl?gene=SIK3&keywords=SIK3.

42. GeneCards Human Gene Database. TAGLN Gene - GeneCards. https://www.genecards.org/cgi-bin/carddisp.pl?gene=TAGLN&keywords=TAGLN.

43. Hoffmann, T. J. et al. Genome-wide association analyses using electronic health records identify new loci influencing blood pressure variation. Nat. Genet. 49, 54–64 (2017).

44. Sinnott-Armstrong, N. et al. Genetics of 35 blood and urine biomarkers in the UK Biobank. Nat. Genet. 53, 185–194 (2021).

45. The Molecular Mechanisms of Cardiac Development and Related Diseases Yingrui Li.

46. Simpson, N. H. et al. Genome-wide analysis identifies a role for common copy number variants in specific language impairment. Eur. J. Hum. Genet. 23, 1370–1377 (2015).

47. SOX7 expression is critically required in FLK1-expressing cells for vasculogenesis and angiogenesis during mouse embryonic development. Mechanisms of Development 146, 31–41 (2017).

48. Afouda, B. A. Towards Understanding the Gene-Specific Roles of GATA Factors in Heart Development: Does GATA4 Lead the Way? International Journal of Molecular Sciences 23, 5255 (2022).

49. Manning, A. K. et al. A long non-coding RNA, LOC157273, is an effector transcript at the chromosome 8p23.1-PPP1R3B metabolic traits and type 2 diabetes risk locus. Front. Genet. 11, 615 (2020).

50. The MTMR9 rs2293855 polymorphism is associated with glucose tolerance, insulin secretion, insulin sensitivity and increased risk of prediabetes. Gene 546, 150–155 (2014).

51. Secolin, R. et al. Exploring a Region on Chromosome 8p23.1 Displaying Positive Selection Signals in Brazilian Admixed Populations: Additional Insights Into Predisposition to Obesity and Related Disorders. Front. Genet. 12, 636542 (2021).

52. Guo, Y. et al. Genetic association of inflammatory marker GlycA with lung function and respiratory diseases. Nat. Commun. 15, 3751 (2024).

53. Zhong, J. et al. MFHAS1 Is Associated with Sepsis and Stimulates TLR2/NF-κB Signaling Pathway Following Negative Regulation. PLOS ONE 10, e0143662 (2015).

54. Thomas, M. F. et al. Eri1 regulates microRNA homeostasis and mouse lymphocyte development and antiviral function. Blood 120, (2012).

55. Cocciolone, A. J. et al. Elastin, arterial mechanics, and cardiovascular disease. American Journal of Physiology-Heart and Circulatory Physiology (2018) doi:10.1152/ajpheart.00087.2018.

56. Wahart, A. et al. Role of elastin peptides and elastin receptor complex in metabolic and cardiovascular diseases. The FEBS Journal 286, 2980–2993 (2019).

57. Francis, C. M. et al. Genome-wide associations of aortic distensibility suggest causality for aortic aneurysms and brain white matter hyperintensities. Nature Communications 13, 1–18 (2022).

58. The canonical smooth muscle cell marker TAGLN is present in endothelial cells and is involved in angiogenesis Kiyomi Tsuji-Tamura 1. Saori Morino-Koga 1,.

59. Kinase 3 Promotes Vascular Smooth Muscle Cell Proliferation and Arterial Restenosis by Regulating AKT and PKA-CREB Signaling Yujun Cai 1 2, Xue-Lin Wang 2. Jonathan Dong 2,.

60. Chironi, G. et al. Influence of Hypertension on Early Carotid Artery Remodeling. *Arteriosclerosis*, Thrombosis, and Vascular Biology (2003) doi:10.1161/01.ATV.0000083342.98342.22.

61. Carotid intima-media thickness, cardiovascular disease, and risk factors in 29,000 UK Biobank adults. American Journal of Preventive Cardiology 22, 101011 (2025).

62. American Heart Association Journals. American Heart Association Journals 10.1161/01.STR.0000227223.90239.13.

63. Huang, R. et al. Comparative effects of lipid lowering, hypoglycemic, antihypertensive and antiplatelet medications on carotid artery intima-media thickness progression: a network meta-analysis. Cardiovascular Diabetology 18, 1–10 (2019).

64. Littlejohns, T. J. et al. The UK Biobank imaging enhancement of 100,000 participants: rationale, data collection, management and future directions. Nat. Commun. 11, 2624 (2020).

65. 3D Active Contour Segmentation of Anatomical Structures: Significantly Improved Efficiency and Reliability.

66. Isensee, F., Jaeger, P. F., Kohl, S. A. A., Petersen, J. & Maier-Hein, K. H. nnU-Net: a self-configuring method for deep learning-based biomedical image segmentation. Nat. Methods 18, 203–211 (2021).

67. Mbatchou, J. et al. Computationally efficient whole-genome regression for quantitative and binary traits. Nat. Genet. 53, 1097–1103 (2021).

68. UKBB Analysis - regenie. https://rgcgithub.github.io/regenie/recommendations/.

69. Chang, C. C. et al. Second-generation PLINK: rising to the challenge of larger and richer datasets. Gigascience 4, 7 (2015).

70. Modin, H. E. et al. Pack-Year Cigarette Smoking History for Determination of Lung Cancer Screening Eligibility. Comparison of the Electronic Medical Record versus a Shared Decision-making Conversation. Annals of the American Thoracic Society (2017) doi:10.1513/AnnalsATS.201612-984OC.

71. Schoeler, T., Pingault, J.-B. & Kutalik, Z. The impact of self-report inaccuracy in the UK Biobank and its interplay with selective participation. Nat Hum Behav (2024) doi:10.1038/s41562-024-02061-w.

72. Dankner, R., Geulayov, G., Olmer, L. & Kaplan, G. Undetected type 2 diabetes in older adults. Age Ageing 38, 56–62 (2009).

73. Pierce, M. B., Zaninotto, P., Steel, N. & Mindell, J. Undiagnosed diabetes—data from the English longitudinal study of ageing. Diabetic Medicine 26, 679–685 (2009).

74. Qiao, Z. et al. Estimation and implications of the genetic architecture of fasting and non-fasting blood glucose. Nat Commun 14, 451 (2023).

75. Lugner, M., Rawshani, A., Helleryd, E. & Eliasson, B. Identifying top ten predictors of type 2 diabetes through machine learning analysis of UK Biobank data. Sci Rep 14, 2102 (2024).

76. Benjamini, Y. & Hochberg, Y. Controlling the False Discovery Rate: A Practical and Powerful Approach to Multiple Testing. J. R. Stat. Soc. Series B Stat. Methodol. 57, 289–300 (1995).

77. Böttger, L. et al. Sex-specific disease association and genetic architecture of retinal vascular traits. BioRxiv.

