## Supplementary Figures for "Genetic and Etiological Insights from Automated Lumen Diameter Measurements in Carotid Ultrasounds of the UK Biobank"

**a.**

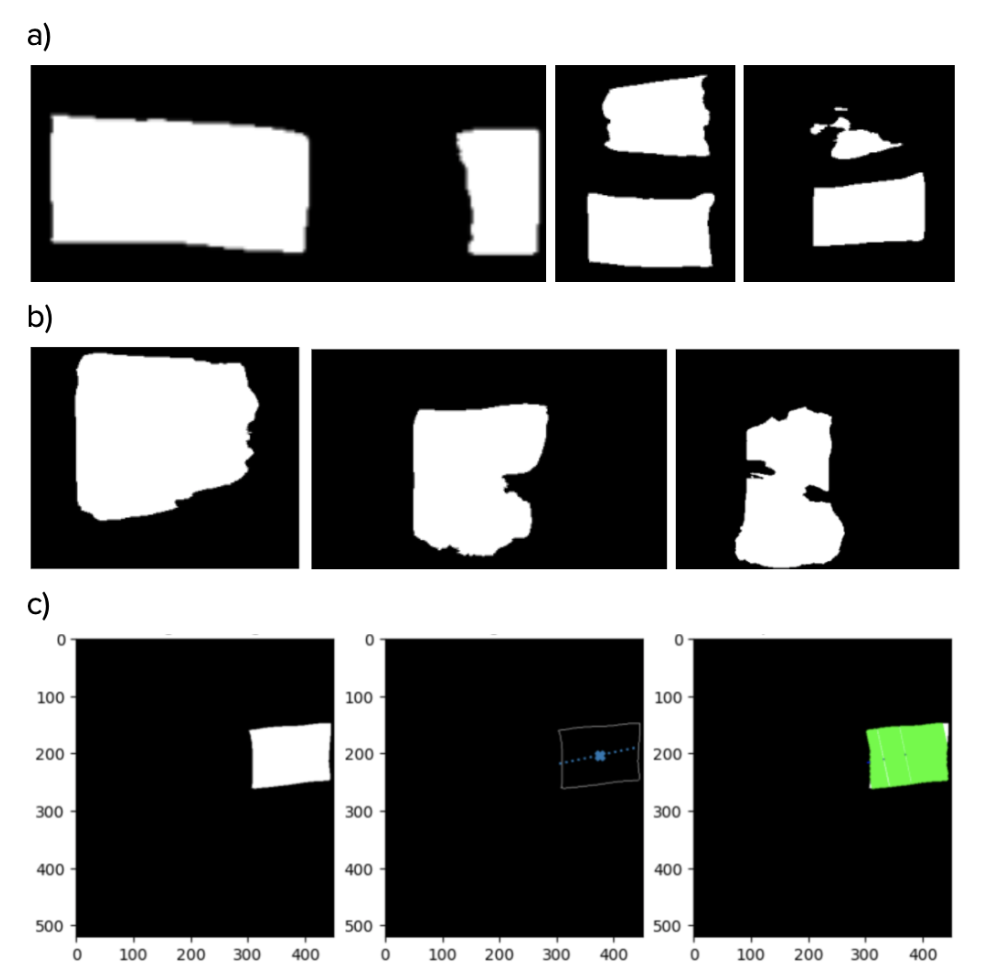

**b.**

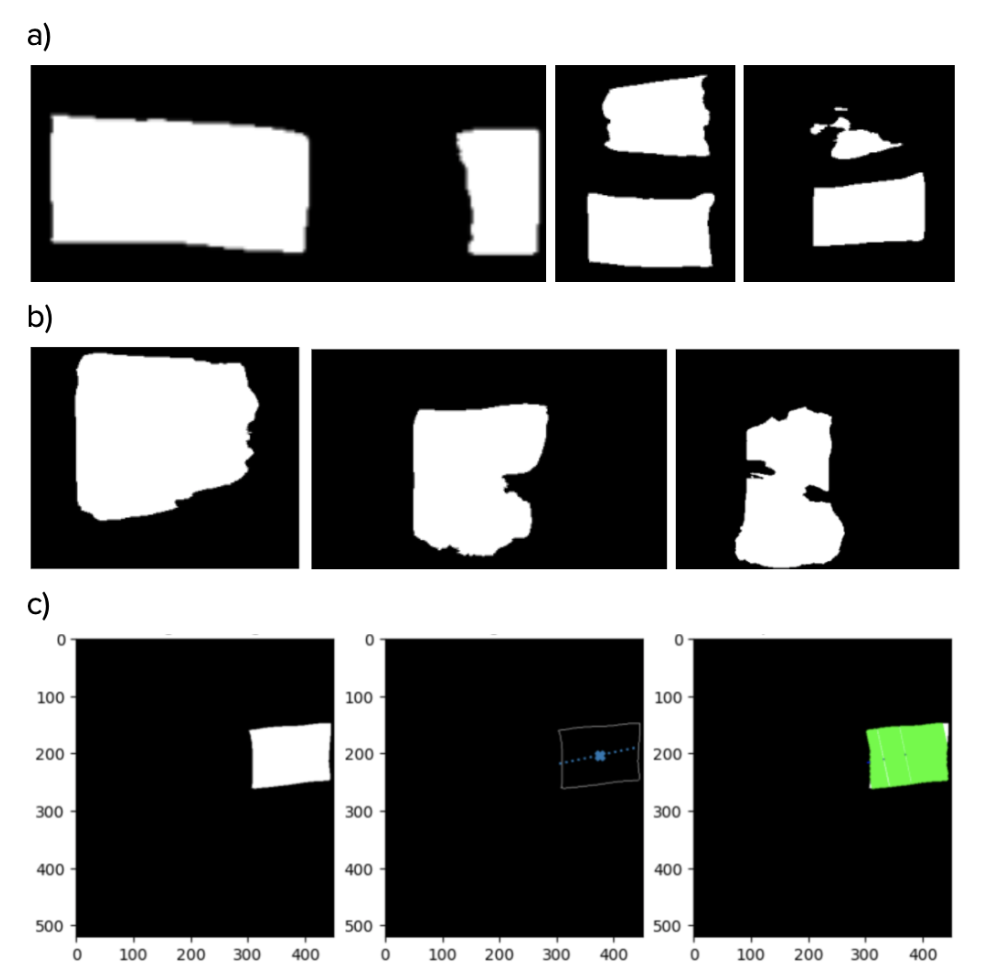

**Supplementary Figure 1 | QC Process for Segmented Lumen Objects.** **a)** Example segmentations evaluated based on the number of segmented objects per image. When multiple objects were detected, their areas were compared, and objects with areas smaller than half the maximum object area were excluded. Images without any qualifying objects were discarded (like the image in the centre). **b)** Examples of the shape and regularity assessment of segmented objects. Objects were required to resemble rectangles or squares and have fewer than 215 contour points to pass QC (none of these images passed this criterion).

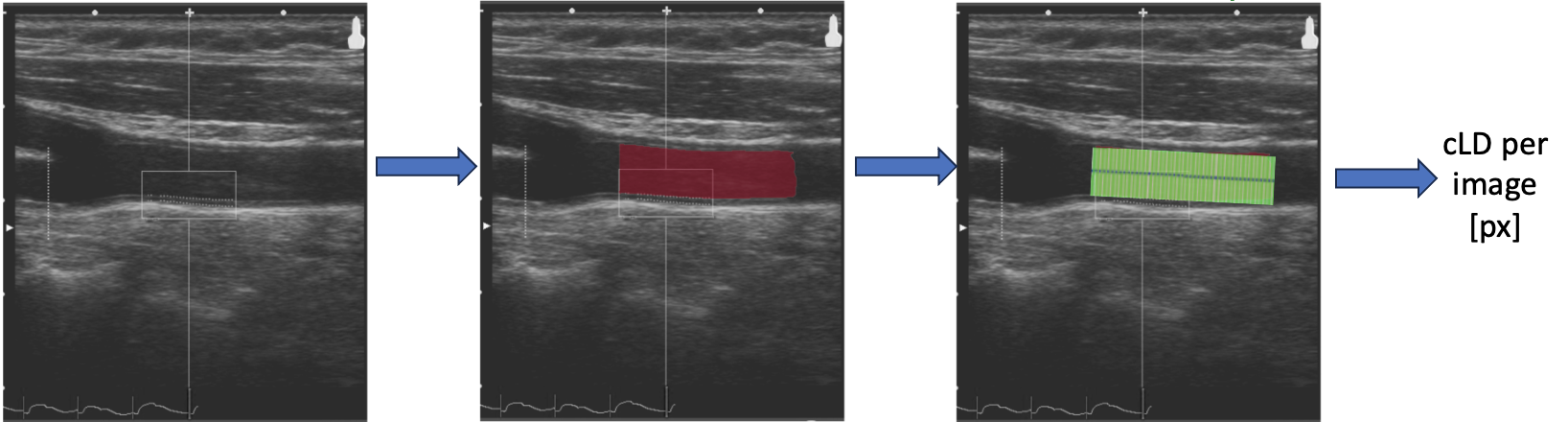

**Supplementary Figure 2 | cLD Measurement.** Measurement of the lumen diameter on a sample image from the UK Biobank.

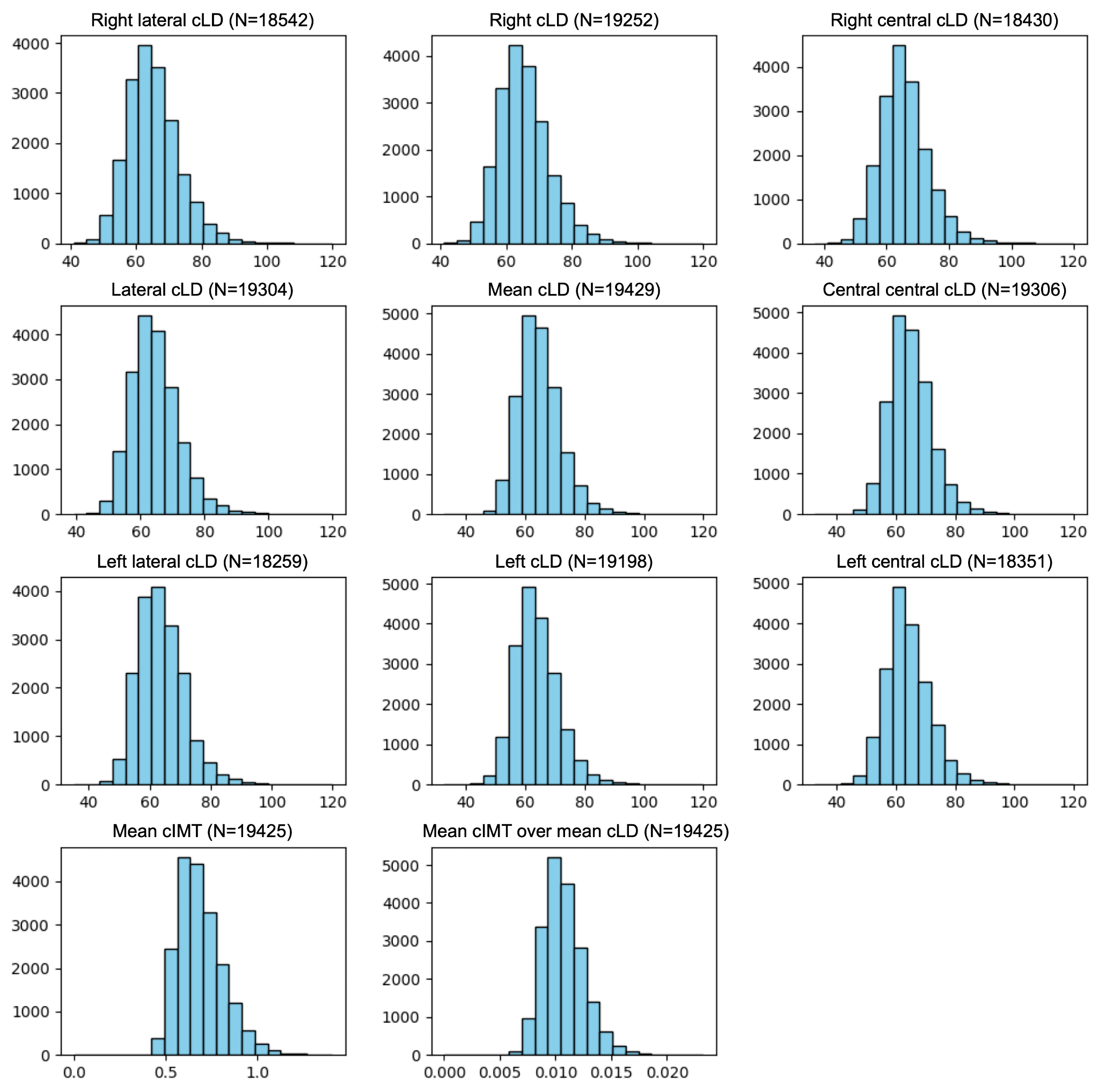

**Supplementary Figure 3 | Distributions of cLD and cIMT (subject Level).** Distributions of cLD measurements and derived phenotypes, for instance 2, following the exclusion of outliers exceeding 10 times the standard deviation of the respective phenotype. Initial measurements were taken at four angles (120º, right lateral cLD; 150º, right central cLD; 210º, left central cLD; and 240º, left lateral cLD), with additional phenotypes derived as follows: right cLD is the mean of the right central and lateral cLDs; left cLD is the mean of the left central and lateral cLDs; lateral cLD is the mean of the left and right lateral measurements; central cLD is the mean of the left and right central measurements; and mean cLD is the mean of all the four cLD measurements. The figure also includes the distributions of the mean carotid intima-media thickness, or mean cIMT, and the ratio of mean cIMT to mean cLD.

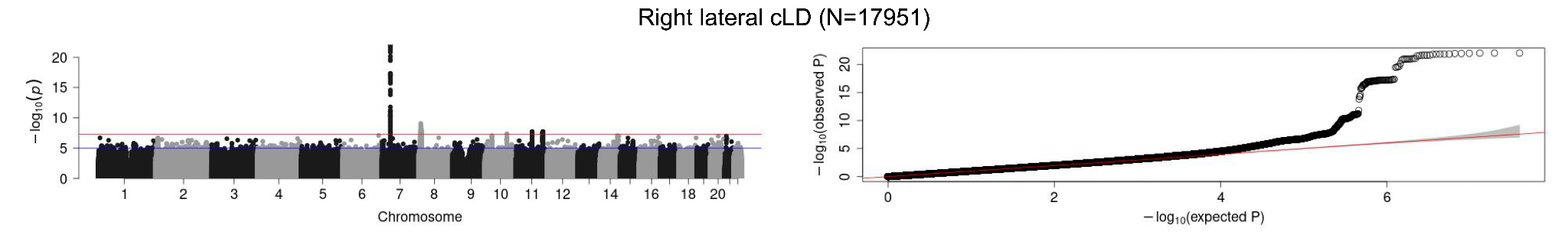

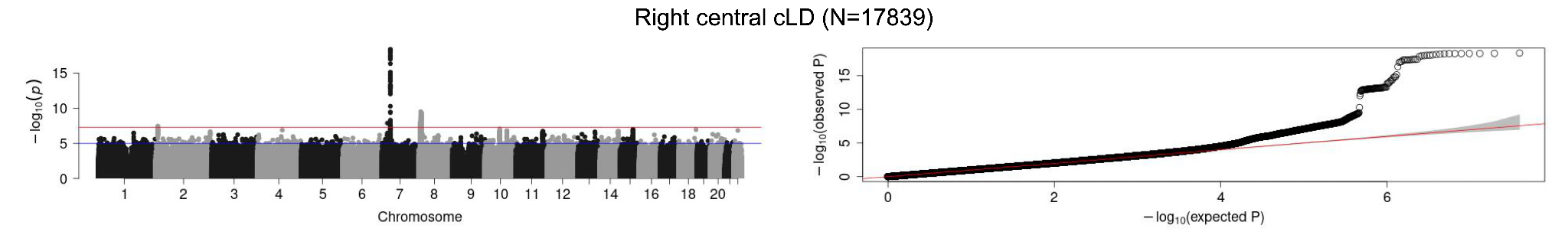

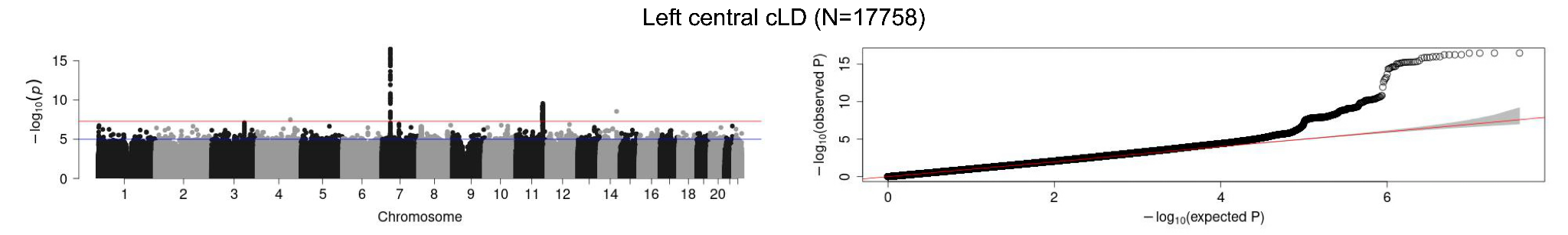

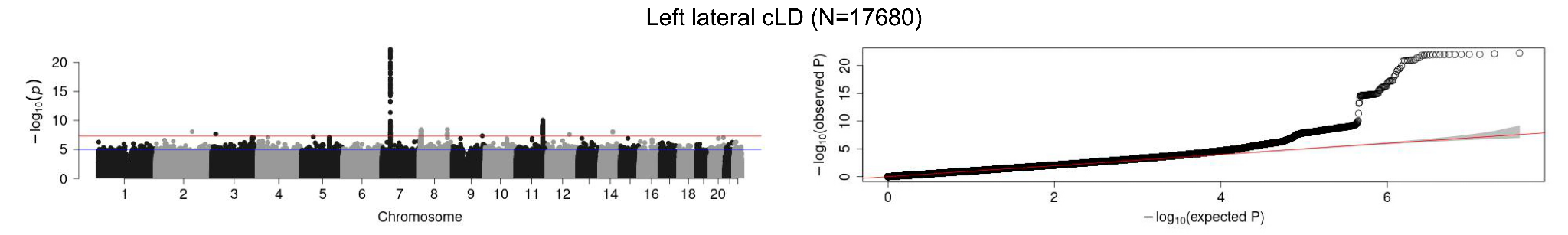

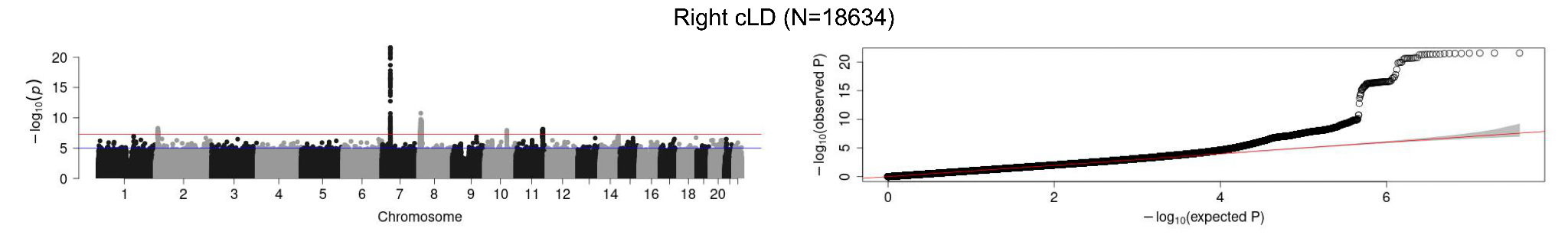

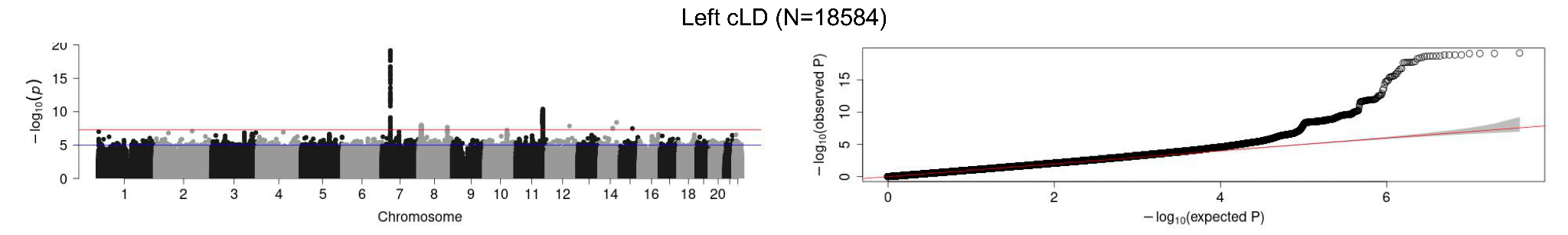

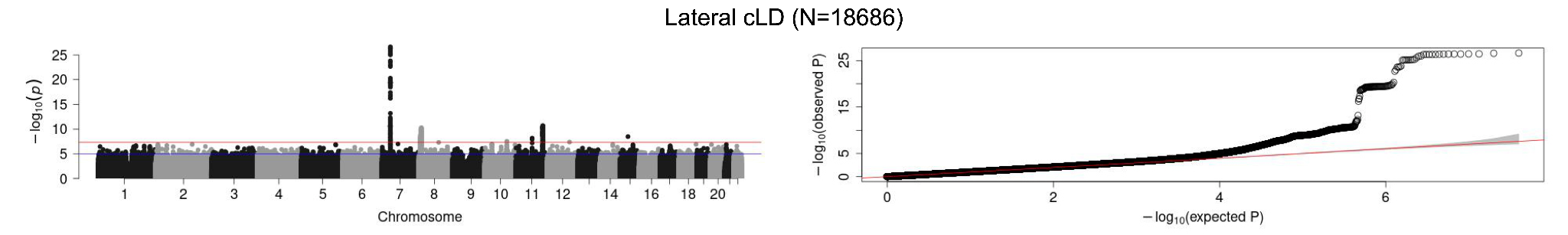

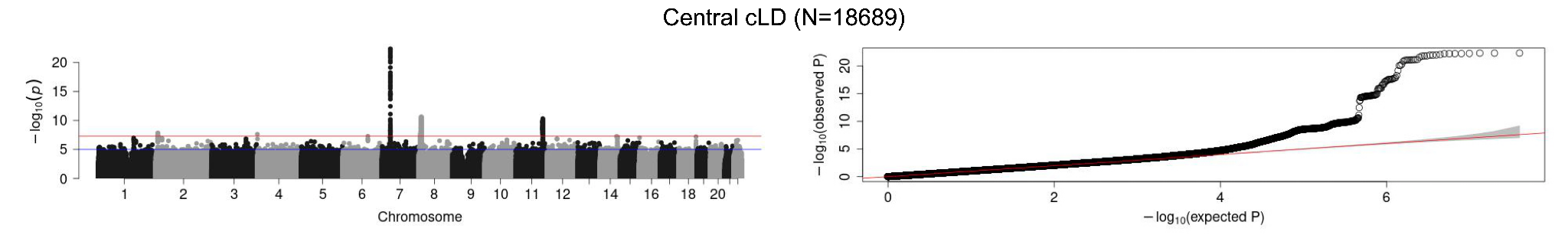

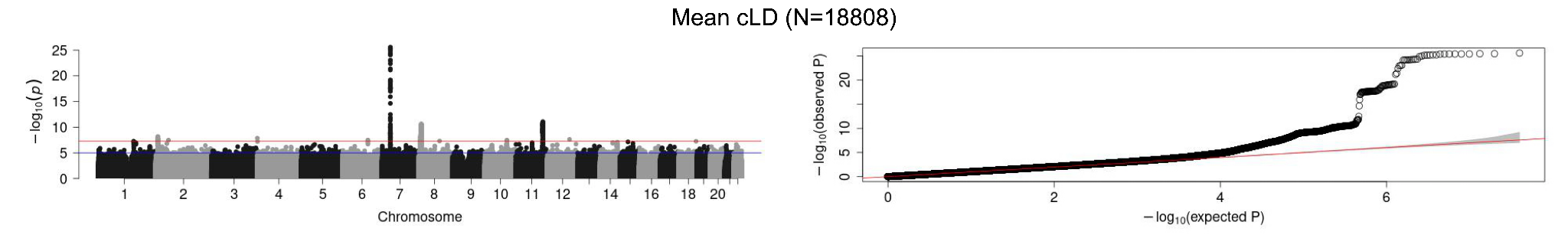

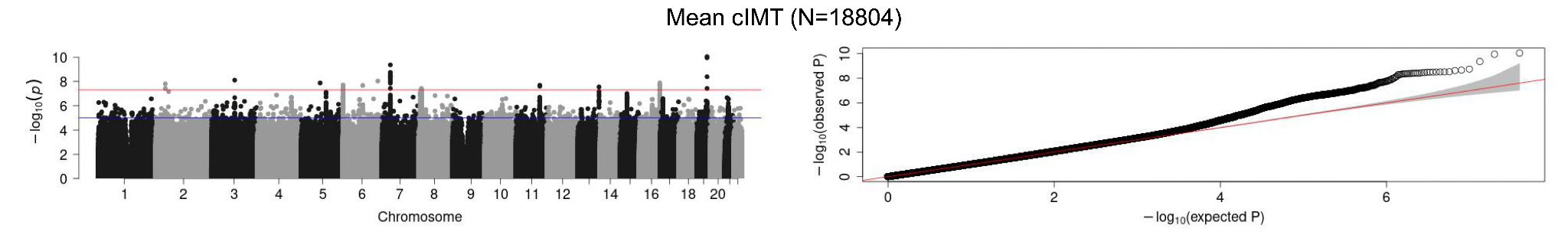

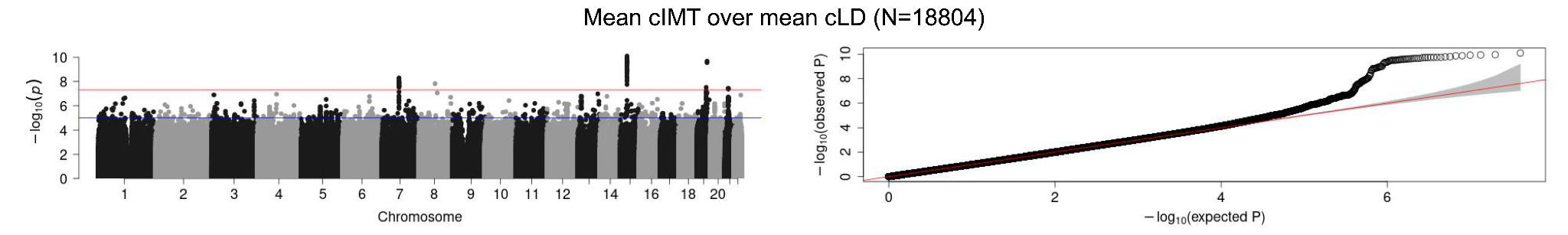

**Supplementary Figure 4 | Manhattan and Quantile-Quantile (QQ) plots.** On the left, Manhattan plots for each phenotype. The x-axis displays the positions across the 22 Chromosomes, while the y-axis represents the -log10(p) associated with each SNP. The red line indicates the Bonferroni threshold, above which the dots are considered significantly associated. On the right, QQ plots for each phenotype. The x-axis represents expected quantiles, while the y-axis represents observed quantiles. The points on the plot compare the observed quantiles of the phenotypes to the expected quantiles based on a theoretical distribution. The alignment between observed and expected quantiles indicates the similarity between the phenotype distributions and the theoretical distributions. The sample sizes for each analysis are reported in the title of each figure.

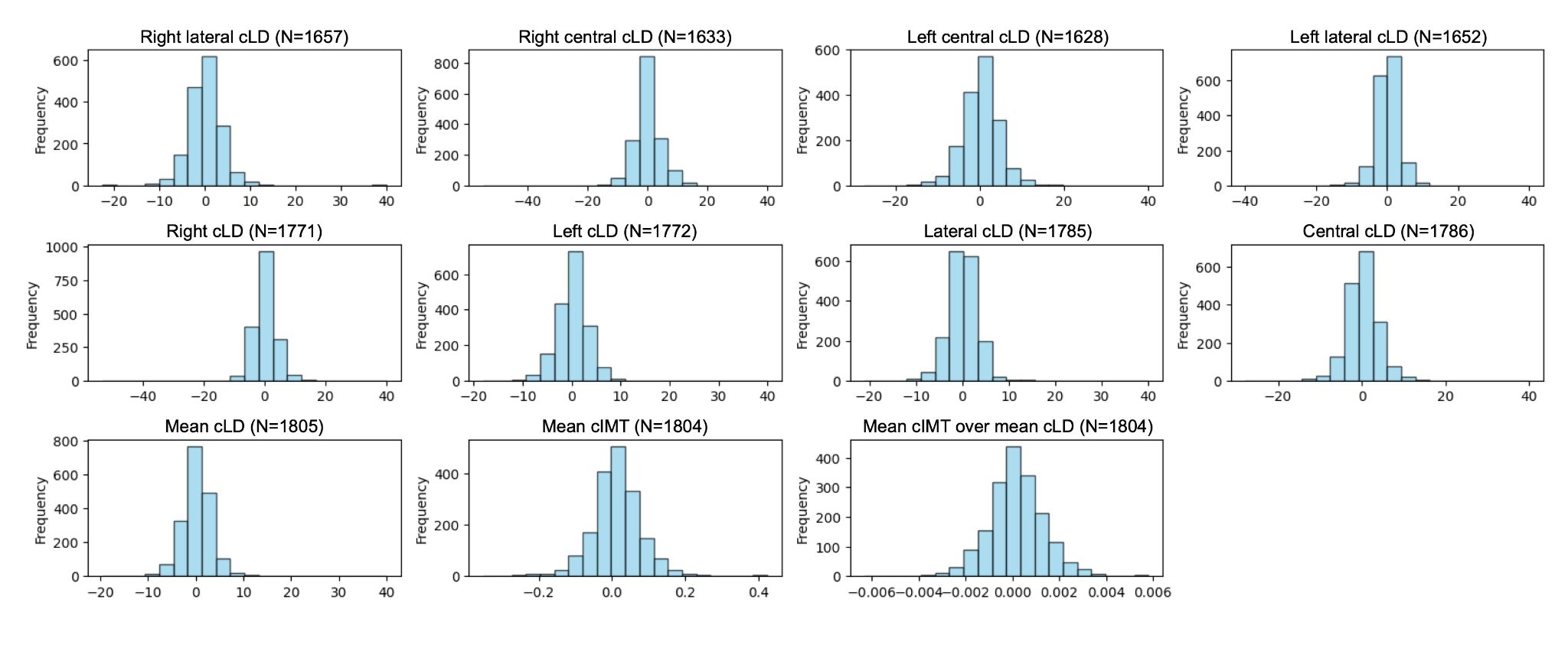

**Supplementary Figure 5 | Histogram of Phenotype Differences Between Instances (Subject Level).** Distributions of phenotype differences between the two instances for individual subjects. Each plot represents the differences calculated as the difference of *phenotype(instance 3)_i* and *phenotype(instance 2)_i*, where *i* represents individual subjects. The number in parentheses indicates the count of subjects with the respective phenotype measurements available for both instances. The distributions appear symmetric and do not exhibit significant deviations, suggesting the stability of the carotid lumen diameter measurements across the two time points. This consistency indicates that the phenotypes remain relatively unchanged over time, supporting their potential reliability as biomarkers.

**a.**

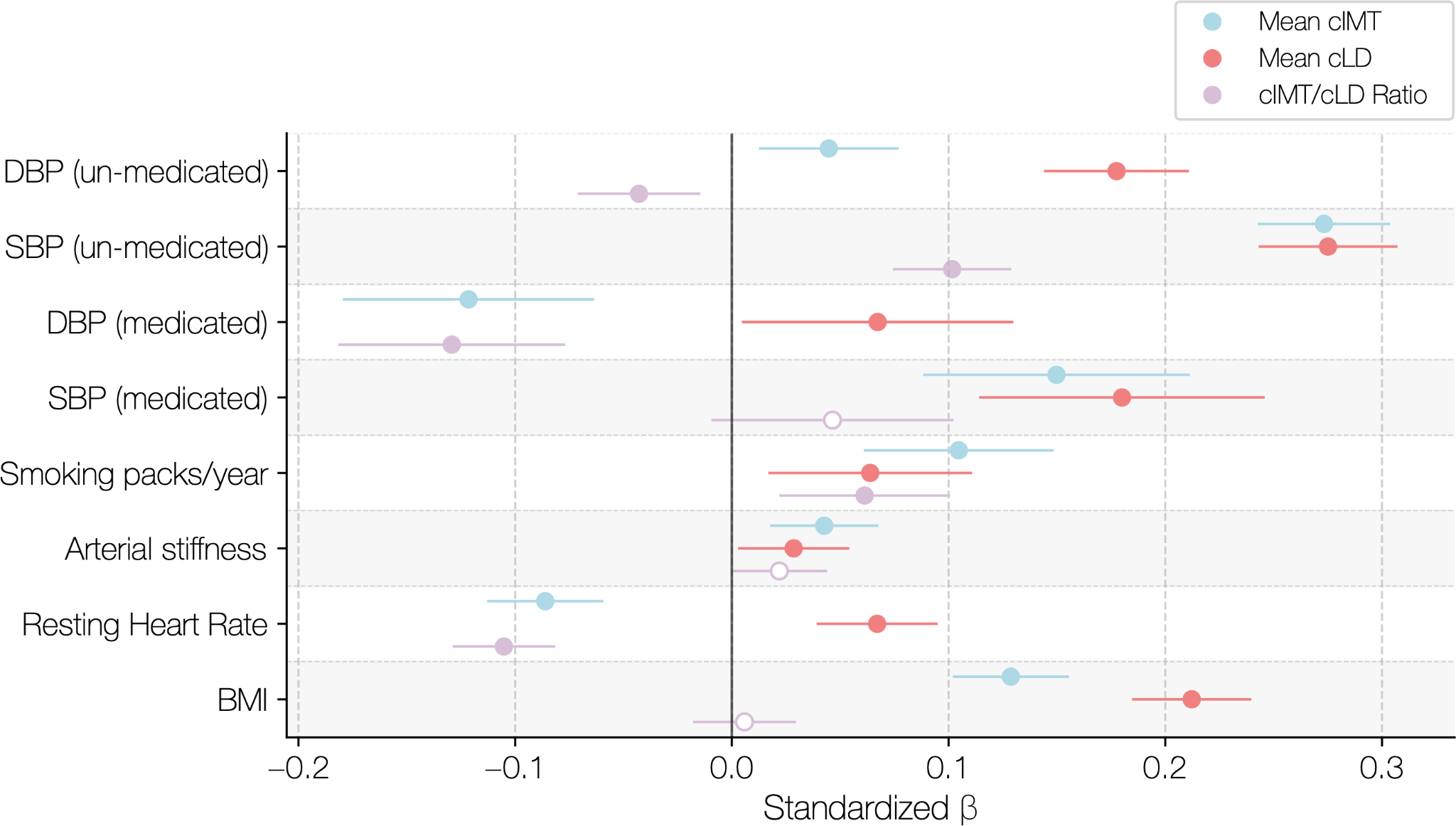

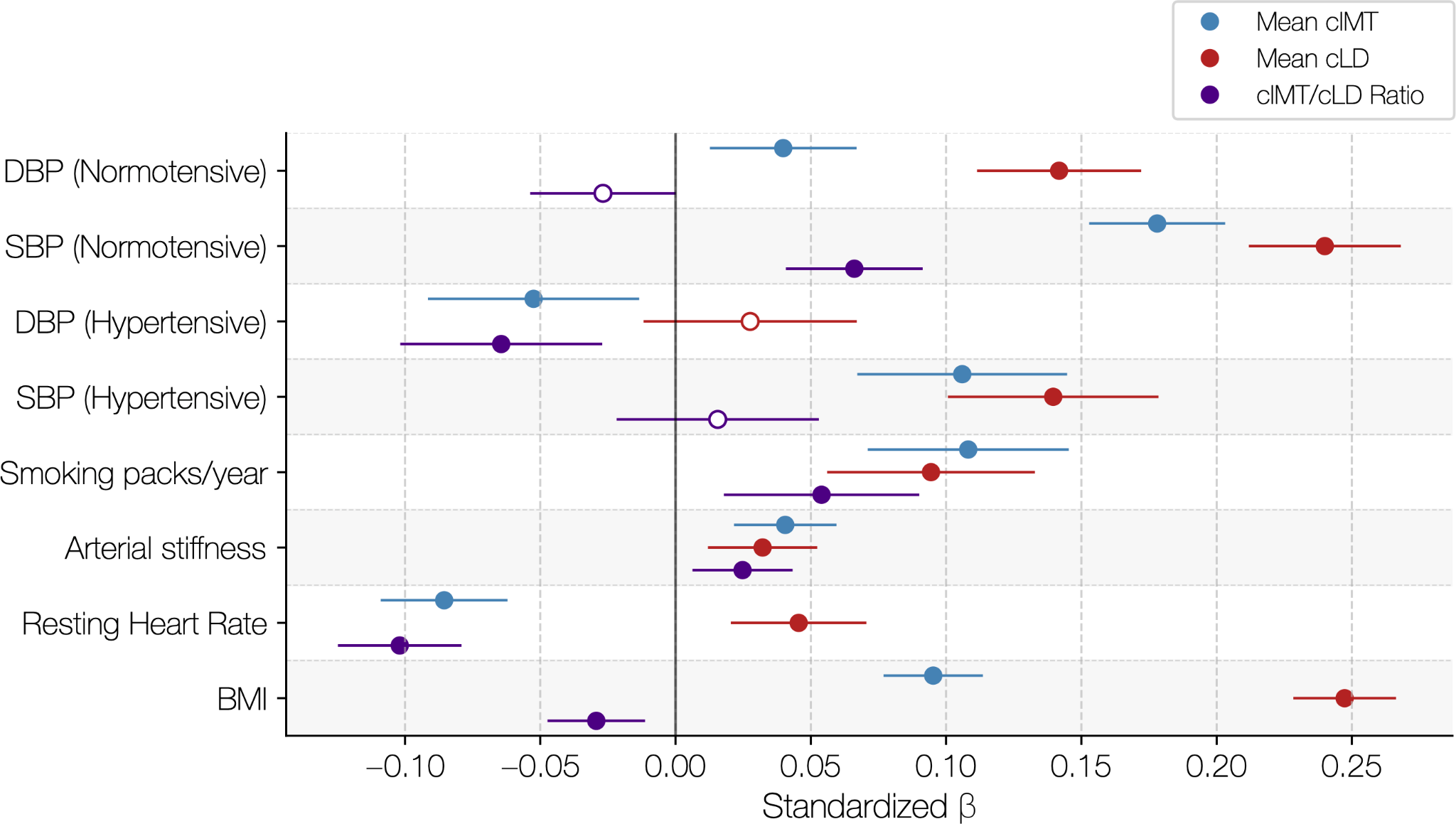

**b.**

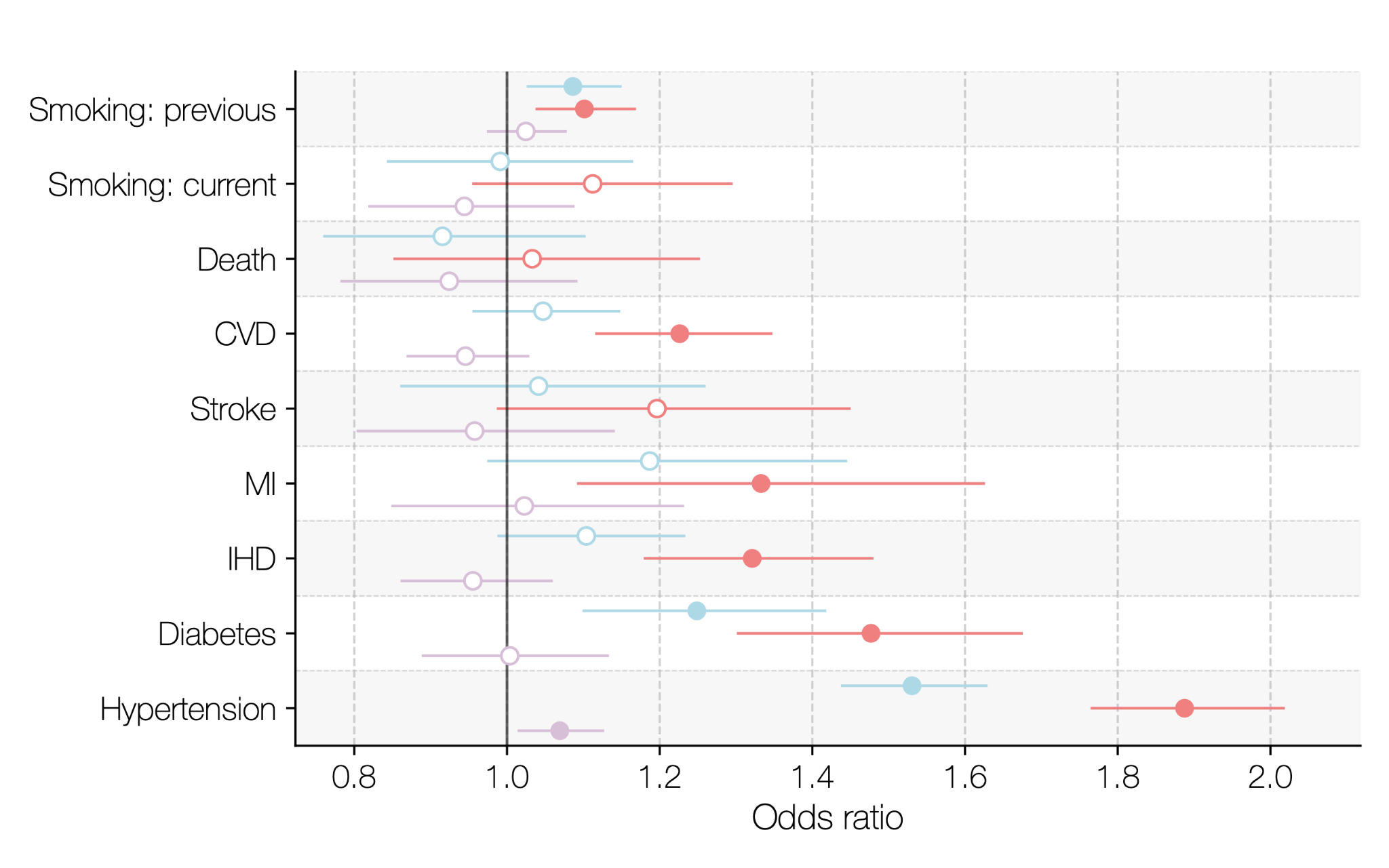

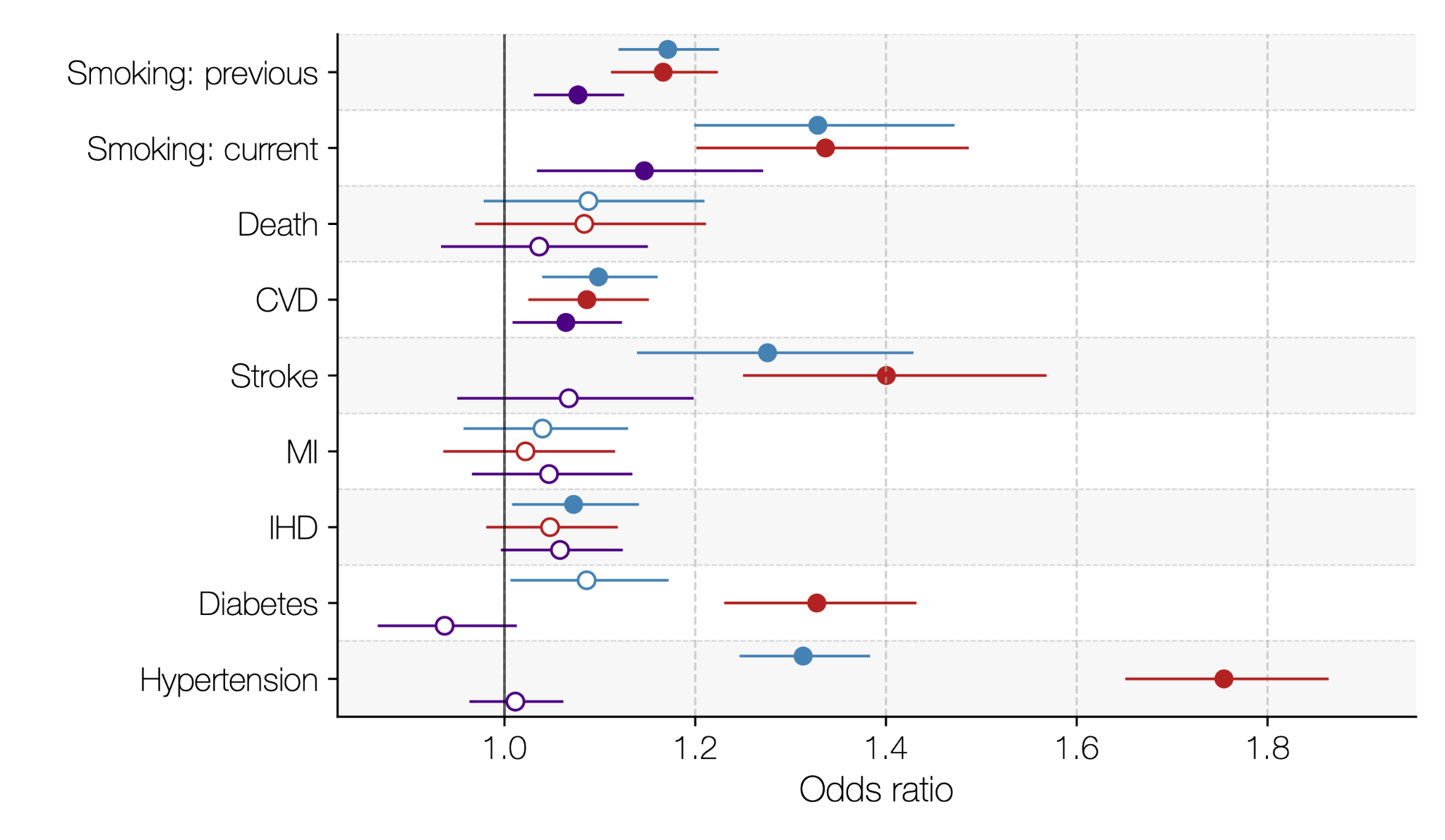

**Supplementary Figure 6** | **Diagnostic performance of mean cIMT, mean cLD, and their ratio in females and males.** **a)** Standardised regression coefficients from models assessing the independent associations of each phenotype with major continuous risk factors (adjusted for age and height). Females (N_F_ = 9 522) are displayed on the left and males (N_M_ = 9 245) on the right. **b)** Odds ratios from analogous models evaluating associations with major categorical risk factors and prevalent disease outcomes (adjusted for age and height). Abbreviations: SBP/DBP: systolic/diastolic blood pressure; BMI: body mass index; MI: myocardial infarction; IHD: ischaemic heart disease.

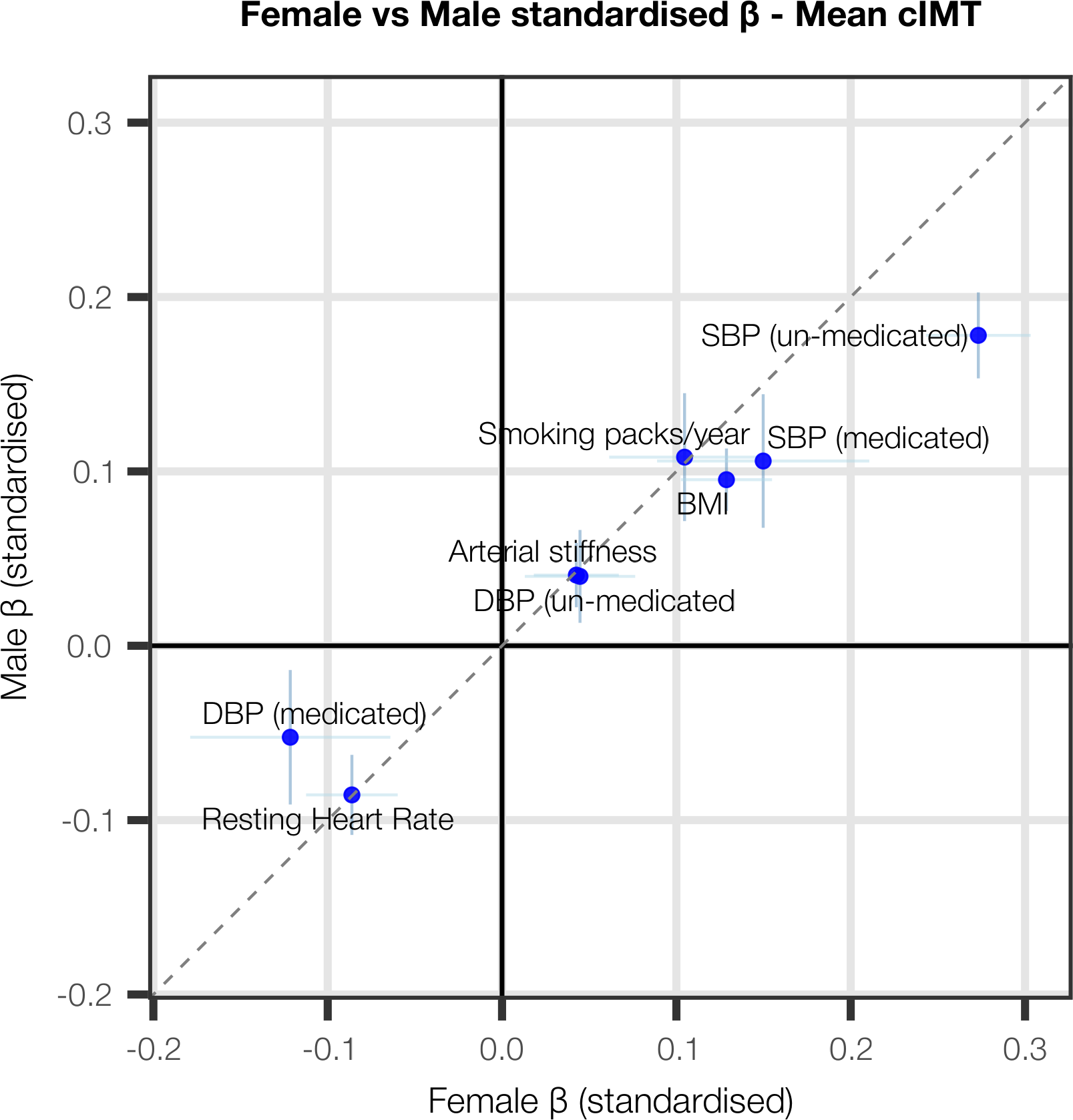

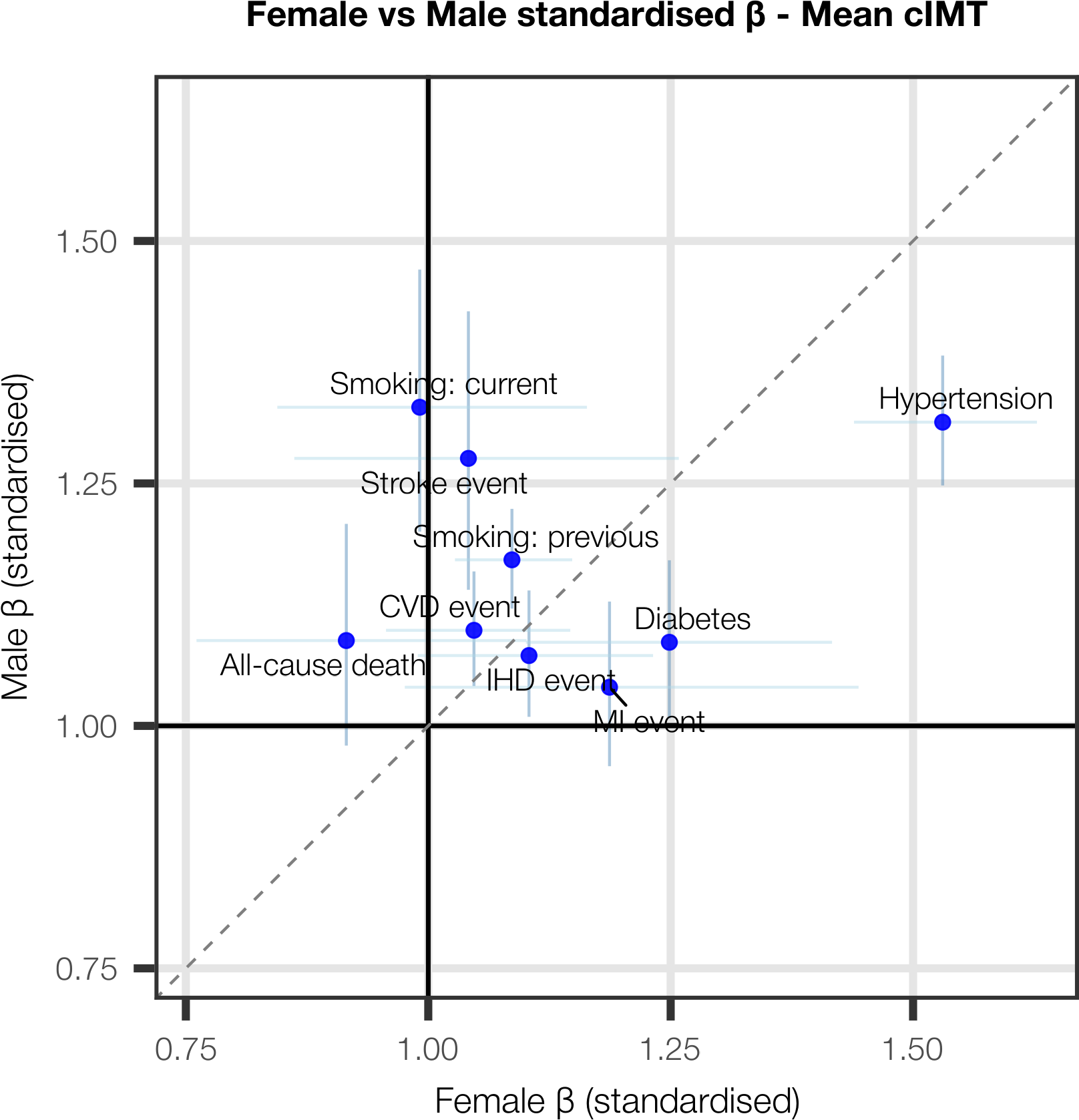

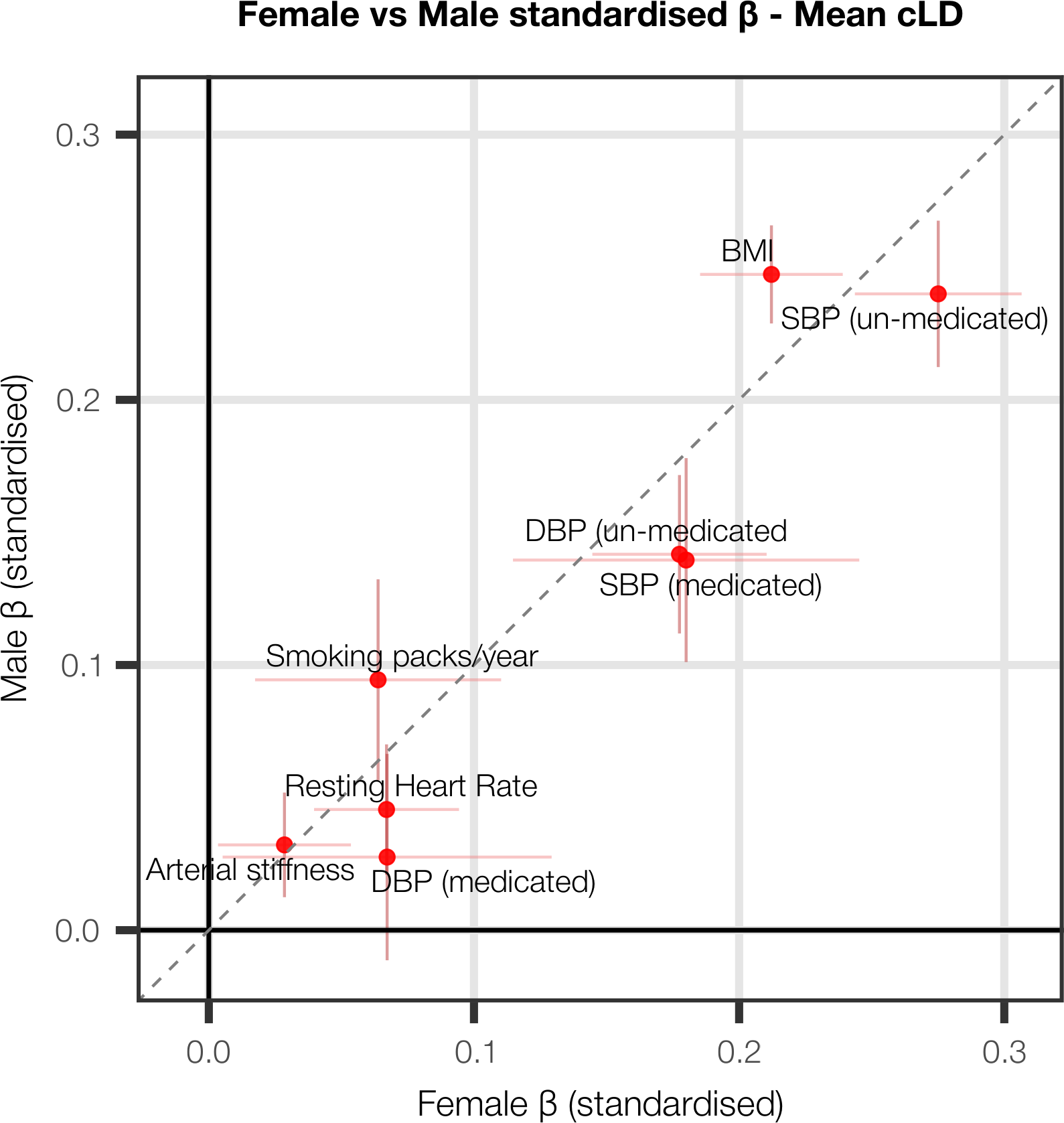

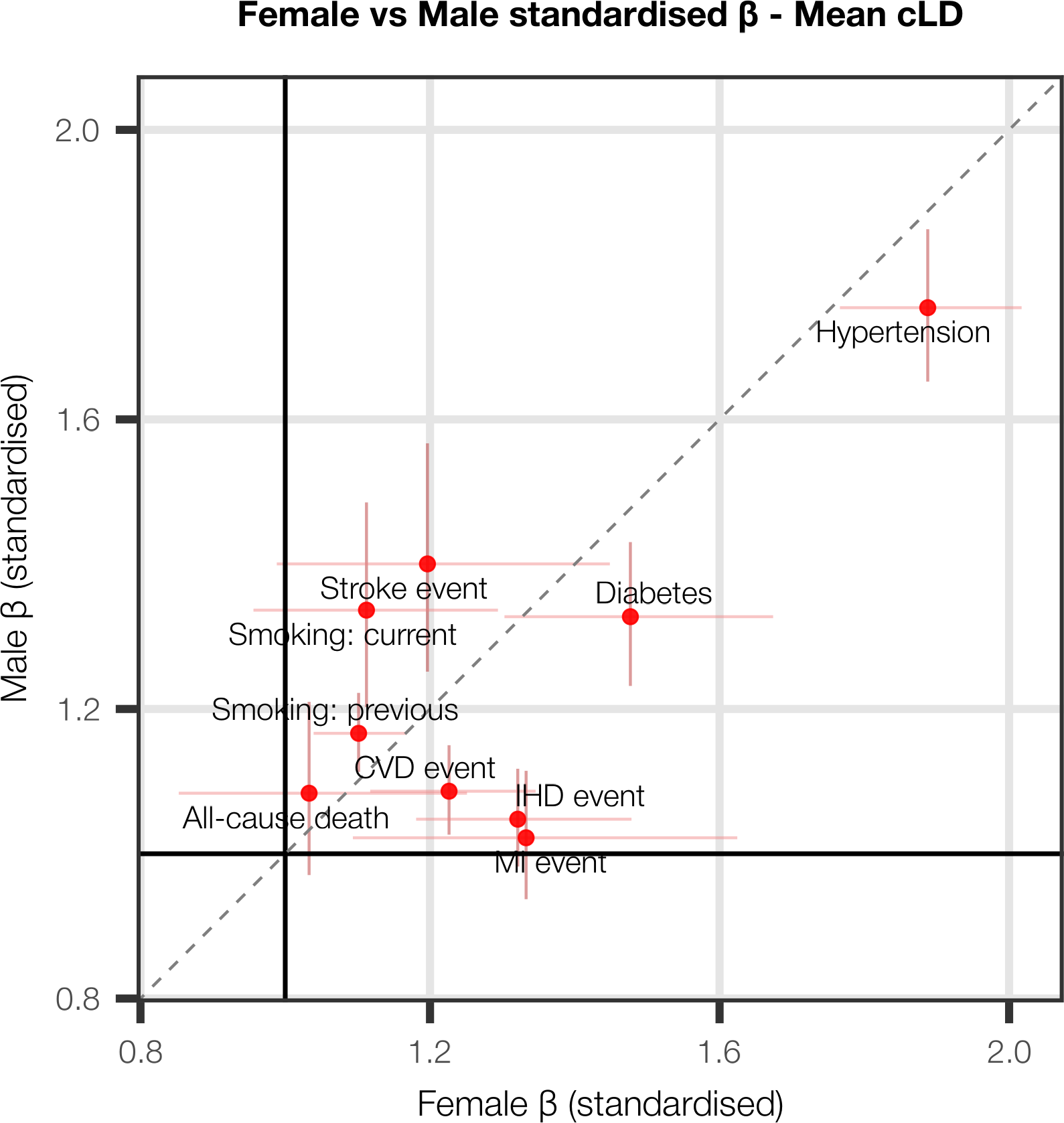

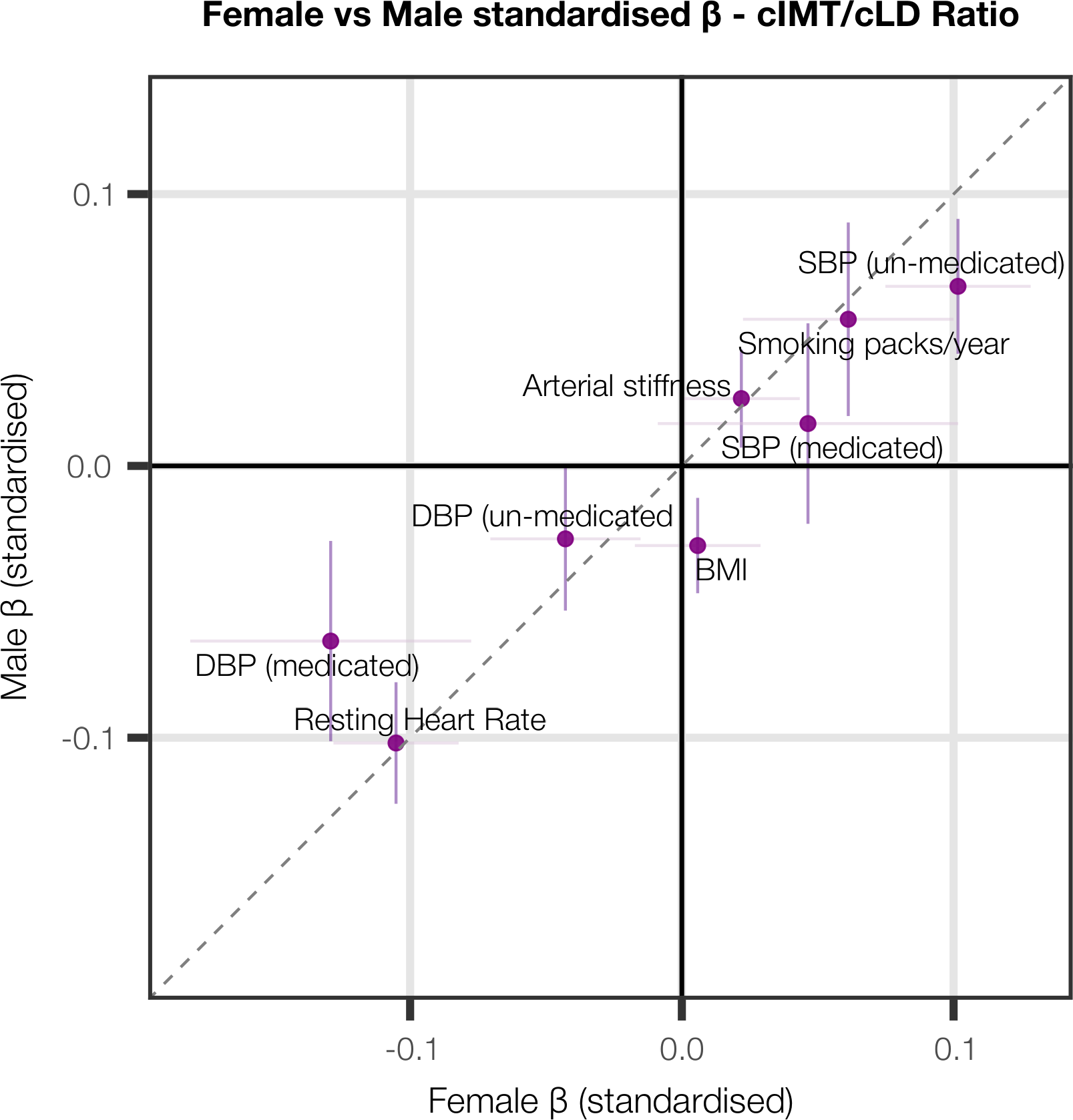

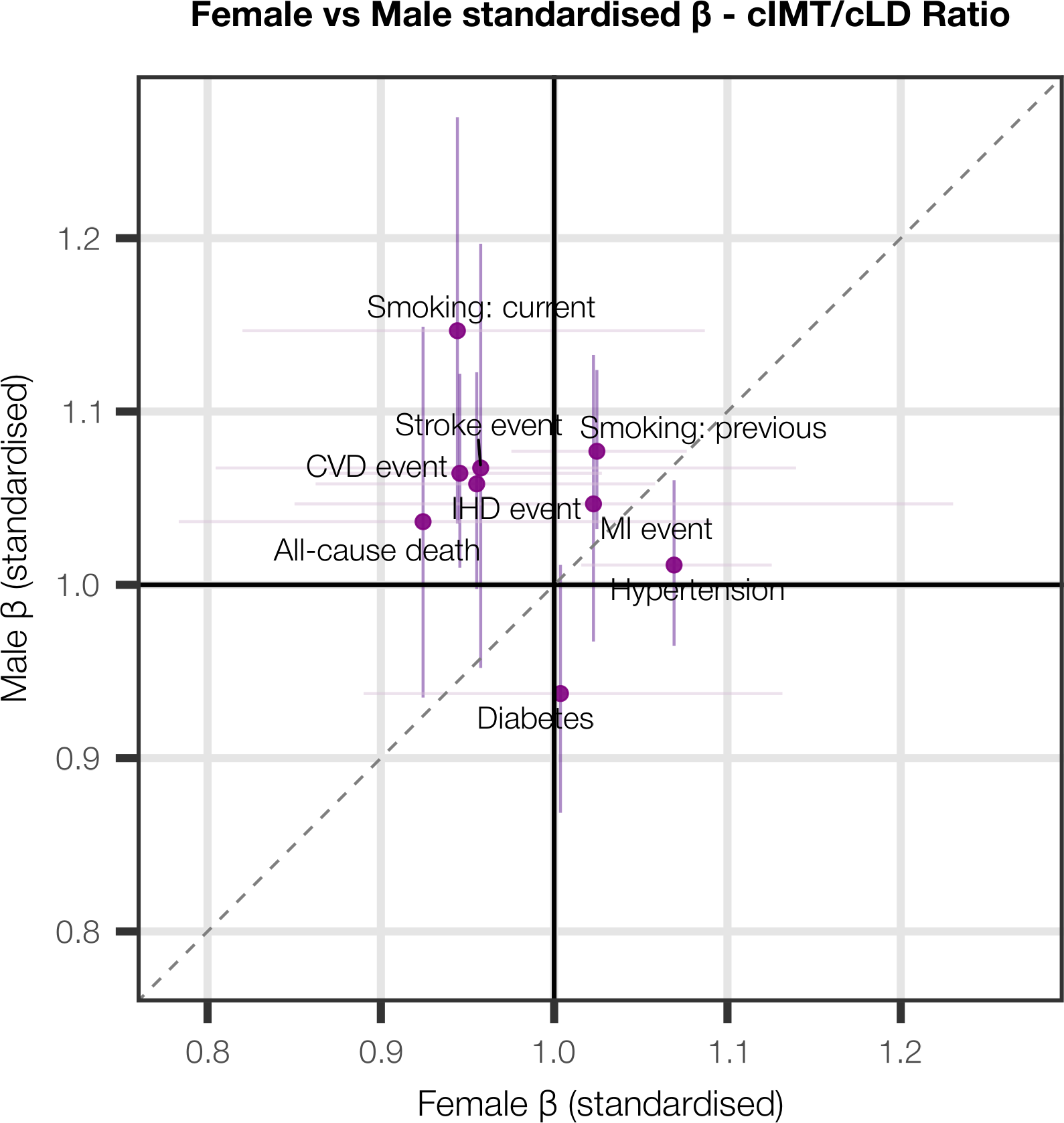

**Supplementary Figure 7** | **Diagnostic performance of mean cIMT, mean cLD, and their ratio in males and females.** Standardised regression coefficients and odds ratios from models assessing the independent associations of each phenotype with major continuous risk factors (adjusted for age and height). Females (N_F_ = 9 522) are plotted on the x axis and males (N_M_ = 9 245) on the y axis. Abbreviations: SBP/DBP: systolic/diastolic blood pressure; BMI: body mass index; MI: myocardial infarction; IHD: ischaemic heart disease.

**a.**

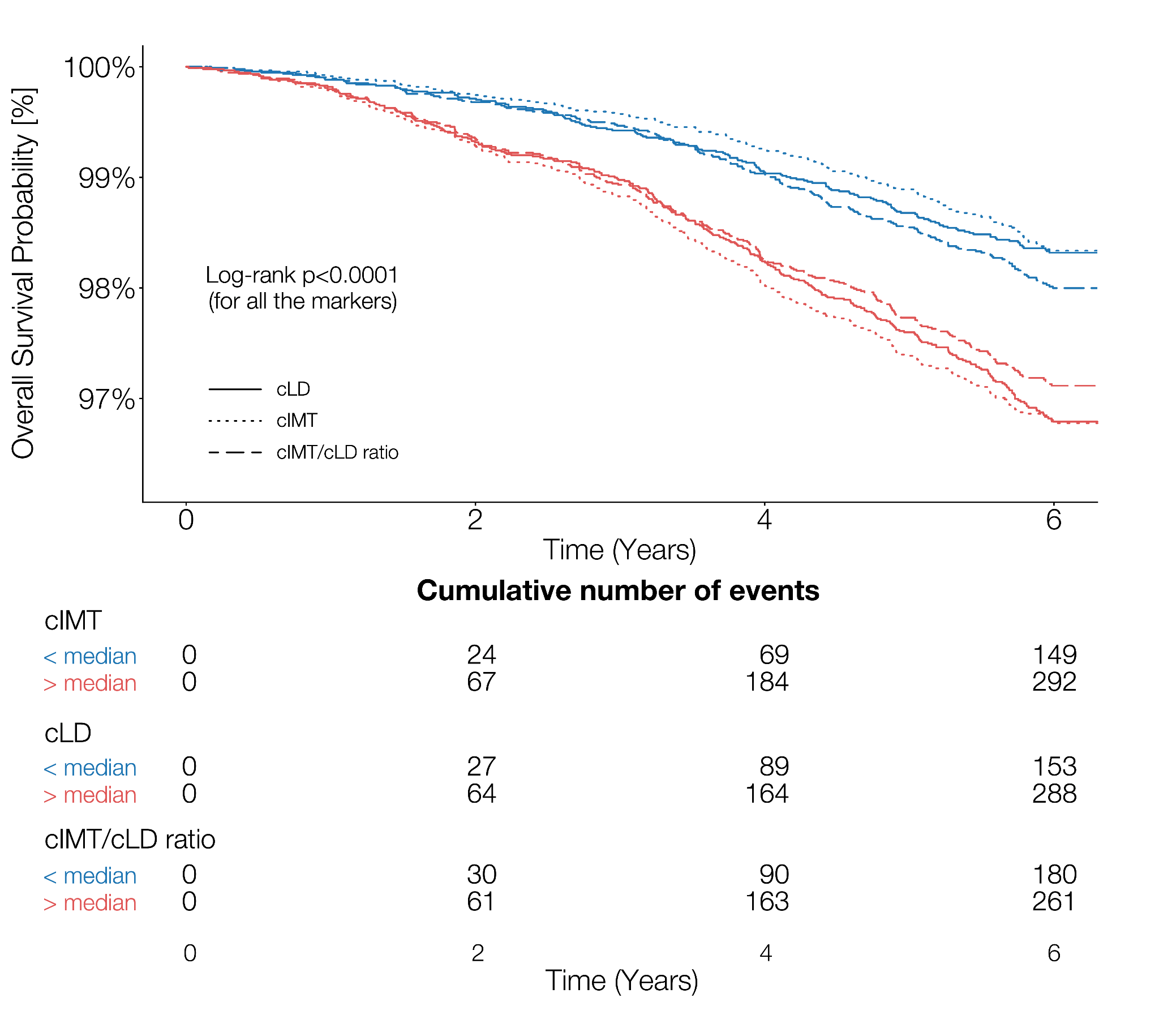

**b.**

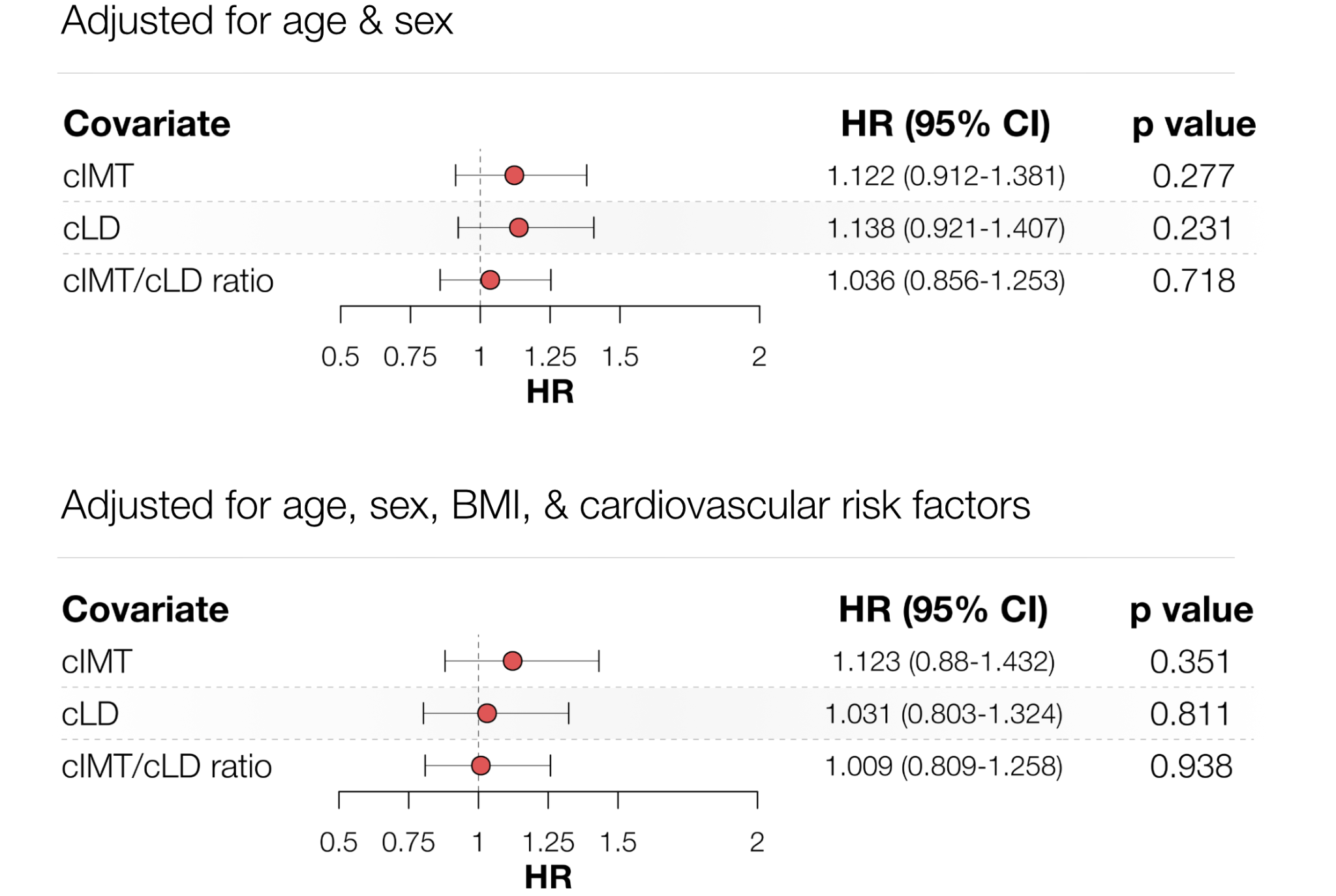

**c.**

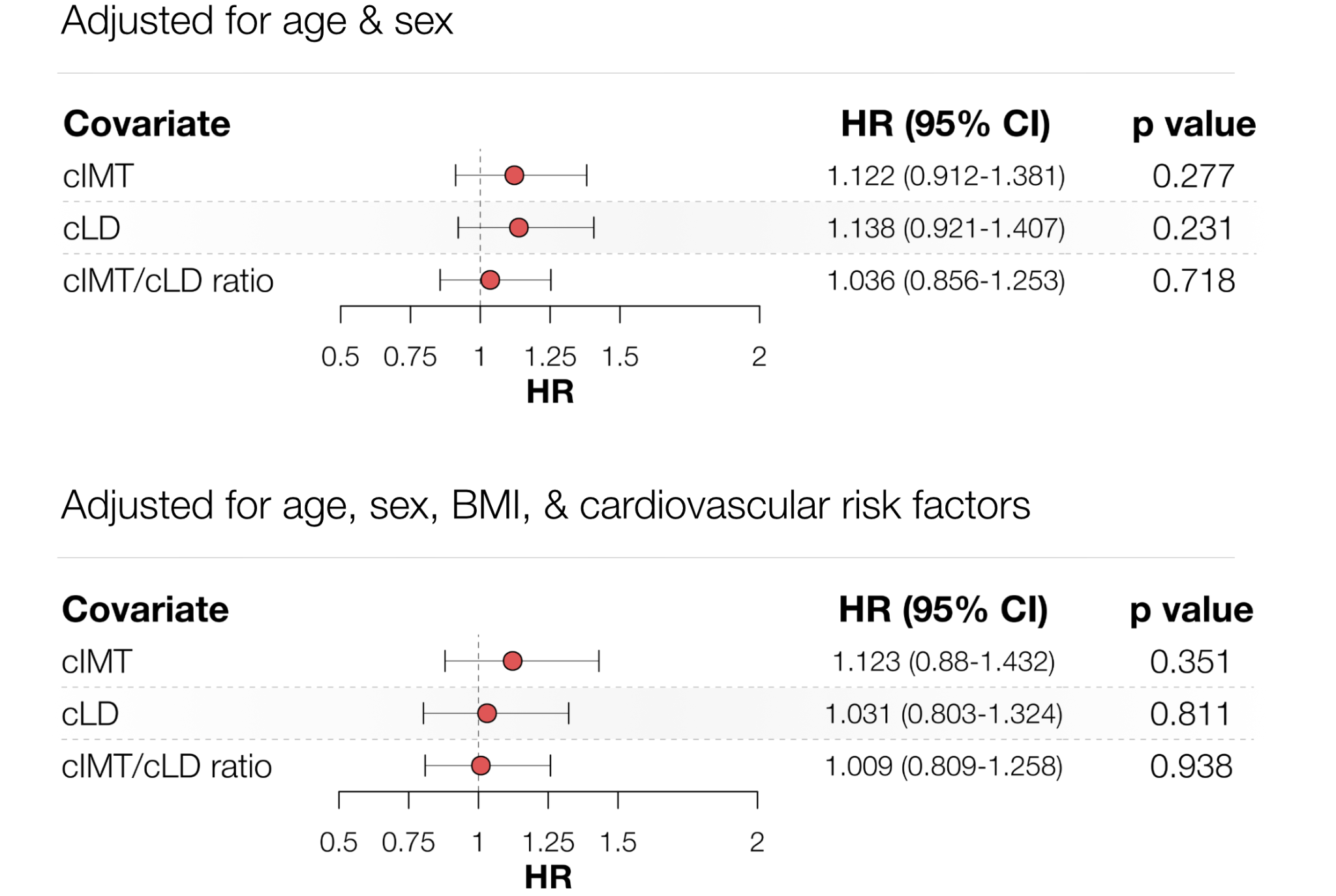

**Supplementary Figure 8 | Prognostic performance of mean cIMT, mean cLD, and their ratio for all-cause mortality.** **a)** Kaplan-Meier survival analysis of the three different phenotypes for all-cause mortality (N=18 767). The two groups were obtained by splitting the data on the median of the respective phenotype. **b)** Forest plots for the three different phenotypes for all-cause mortality. Here, the hazard ratios (HR) are calculated relative to individuals with phenotype values below the median. The HRs and the p-values were obtained from three independent Cox models (one per phenotype) adjusted for age and sex (N=18 767; total number of all-cause deaths: 441). In **c)**, we additionally adjusted for BMI, systolic blood pressure, smoking status, diabetes, blood pressure medications, and previous cardiovascular events (N=15 011 after excluding non-complete observations; total number of all-cause deaths: 329).

**a.**

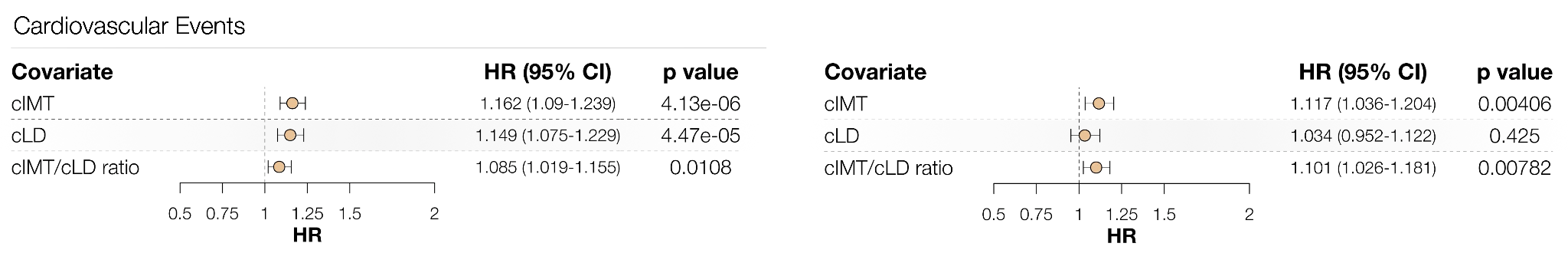

**b.**

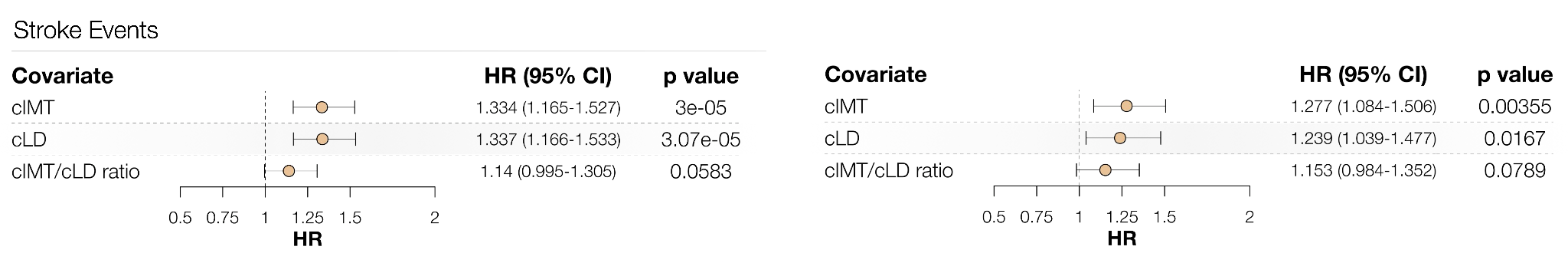

**c.**

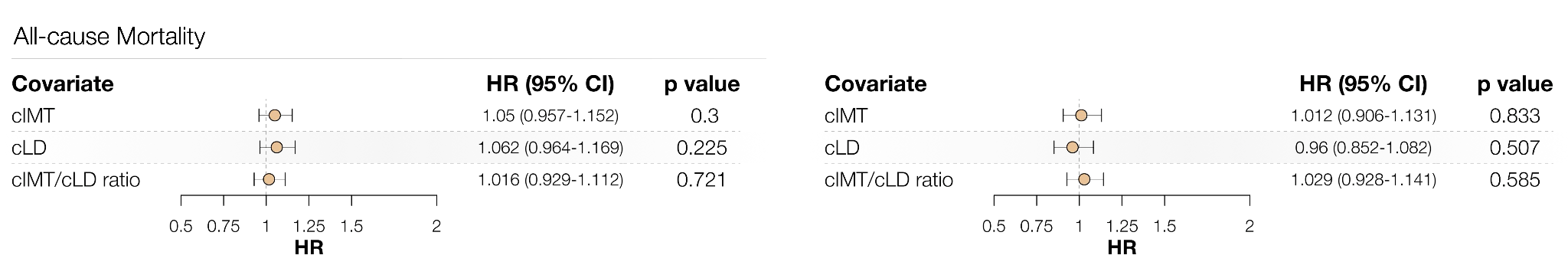

**Supplementary Figure 9 | Survival analysis of the continuous traits**. Hazard Ratios (HRs) for the three carotid phenotypes for **a)** cardiovascular events, **b)** stroke events, and **c)** all-cause mortality. All the HRs and the p-values were obtained from three independent Cox models (one per phenotype, after z-scoring, in a continuous setting) after adjusting for age and sex (on the left; N=18 767; total number of cardiovascular events: 893; total number of stroke events: 179; total number of all-cause deaths: 441), and additionally adjusted for BMI, systolic blood pressure, smoking status, diabetes, blood pressure medications, and previous cardiovascular events (on the right; N=15 011 after excluding non-complete observations; total number of cardiac events: 685; total number of stroke events: 129; total number of all-cause deaths: 329).

**a.**

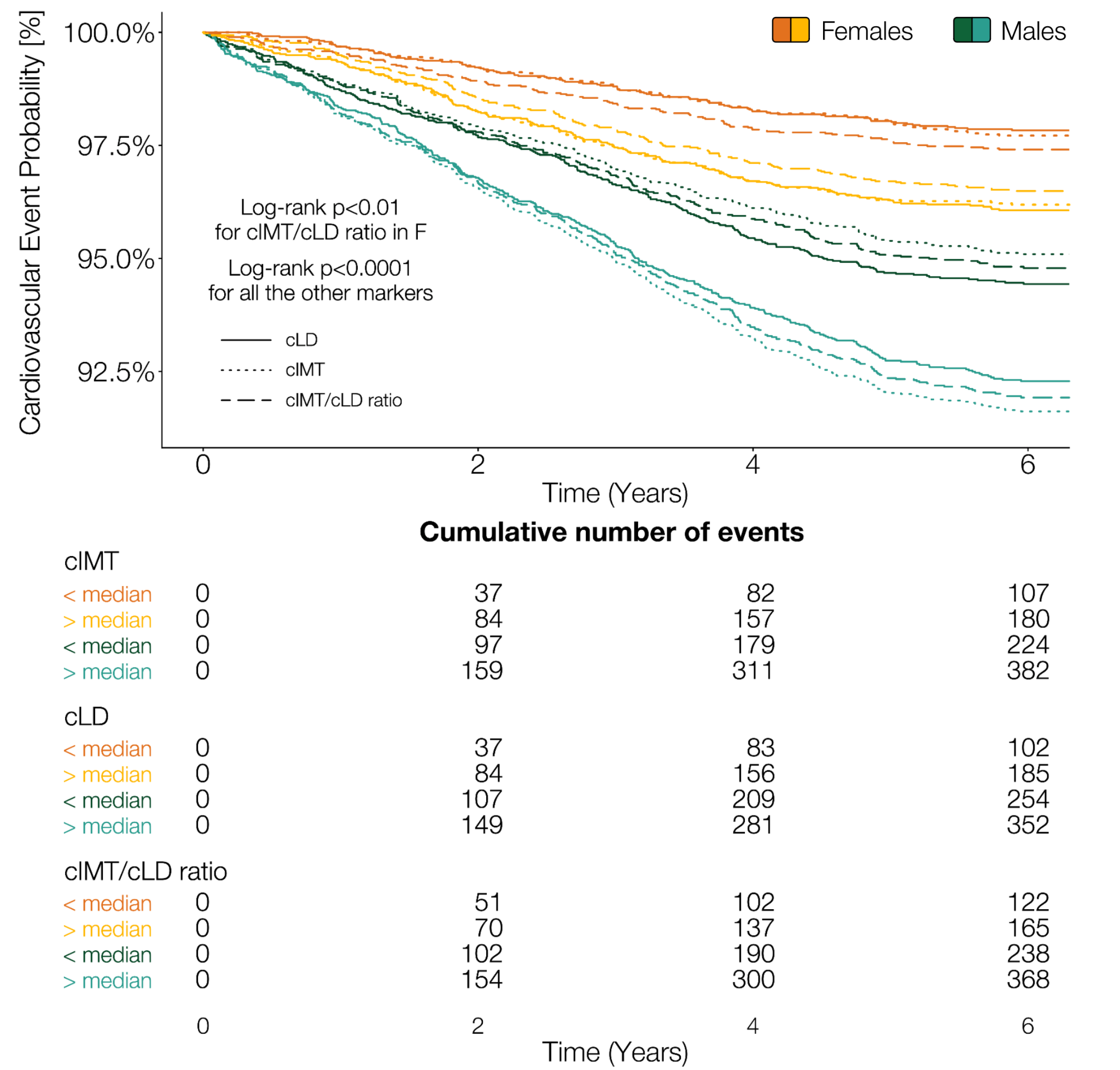

**b.**

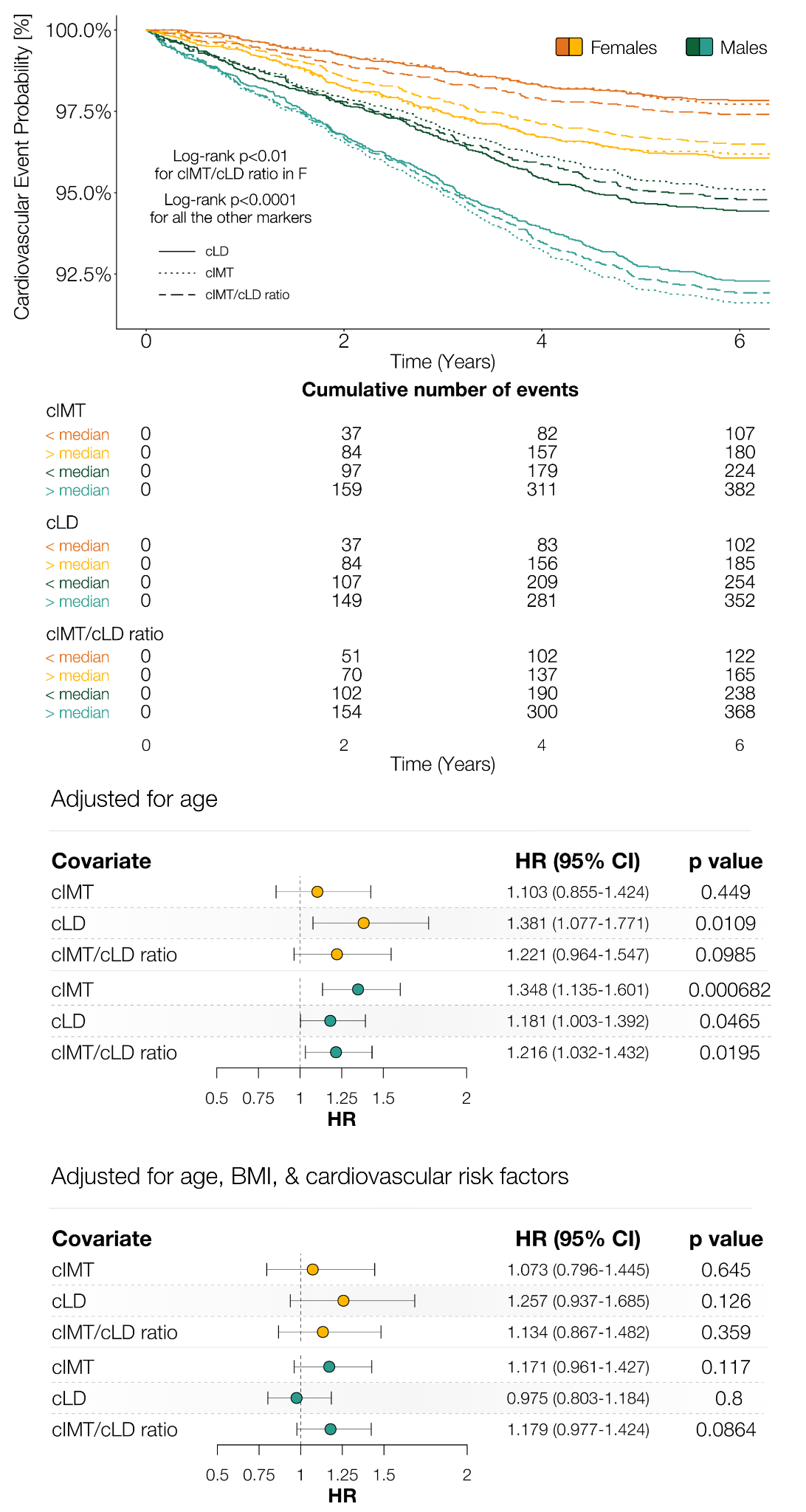

**c.**

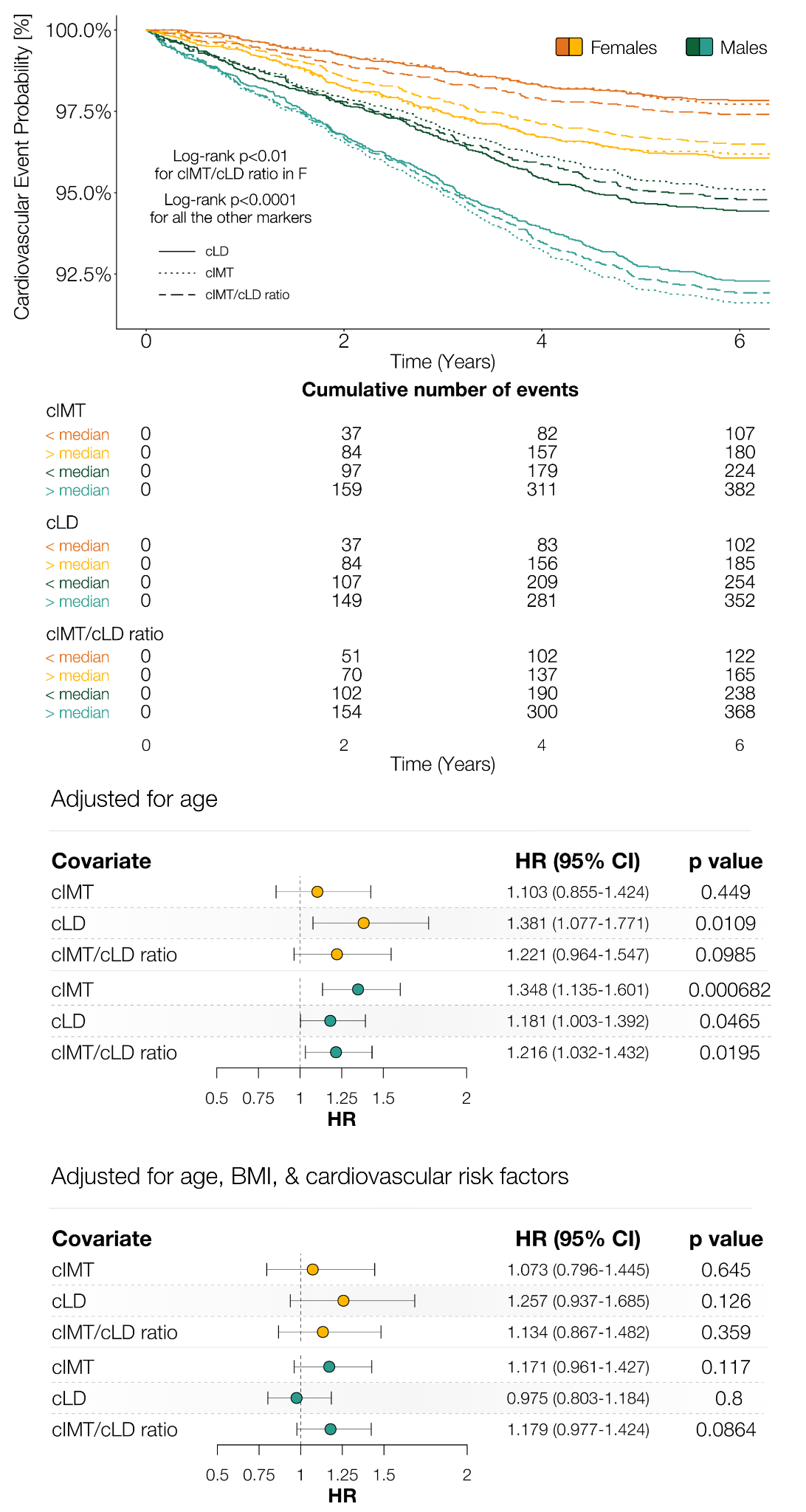

**Supplementary Figure 10 | Sex-specific prognostic performance of mean cIMT, mean cLD, and their ratio for cardiovascular events.**

**a.**

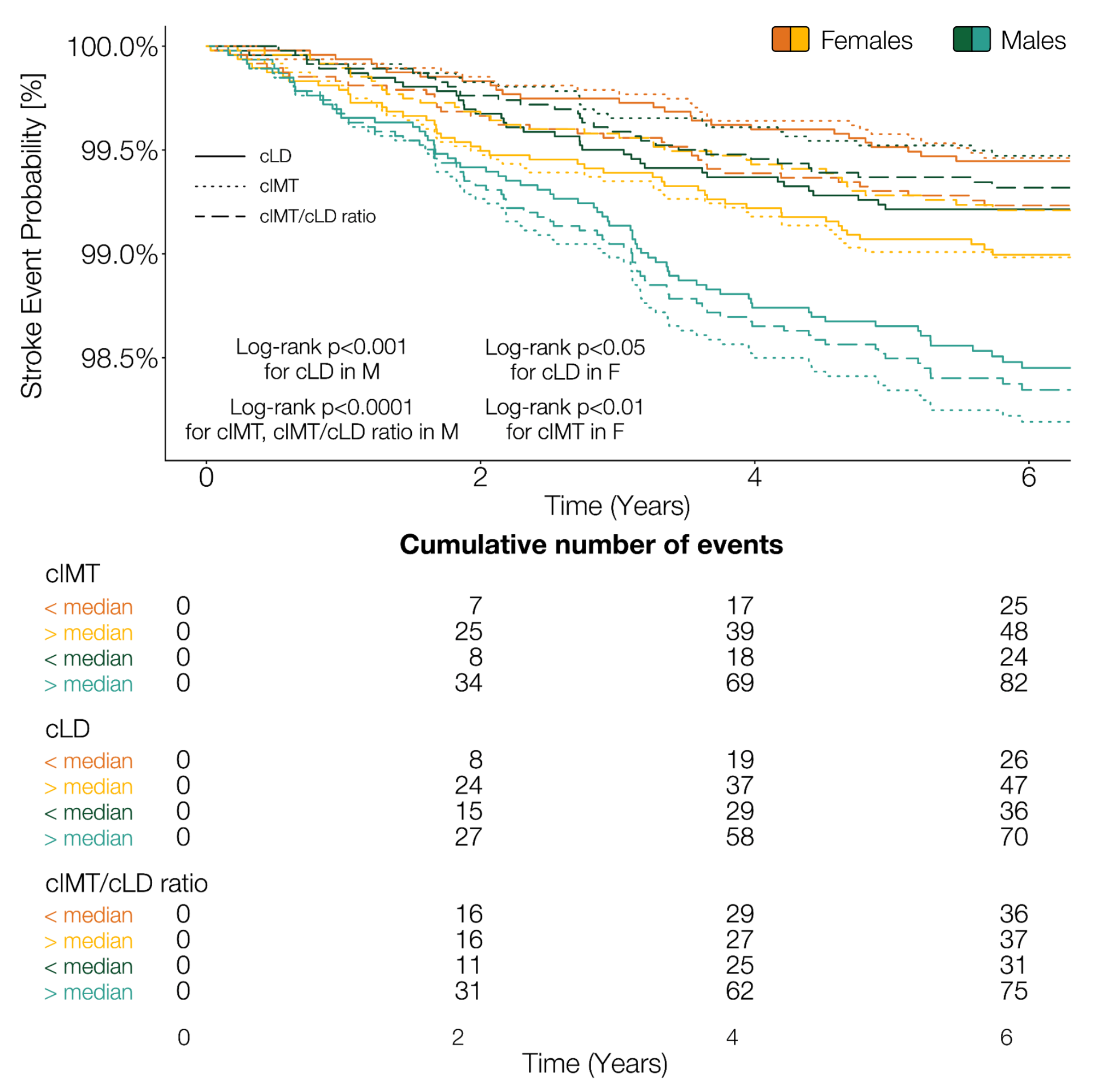

**b.**

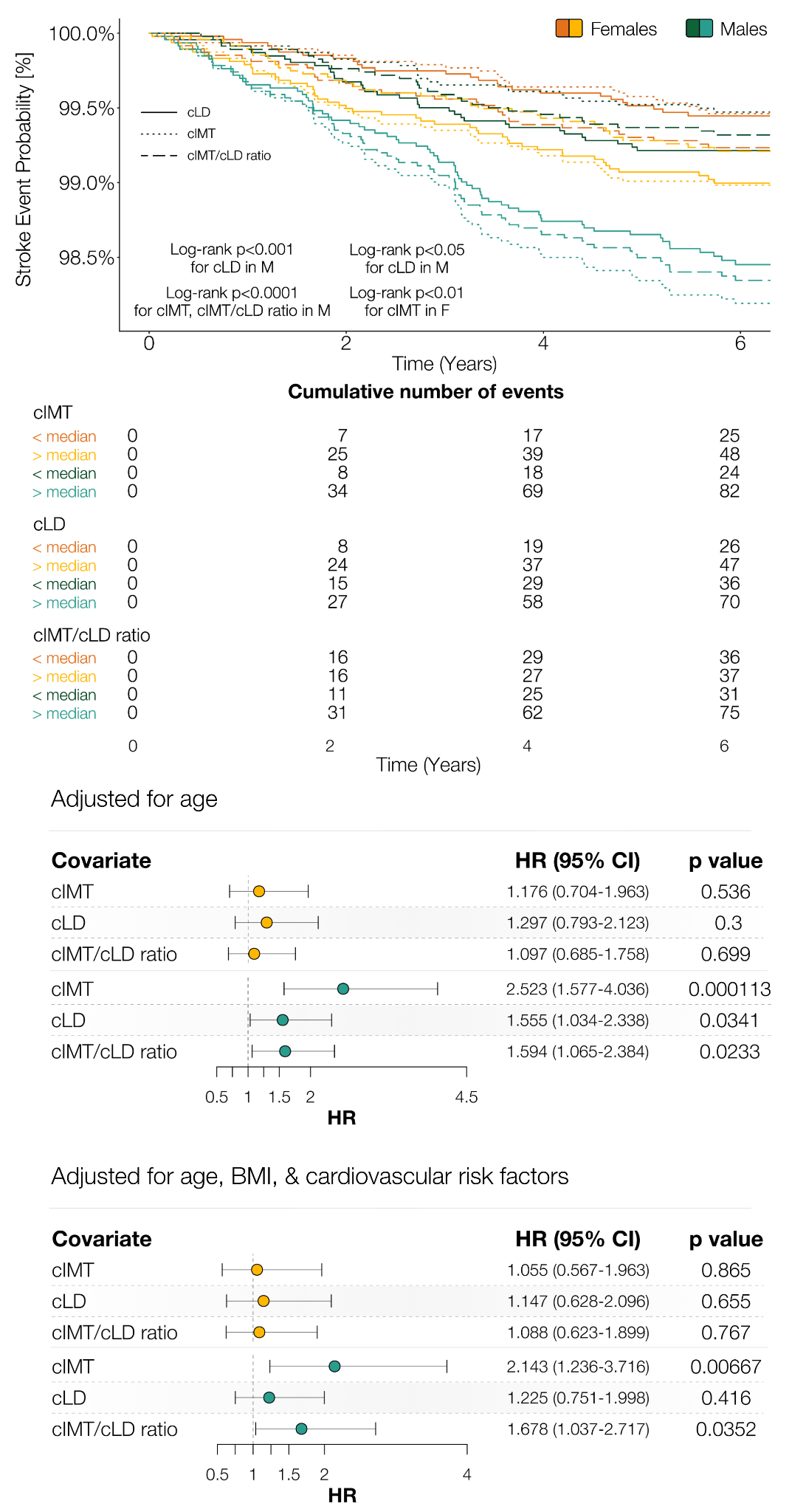

**c.**

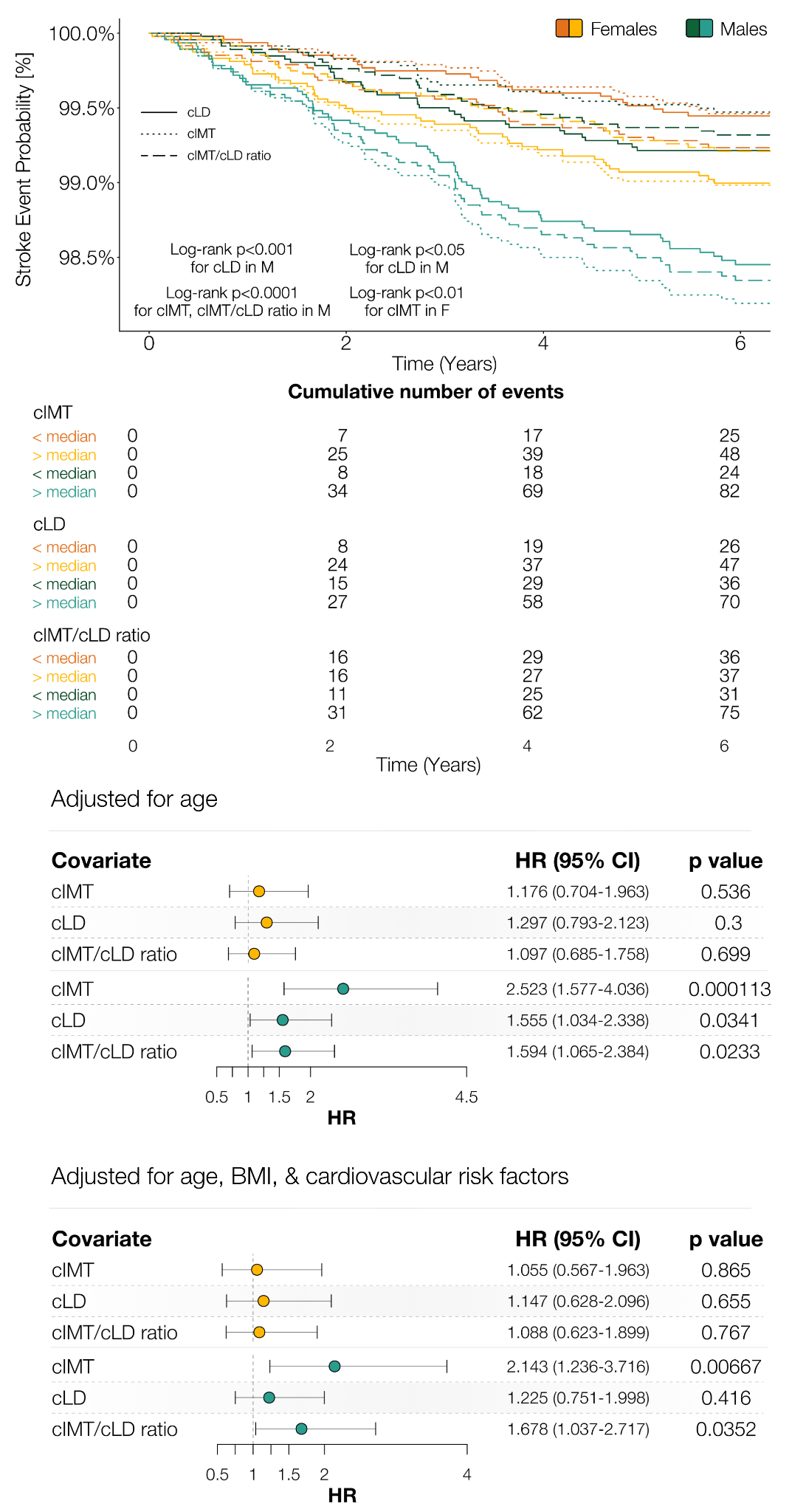

**Supplementary Figure 11 | Sex-specific prognostic performance of mean cIMT, mean cLD, and their ratio for stroke events.**

**a.**

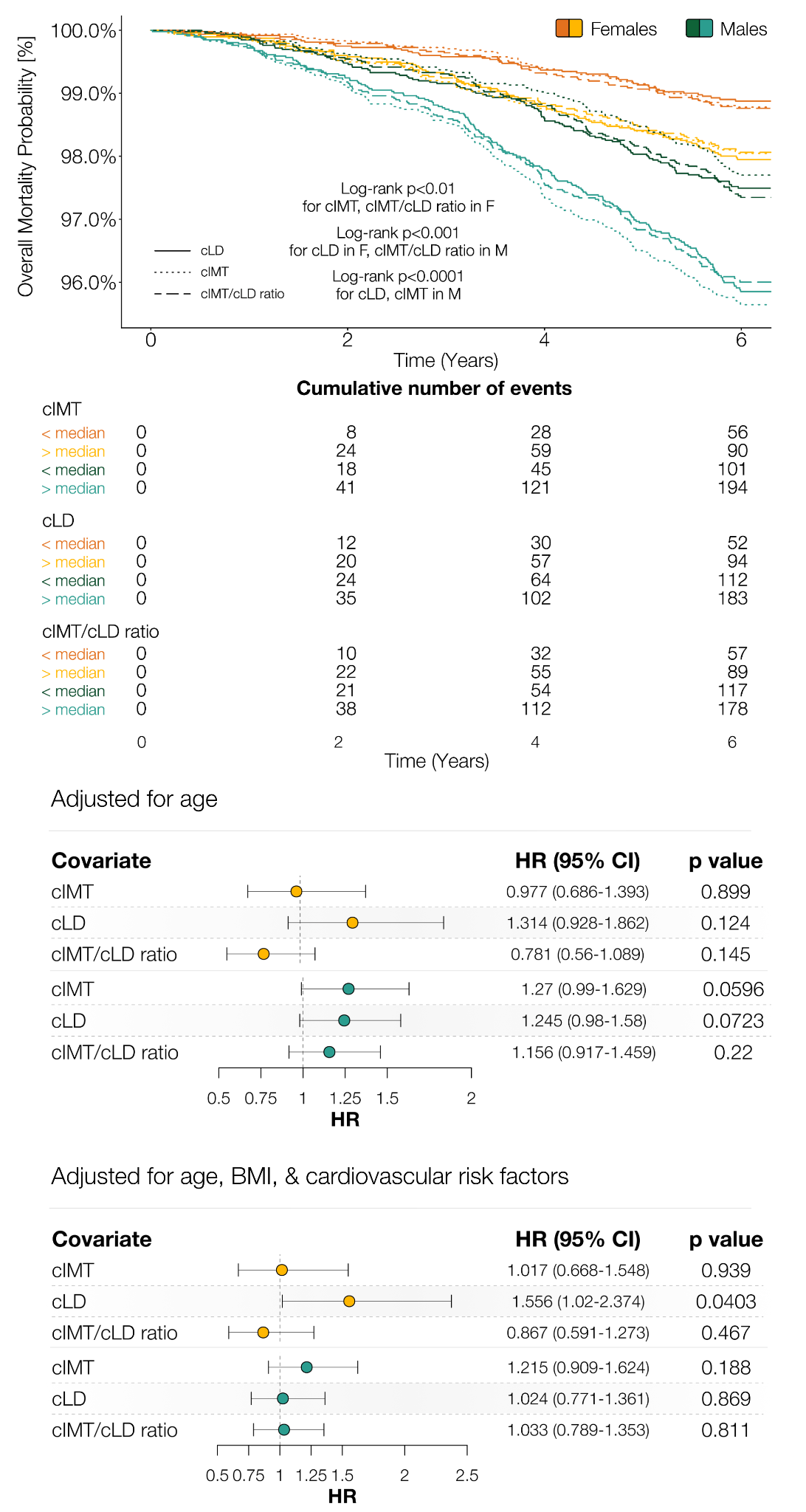

**b.**

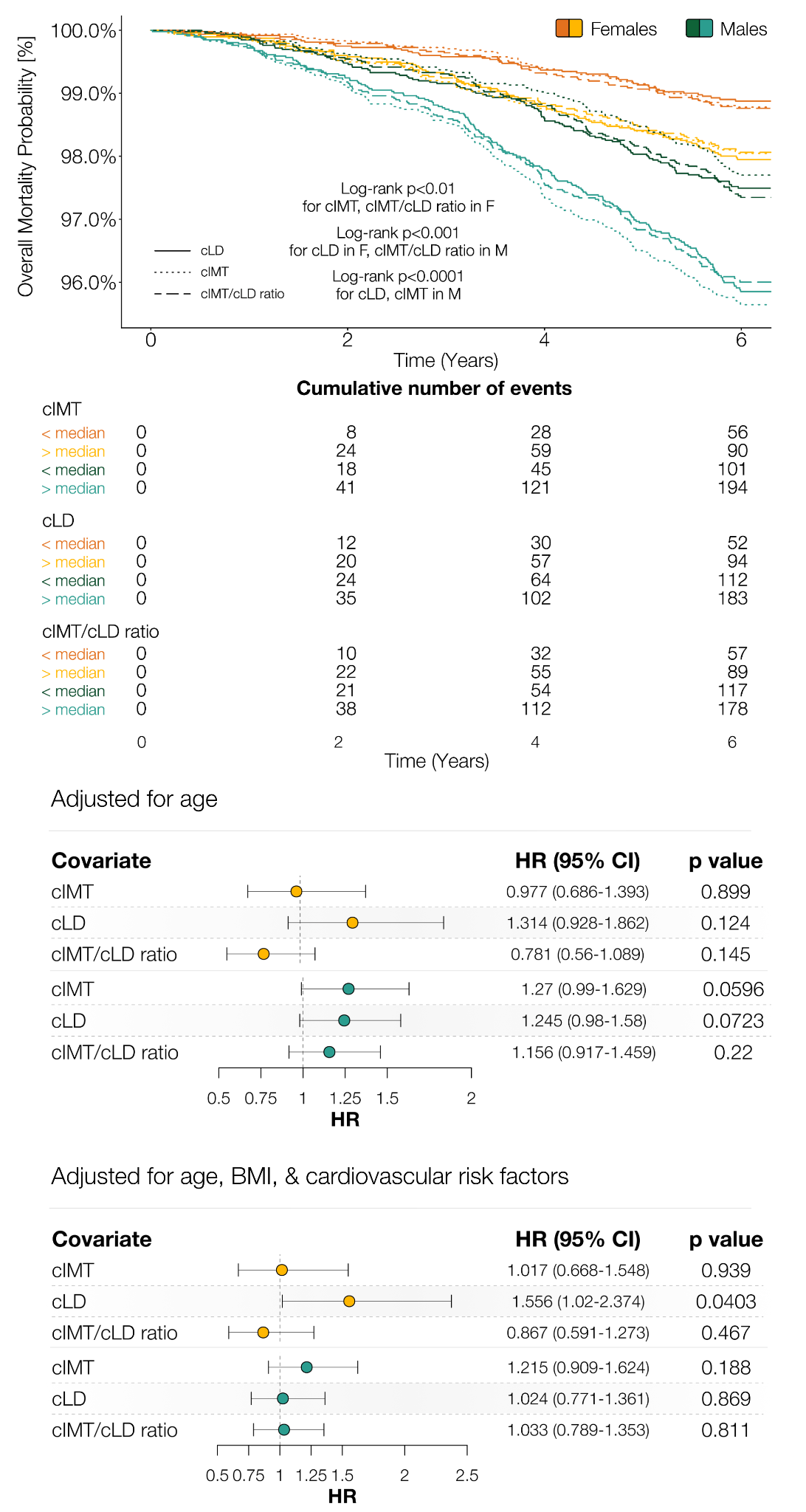

**c.**

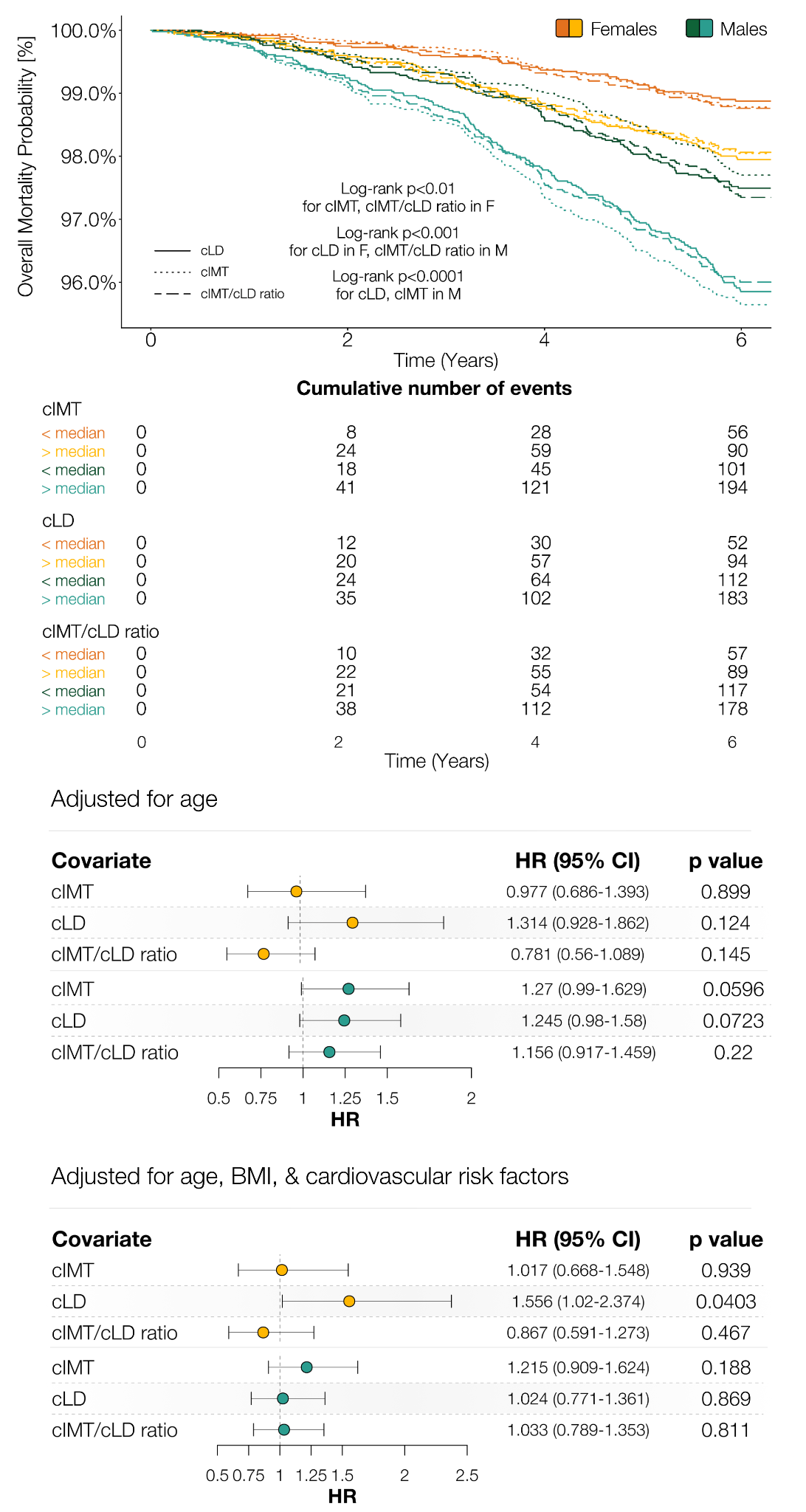

**Supplementary Figure 12 | Sex-specific prognostic performance of mean cIMT, mean cLD, and their ratio for all-cause mortality.**
