## Supplementary Tables for "Genetic and Etiological Insights from Automated Lumen Diameter Measurements in Carotid Ultrasounds of the UK Biobank"

**a.**

|  | **Right lateral cLD** | **Right central cLD** | **Left central cLD** | **Left lateral cLD** | **Right cLD** | **Left cLD** | **Lateral cLD** | **Central cLD** | **Mean cLD** |
| --- | --- | --- | --- | --- | --- | --- | --- | --- | --- |
| **Right lateral cLD** |  | 0.95 | 1.03 | 0.96 | 0.99 | 1.01 | 0.95 | 0.99 | 0.99 |
| **Right central cLD** | 0.894842 |  | 0.95 | 1 | 0.99 | 0.93 | 1 | 0.99 | 0.98 |
| **Left central cLD** | 0.701048 | 0.689665 |  | 0.94 | 0.98 | 1 | 0.92 | 1 | 0.98 |
| **Left lateral cLD** | 0.74509 | 0.695949 | 0.89259 |  | 0.98 | 0.94 | 1 | 0.99 | 0.97 |
| **Right cLD** | 0.974482 | 0.973891 | 0.70947 | 0.736385 |  | 0.98 | 0.98 | 1 | 1 |
| **Left cLD** | 0.736279 | 0.705173 | 0.972331 | 0.974074 | 0.73415 |  | 0.92 | 0.99 | 0.97 |
| **Lateral cLD** | 0.939532 | 0.846346 | 0.839221 | 0.933951 | 0.910925 | 0.903525 |  | 0.98 | 0.97 |
| **Central cLD** | 0.858609 | 0.925648 | 0.918945 | 0.850551 | 0.909259 | 0.899943 | 0.903036 |  | 0.99 |
| **Mean cLD** | 0.920699 | 0.90541 | 0.899153 | 0.914311 | 0.93332 | 0.926634 | 0.975203 | 0.973993 |  |

**b.**

|  | **Right lateral cLD** | **Right central cLD** | **Left central cLD** | **Left lateral cLD** | **Right cLD** | **Left cLD** | **Lateral cLD** | **Central cLD** | **Mean cLD** |
| --- | --- | --- | --- | --- | --- | --- | --- | --- | --- |
| **Right lateral cLD** |  | 0.0426 | 0.0266 | 0.034 | 0.0113 | 0.0075 | 0.032 | 0.0093 | 0.0202 |
| **Right central cLD** | 0.0426 |  | 0.0447 | 0.0255 | 0.0176 | 0.0401 | 0.0078 | 0.0278 | 0.0142 |
| **Left central cLD** | 0.0266 | 0.0447 |  | 0.0447 | 0.0154 | 0.0085 | 0.0397 | 0.0245 | 0.0127 |
| **Left lateral cLD** | 0.034 | 0.0255 | 0.0447 |  | 0.0161 | 0.0355 | 0.008 | 0.0121 | 0.0263 |
| **Right cLD** | 0.0113 | 0.0176 | 0.0154 | 0.0161 |  | 0.008 | 0.0099 | 0.0039 | 0.0041 |
| **Left cLD** | 0.0075 | 0.0401 | 0.0085 | 0.0355 | 0.008 |  | 0.0318 | 0.012 | 0.0124 |
| **Lateral cLD** | 0.032 | 0.0078 | 0.0397 | 0.008 | 0.0099 | 0.0318 |  | 0.0137 | 0.0153 |
| **Central cLD** | 0.0093 | 0.0278 | 0.0245 | 0.0121 | 0.0039 | 0.012 | 0.0137 |  | 0.0137 |
| **Mean cLD** | 0.0202 | 0.0142 | 0.0127 | 0.0263 | 0.0041 | 0.0124 | 0.0153 | 0.0137 |  |

**Supplementary Table 1 | Phenotypic and genetic correlation between the different carotid lumen diameter phenotypes. a)** Phenotypic (Pearson’s, below the diagonal) and genetic (above the diagonal) correlation, and **b)** genetic standard deviation of the different estimates of carotid lumen diameter. For the phenotypic correlation analysis, we z-scored and covariate-corrected (see Methods) all phenotypes. For the genetic correlation, we qq-normalised and covariate-corrected all phenotypes. Abbreviations: cLD, carotid lumen diameter.

**a.**

|  | **Mean cLD** | **Mean cIMT** | **Mean cIMT over mean cLD** |
| --- | --- | --- | --- |
| **Mean cLD** |  | 0.58 | -0.11 |
| **Mean cIMT** | 0.370057 |  | 0.74 |
| **Mean cIMT over mean cLD** | -0.232589 | 0.810348 |  |

**b.**

|  | **Mean cLD** | **Mean cIMT** | **Mean cIMT over mean cLD** |
| --- | --- | --- | --- |
| **Mean cLD** |  | 0.1 | 0.11 |
| **Mean cIMT** | 0.1 |  | 0.05 |
| **Mean cIMT over mean cLD** | 0.11 | 0.05 |  |

**Supplementary Table 2 | Phenotypic and genetic correlation between the different carotid phenotypes. a)** Phenotypic (Pearson’s, below the diagonal) and genetic (above the diagonal) correlation, and **b)** genetic standard deviation of the different carotid phenotypes. For the phenotypic correlation analysis, we z-scored and covariate-corrected (see Methods) all phenotypes. For the genetic correlation, we qq-normalised and covariate-corrected all phenotypes. Abbreviations: cLD, carotid lumen diameter; cIMT, carotid intima-media thickness.

|  | **h2** | **std** |
| --- | --- | --- |
| **Right lateral cLD** | 0.28 | 0.05 |
| **Right central cLD** | 0.25 | 0.06 |
| **Left central cLD** | 0.22 | 0.04 |
| **Left lateral cLD** | 0.25 | 0.05 |
| **Right cLD** | 0.28 | 0.06 |
| **Left cLD** | 0.25 | 0.05 |
| **Lateral cLD** | 0.31 | 0.06 |
| **Central cLD** | 0.28 | 0.06 |
| **Mean cLD** | 0.31 | 0.06 |
| **Mean cIMT** | 0.23 | 0.04 |
| **Mean cIMT over mean cLD** | 0.14 | 0.03 |

**Supplementary Table 3 | SNP heritability (h2), computed using LDSR, from GWAS summary statistics, computed using Regenie.**

| **Characteristics** | **Description** | **Number** | **Percentage** |
| --- | --- | --- | --- |
| **Sex**  (UKB data field 21 003) | Female | 9 550 | 49.18% |
|  | Male | 9 279 | 47.78% |
|  | NA | 590 | 3.04% |
| **Age at Imaging (instance 2)**  (UKB data field 22 001) | Median | 55 |  |
|  | Range | 40 – 70 |  |
| **Standing height (instance 2)**  (UKB data field 50) | Median | 169 |  |
|  | Range | 134 – 202 |  |
|  | NA | 309 | 1.52% |
| **BMI (instance 2)**  (UKB data field 21 001) | Median | 25.95 |  |
|  | Range | 13.39 – 69.63 |  |
|  | NA | 340 | 1.75% |
| **Ethnicity**  (UKB data field 21 000) | African | 39 | 0.25% |
|  | Any other Asian background | 20 | 0.13% |
|  | Any other Black background | 3 | 0.02% |
|  | Any other mixed background | 19 | 0.12% |
|  | Any other white background | 383 | 2.47% |
|  | Asian or Asian British | 1 | 0.01% |
|  | Bangladeshi | 2 | 0.01% |
|  | British | 14 323 | 92.19% |
|  | Caribbean | 48 | 0.31% |
|  | Chinese | 48 | 0.31% |
|  | Do not know | 2 | 0.01% |
|  | Indian | 95 | 0.61% |
|  | Irish | 384 | 2.47% |
|  | Mixed | 2 | 0.01% |
|  | Other ethnic group | 53 | 0.34% |
|  | Pakistani | 30 | 0.19% |
|  | Prefer not to answer | 31 | 0.20% |
|  | White | 11 | 0.07% |
|  | White and Asian | 16 | 0.10% |
|  | White and Black African | 8 | 0.05% |
|  | White and Black Caribbean | 15 | 0.10% |
|  | NA | 8 | 0.04% |

**Supplementary Table 4 | Main demographics of the carotid ultrasound cohort (N=19** **419).** Abbreviations: BMI, body mass index; NA, not available; UKB, UK Biobank.

| **Phenotype** | **Chr** | **Variant** | **p** | **Additional Information** |
| --- | --- | --- | --- | --- |
| **Mean  cLD** | 4 | rs782703932 | 1.41E-08 | NA |
|  | 6 | rs144497254 | 3.26E-08 | NA |
|  | 7 | rs343029 | 2.73E-26 | Previously associated with cIMT ([EBI GWAS Catalog](https://www.ebi.ac.uk/gwas/variants/rs343029)) |
|  | 8 | 8:8703781_GGA_G | 4.68E-10 | NA |
|  | “ | rs113906777 | 4.82E-08 | NA |
|  | “ | rs13254263 | 7.63E-10 | NA |
|  | “ | rs7844895 | 2.45E-11 | NA |
|  | “ | rs7838131 | 3.20E-08 | Linked to **BP, BMI** ([EBI GWAS Catalog](https://www.ebi.ac.uk/gwas/variants/rs7838131)), and **CAD** |
|  | 10 | rs57178050 | 3.31E-08 | Genes: **CC2D2B, RPL21P90, CCNJ, MIR3157** |
|  | 11 | rs111677878 | 8.62E-12 | NA |
|  | 12 | rs11108966 | 2.41E-08 | NA |
| **Mean  cIMT** | 2 | rs374418010 | 1.58E-08 | NA |
|  | 3 | rs146453480 | 7.94E-09 | NA |
|  | 5 | rs559857649 | 1.35E-08 | NA |
|  | 7 | rs342988 | 4.25E-10 | Previously associated with cIMT |
|  | 8 | rs2572397 | 3.76E-08 | [Genes: LINC00529, RPL19P13, EBI GWAS Catalog](https://www.ebi.ac.uk/gwas/variants/rs2572397) |
|  | 11 | rs974819 | 2.11E-08 | [CHD association](https://www.sciencedirect.com/science/article/pii/S0049384812002459), [SNPedia](https://www.snpedia.com/index.php/Rs974819) |
|  | 13 | rs11617955 | 2.86E-08 | [EBI GWAS Catalog](https://www.ebi.ac.uk/gwas/variants/rs11617955) |
|  | 16 | rs548591 | 1.34E-08 | [Found in BioRxiv](https://www.biorxiv.org/content/10.1101/718684v1.full.pdf) |
|  | 19 | rs1065853 | 8.99E-11 | Previously associated with cIMT, and associated with **lipoprotein metabolism** [GWAS](https://www.ebi.ac.uk/gwas/variants/rs1065853) |
| **Mean cIMT over mean cLD** | 7 | rs7792074 | 5.21E-09 | NA |
|  | 8 | rs574485439 | 1.53E-08 | NA |
|  | 15 | rs625034 | 8.16E-11 | [Associated with TAA, BioRxiv](https://www.biorxiv.org/content/10.1101/2021.02.05.429911v1.full.pdf) |
|  | 19 | rs111688353 | 3.11E-08 | NA |
|  | “ | rs1065853 | 2.10E-10 | Previously associated with cIMT, and associated with **lipoprotein metabolism** [GWAS](https://www.ebi.ac.uk/gwas/variants/rs1065853) |
|  | 21 | rs62208686 | 3.72E-08 | NA |

**Supplementary Table 5 | Genetic variants associated with mean cIMT, mean cLD, and the ratio of mean cIMT to mean cLD.** The table lists the Chr and variant ID (e.g., rsID) for each variant and additional information on prior associations with cardiovascular traits, diseases, or relevant genes. Notable associations include links to conditions such as coronary heart disease (CHD), atherosclerosis, and lipoprotein metabolism, referencing external databases like the EBI GWAS Catalogue, SNPedia, and BioRxiv. Abbreviations: NA, not available; BP, blood pressure; BMI: body mass index; CAD: coronary artery disease.

| **Phenotype** | **Instance 2** | **Instance 3** | **Both instances** | **Only instance 3** |
| --- | --- | --- | --- | --- |
| **Right Lateral cLD** | 18 542 | 2 047 | 1 657 | 390 |
| **Right Central cLD** | 18 430 | 2 041 | 1 633 | 408 |
| **Left Central cLD** | 18 351 | 2 047 | 1 628 | 419 |
| **Left Lateral cLD** | 18 259 | 2 064 | 1 652 | 412 |
| **Mean cLD** | 19 429 | 2 151 | 1 805 | 346 |

**Supplementary Table 6 | Number of phenotypes available for each instance (2 and 3) and their overlap.** Here, "Instance 2" refers to the total number of measurements available at instance 2, "Instance 3" refers to the total measurements at instance 3, "Both instances" indicates the number of measurements present in both instances for the same subjects, and "Only instance 3" represents measurements available exclusively in instance 3. All of the numbers refer to the dataset before applying standard outlier removal and filtering for complete cases (with all the covariates).

**a.**

| **Disease/ Risk** | **UKB Data Field** | **Field Name** |
| --- | --- | --- |
| Age | 21003 | Age at assessment |
| Sex | 22001 | Sex |
| Height | 50 | Standing height |
| BMI | 21001 | BMI |
| Systolic Blood Pressure | 4080 | Systolic blood pressure  (automated reading) |
| Diastolic Blood Pressure | 4079 | Diastolic blood pressure  (automated reading) |
| Smoking status | 20116 | Smoking status |
| Smoking Pack-years | 20161 | Pack-years of smoking |
| Diabetes | 2976 | Age diabetes diagnosed |
|  | 30740 | Glucose |
|  | 30750 | Glycated haemoglobin (HbA1c) |
|  | 2443 | Diabetes diagnosed by doctor |
| Resting Heart Rate | 102 | Pulse rate, automated reading |
| Arterial Stiffness | 21021 | Pulse Wave Arterial Stiffness Index |
| Hypertension | 6153, 6177 | Medication for cholesterol, blood pressure, diabetes (or take exogenous hormones) |
|  | 4079, 4080 | Systolic and diastolic blood pressure |

**b.**

| **Risk Factor** | **UKB Data Field** | **Field Name** |
| --- | --- | --- |
| Medicated for Hypertension | 6153 | Medication for cholesterol, blood pressure, diabetes, or take exogenous hormones |
|  | 6177 | Medication for cholesterol, blood pressure or diabetes |
| Previous Cardiovascular Event | 131296, 131304, 131306, 131298, 131300, 131302, 42000, 42006, 42008, 42010, 131338, 131340, 131288, 131292 | (see **Suppl. Table 5**) |

**Supplementary Table 7.** Data fields used to define risk factors for **a)** the disease association analysis and, additionally, for **b)** the survival analysis.

| **Event Category** | **UKB Data Field** | **Field Name** |
| --- | --- | --- |
| **IHD** (Ischaemic Heart Disease) | 131296 | Date angina first reported |
|  | 131304 | Date other acute IHD first reported |
|  | 131306 | Date IHD first reported |
| **MI**  (Myocardial Infarction) | 131298 | Date acute MI first reported |
|  | 131300 | Date subsequent MI first reported |
|  | 131302 | Date complications after MI first reported |
|  | 42000 | Date of myocardial infarction |
| **Stroke** | 42006 | Date of stroke |
|  | 42008 | Date of ischaemic stroke |
|  | 42010 | Date of intracerebral haemorrhage |
| **NIC** (Non-ischaemic Cardiomyopathies) | 131338 | Date cardiomyopathy first reported |
|  | 131340 | Date cardiomyopathy in other diseases first reported |
|  | 131288 | Date hypertensive heart disease first reported |
|  | 131292 | Date hypertensive heart and renal disease first reported |
| **Thrombotic Event** | 131308 | Date pulmonary embolism first reported |
|  | 131388 | Date arterial embolism and thrombosis first reported |
|  | 131400 | Date other venous embolism and thrombosis first reported |

**Supplementary Table 8 | Data fields used to define prevalent disease outcomes for the disease association analysis and cardiovascular endpoints for the survival analysis.** In the survival analysis, we considered only events reported after the images were acquired (Date of attending assessment centre, Data Field 53).

| **Risk Factor** | **Mean (IQR)** |
| --- | --- |
| Age | 54.9 (12) |
| BMI | 26.6 (5.3) |
| SBP | 139.3 (24) |

| **Risk Factor** | **Prevalence** | **Percentage** |
| --- | --- | --- |
| Sex: male | 9245 | 49.3% |
| Alcohol: Never | 1308 | 7.0% |
| Occasional | 4065 | 21.6% |
| Regular | 13284 | 70.8% |
| NA | 110 | 0.6% |
| Smoking: Never | 11656 | 62.1% |
| Previous | 6283 | 33.5% |
| Current | 662 | 3.5% |
| NA | 166 | 0.9% |
| Diabetes (diagnosis) | 1043 | 5.6% |
| Medicated for Hypertension | 4418 | 23.5% |
| Previous Cardiovascular Event | 1592 | 8.5% |

**Supplementary Table 9.** Baseline characteristics of the survival analysis cohort (N=18 767).

**a.**

| **Event** | **Prevalent** | **Incident (at 6 years)** | **Incident (at 6 years)** |
| --- | --- | --- | --- |
|  | **N = 18 767** | | **N = 15 011** |
| IHD | 1613 | 562 | 439 |
| MI | 718 | 223 | 167 |
| Stroke | 416 | 179 | 129 |
| Thrombotic Event | 335 | 121 | 93 |
| NIC | 84 | 39 | 30 |
| Cardiovascular Event (first event only) | 2305 | 893 | 685 |
| All-cause mortality | 502 | 441 | 329 |

**b.**

| **Event (M/F)** | **Prevalent** | **Incident (at 6 years)** | **Incident (at 6 years)** |
| --- | --- | --- | --- |
|  | **N_F_ = 9 522; N_M_ = 9 245** | | **N_F_ = 7 582; N_M_ = 7 429** |
| IHD | 459/737 | 165/397 | 130/309 |
| MI | 128/590 | 50/173 | 38/129 |
| Stroke | 153/263 | 73/106 | 52/77 |
| Thrombotic Event | 146/189 | 50/71 | 34/59 |
| NIC | 25/59 | 8/31 | 6/24 |
| Cardiovascular Event (first event only) | 737/1568 | 287/606 | 221/464 |
| All-cause mortality | 71/331 | 146/295 | 109/220 |

**Supplementary Table 10.** **a)** Prevalence and incidence of cardiovascular events for the cohorts used in the disease association and the survival analysis (before and after adjusting for all the covariates; N=15 011 are relative to the cohort in the fully adjusted Cox model analyses, after excluding non-complete observations). When combining cardiovascular events (last row), we kept only the first occurrence after imaging for subjects with multiple events. **b)** The same data, stratified by sex. Here, N_F_ indicates the sample size for females, while N_M_ indicates the sample size for males.

**a.**

| **Risk Factor** | **Phenotype** | **Standardised β (95% CI)** | **p-value** |
| --- | --- | --- | --- |
| DBP  (un-medicated) | cIMT | 0.04202 (0.02155–0.06249) | p<0.0001 |
|  | cLD | 0.1605 (0.1384–0.1827) | p<0.0001 |
|  | cIMT/cLD | -0.03518 (-0.05432– -0.01604) | p<0.001 |
| SBP  (un-medicated) | cIMT | 0.2192 (0.1999–0.2385) | p<0.0001 |
|  | cLD | 0.2594 (0.2385–0.2803) | p<0.0001 |
|  | cIMT/cLD | 0.08299 (0.06467–0.1013) | p<0.0001 |
| DBP  (medicated) | cIMT | -0.07285 (-0.1048– -0.04085) | p<0.0001 |
|  | cLD | 0.03793 (0.004988–0.07087) | p<0.05 |
|  | cIMT/cLD | -0.08483 (-0.1148– -0.05484) | p<0.0001 |
| SBP  (un-medicated) | cIMT | 0.12 (0.08738–0.1526) | p<0.0001 |
|  | cLD | 0.1499 (0.1165–0.1832) | p<0.0001 |
|  | cIMT/cLD | 0.02692 (-0.003875–0.05773) | p=0.08663 |
| Smoking  pack-years | cIMT | 0.107 (0.07913–0.1348) | p<0.0001 |
|  | cLD | 0.08438 (0.05523–0.1135) | p<0.0001 |
|  | cIMT/cLD | 0.05665 (0.03037–0.08293) | p<0.0001 |
| Arterial  Stiffness | cIMT | 0.0413 (0.02648–0.05611) | p<0.0001 |
|  | cLD | 0.03082 (0.01523–0.0464) | p<0.001 |
|  | cIMT/cLD | 0.02343 (0.009555–0.0373) | p<0.001 |
| Resting  Heart Rate | cIMT | -0.08551 (-0.1027– -0.06832) | p<0.0001 |
|  | cLD | 0.05444 (0.03627–0.07261) | p<0.0001 |
|  | cIMT/cLD | -0.1032 (-0.1192– -0.08716) | p<0.0001 |
| BMI | cIMT | 0.108 (0.09279–0.1233) | p<0.0001 |
|  | cLD | 0.2338 (0.2181–0.2496) | p<0.0001 |
|  | cIMT/cLD | -0.01433 (-0.02864–-0.00002091) | p<0.05 |

**b.**

| **Trait** | **Phenotype** | **Odds Ratio (95% CI)** | **p-value** |
| --- | --- | --- | --- |
| Smoking  (previous) | cIMT | 1.137 (1.098–1.176) | p<0.0001 |
|  | cLD | 1.138 (1.097–1.18) | p<0.0001 |
|  | cIMT/cLD | 1.054 (1.021–1.089) | p<0.01 |
| Smoking  (current) | cIMT | 1.212 (1.113–1.32) | p<0.0001 |
|  | cLD | 1.251 (1.148–1.365) | p<0.0001 |
|  | cIMT/cLD | 1.069 (0.984–1.161) | p=0.1147 |
| All-cause  mortality | cIMT | 1.042 (0.9512–1.141) | p=0.3773 |
|  | cLD | 1.07 (0.9725–1.177) | p=0.1652 |
|  | cIMT/cLD | 1.004 (0.9195–1.095) | p=0.9369 |
| CVD | cIMT | 1.085 (1.036–1.136) | p<0.001 |
|  | cLD | 1.122 (1.069–1.178) | p<0.0001 |
|  | cIMT/cLD | 1.028 (0.9835–1.075) | p=0.2207 |
| Stroke | cIMT | 1.207 (1.097–1.329) | p<0.001 |
|  | cLD | 1.34 (1.217–1.476) | p<0.0001 |
|  | cIMT/cLD | 1.033 (0.9389–1.137) | p=0.5048 |
| Miocardial  Infarction | cIMT | 1.06 (0.9828–1.143) | p=0.1312 |
|  | cLD | 1.063 (0.9814–1.151) | p=0.134 |
|  | cIMT/cLD | 1.043 (0.97–1.122) | p=0.2557 |
| Ischaemic  Heart Disease | cIMT | 1.08 (1.024–1.139) | p<0.01 |
|  | cLD | 1.107 (1.047–1.171) | p<0.001 |
|  | cIMT/cLD | 1.031 (0.9793–1.085) | p=0.2467 |
| Diabetes | cIMT | 1.125 (1.055–1.2) | p<0.001 |
|  | cLD | 1.364 (1.279–1.454) | p<0.0001 |
|  | cIMT/cLD | 0.9559 (0.8964–1.019) | p=0.1695 |
| Hypertension | cIMT | 1.401 (1.347–1.458) | p<0.0001 |
|  | cLD | 1.818 (1.739–1.901) | p<0.0001 |
|  | cIMT/cLD | 1.038 (1.002–1.075) | p<0.05 |

**Supplementary Table 11. Diagnostic performance of mean cIMT, mean cLD, and their ratio. a)** Standardised regression coefficients from models assessing the independent associations of each phenotype with major continuous risk factors (adjusted for age, sex, and height; N=18 767). **b)** Odds ratios from analogous models evaluating associations with major categorical risk factors and prevalent disease outcomes (adjusted for age, sex, and height; N=18 767). Abbreviations: SBP/DBP: systolic/diastolic blood pressure; BMI: body mass index; CVD: cardiovascular diseases; MI: myocardial infarction; IHD: ischaemic heart disease.

**a.**

| **Risk Factor** | **Phenotype** | **Standardised β_F_ & β_M_ (95% CI)** | **p-value (F/M)** |
| --- | --- | --- | --- |
| DBP  (un-medicated) | cIMT | 0.0447 (0.0131–0.0764)  0.0398 (0.0132–0.0664) | p<0.01  p<0.01 |
|  | cLD | 0.1775 (0.1446–0.2104)  0.1418 (0.1119–0.1716) | p<0.0001  p<0.0001 |
|  | cIMT/cLD | -0.04285 (-0.07049– -0.01521)  -0.02683 (-0.05324– -0.0004245) | p<0.01  p<0.05 |
| SBP  (un-medicated) | cIMT | 0.2732 (0.2432–0.3032)  0.178 (0.1533–0.2027) | p<0.0001  p<0.0001 |
|  | cLD | 0.2751 (0.2437–0.3065)  0.24 (0.2124–0.2676) | p<0.0001  p<0.0001 |
|  | cIMT/cLD | 0.1016 (0.07489–0.1284)  0.06607 (0.04125–0.09089) | p<0.0001  p<0.0001 |
| DBP  (medicated) | cIMT | -0.1215 (-0.1789– -0.06406)  -0.05247 (-0.09103– -0.01392) | p<0.0001  p<0.01 |
|  | cLD | 0.06725 (0.005207–0.1293)  0.02757 (-0.01135–0.06649) | p<0.05  p=0.1649 |
|  | cIMT/cLD | -0.1292 (-0.1809–-0.07752)  -0.06444 (-0.1013–-0.02761) | p<0.0001  p<0.001 |
| SBP  (un-medicated) | cIMT | 0.1498 (0.08893–0.2107)  0.1059 (0.06765–0.1443) | p<0.0001  p<0.0001 |
|  | cLD | 0.18 (0.1147–0.2453)  0.1396 (0.1011–0.178) | p<0.0001  p<0.0001 |
|  | cIMT/cLD | 0.04641 (-0.008866–0.1017)  0.01558 (-0.0213–0.05246) | p=0.09977  p=0.4075 |
| Smoking  pack-years | cIMT | 0.1047 (0.06146–0.1479)  0.1082 (0.07149–0.1448) | p<0.0001  p<0.0001 |
|  | cLD | 0.06384 (0.01744–0.1102)  0.09441 (0.05651–0.1323) | p<0.01  p<0.0001 |
|  | cIMT/cLD | 0.06124 (0.0225–0.09999)  0.05396 (0.01835–0.08956) | p<0.01  p<0.01 |
| Arterial  Stiffness | cIMT | 0.04261 (0.01824–0.06698)  0.04053 (0.02207–0.05899) | p<0.001  p<0.0001 |
|  | cLD | 0.02853 (0.003465–0.0536)  0.03217 (0.01244–0.05189) | p<0.05  p<0.01 |
|  | cIMT/cLD | 0.0219 (0.0003972–0.0434)  0.02476 (0.006736–0.04279) | p<0.05  p<0.01 |
| Resting  Heart Rate | cIMT | -0.08613 (-0.1124– -0.05985)  -0.08553 (-0.1085– -0.06256) | p<0.0001  p<0.0001 |
|  | cLD | 0.06702 (0.0397–0.09435)  0.04551 (0.02095–0.07007) | p<0.0001  p<0.001 |
|  | cIMT/cLD | -0.1052 (-0.1282–-0.08215)  -0.102 (-0.1243–-0.07959) | p<0.0001  p<0.0001 |
| BMI | cIMT | 0.1287 (0.1026–0.1549)  0.09523 (0.07734–0.1131) | p<0.0001  p<0.0001 |
|  | cLD | 0.2122 (0.1853–0.2391)  0.2473 (0.2288–0.2658) | p<0.0001  p<0.0001 |
|  | cIMT/cLD | 0.005826 (-0.0173–0.02896)  -0.0293 (-0.04685–-0.01176) | p=0.6215  p<0.01 |

**b.**

| **Trait** | **Phenotype** | **OR_F_ & OR_M_ (95% CI)** | **p-value (F/M)** |
| --- | --- | --- | --- |
| Smoking  (previous) | cIMT | 1.086 (1.028–1.148)  1.171 (1.121–1.224) | p<0.01  p<0.0001 |
|  | cLD | 1.101 (1.039–1.167)  1.166 (1.113–1.222) | p<0.01  p<0.0001 |
|  | cIMT/cLD | 1.025 (0.9753–1.077)  1.077 (1.032–1.124) | p=0.3334  p<0.001 |
| Smoking  (current) | cIMT | 0.9912 (0.8443–1.164)  1.329 (1.2–1.47) | p=0.914  p<0.0001 |
|  | cLD | 1.112 (0.9561–1.294)  1.337 (1.203–1.485) | p=0.1682  p<0.0001 |
|  | cIMT/cLD | 0.9442 (0.8201–1.087)  1.147 (1.035–1.27) | p=0.424  p<0.01 |
| All-cause  mortality | cIMT | 0.9155 (0.761–1.101)  1.088 (0.9797–1.208) | p=0.3492  p=0.115 |
|  | cLD | 1.033 (0.8529–1.251)  1.084 (0.9706–1.21) | p=0.74  p=0.153 |
|  | cIMT/cLD | 0.9244 (0.7834–1.091)  1.036 (0.935–1.149) | p=0.3516  p=0.496 |
| CVD | cIMT | 1.047 (0.9564–1.146)  1.099 (1.041–1.159) | p=0.3192  p<0.001 |
|  | cLD | 1.226 (1.117–1.346)  1.086 (1.026–1.15) | p<0.0001  p<0.01 |
|  | cIMT/cLD | 0.9456 (0.8702–1.028)  1.064 (1.01–1.122) | p=0.187  p<0.05 |
| Stroke | cIMT | 1.041 (0.8618–1.258)  1.276 (1.14–1.427) | p=0.6744  p<0.0001 |
|  | cLD | 1.196 (0.9883–1.448)  1.4 (1.252–1.567) | p=0.06585  p<0.0001 |
|  | cIMT/cLD | 0.9576 (0.8047–1.14)  1.067 (0.952–1.197) | p=0.6256  p=0.2637 |
| Myocardial Infarction | cIMT | 1.187 (0.9758–1.444)  1.04 (0.9584–1.128) | p=0.08639  p=0.3475 |
|  | cLD | 1.333 (1.093–1.624)  1.022 (0.9373–1.115) | p<0.01  p=0.6212 |
|  | cIMT/cLD | 1.023 (0.8502–1.23)  1.047 (0.9673–1.133) | p=0.8118  p=0.257 |
| Ischaemic  Heart Disease | cIMT | 1.104 (0.9892–1.232)  1.072 (1.009–1.14) | p=0.07739  p<0.05 |
|  | cLD | 1.321 (1.181–1.478)  1.048 (0.9824–1.117) | p<0.0001  p=0.1556 |
|  | cIMT/cLD | 0.9553 (0.8624–1.058)  1.058 (0.9975–1.123) | p=0.3813  p=0.06044 |
| Diabetes | cIMT | 1.249 (1.101–1.416)  1.086 (1.008–1.171) | p<0.001  p<0.05 |
|  | cLD | 1.477 (1.303–1.674)  1.327 (1.232–1.431) | p<0.0001  p<0.0001 |
|  | cIMT/cLD | 1.004 (0.8901–1.132)  0.9373 (0.8685–1.012) | p=0.9526  p=0.09575 |
| Hypertension | cIMT | 1.531 (1.439–1.628)  1.313 (1.248–1.382) | p<0.0001  p<0.0001 |
|  | cLD | 1.888 (1.766–2.017)  1.754 (1.652–1.863) | p<0.0001  p<0.0001 |
|  | cIMT/cLD | 1.069 (1.016–1.126)  1.011 (0.9648–1.06) | p<0.05  p=0.6375 |

**Supplementary Table 12. Diagnostic performance of mean cIMT, mean cLD, and their ratio in females and males. a)** Standardised regression coefficients from models assessing the independent associations of each phenotype with major continuous risk factors (adjusted for age and height). Females (N_F_ = 9 522) are reported at the top of each row and males (N_M_ = 9 245) at the bottom. **b)** Odds ratios from analogous models evaluating associations with major categorical risk factors and prevalent disease outcomes (adjusted for age and height). Abbreviations: SBP/DBP: systolic/diastolic blood pressure; BMI: body mass index; MI: myocardial infarction; IHD: ischaemic heart disease.
